# Place or placeholder? AlphaEarth satellite embeddings did not detectably improve weekly dengue forecasts in four countries

**DOI:** 10.64898/2026.11.23.26368794

**Authors:** Brij K. Patel, Adithya V. Asokan, Santosh Pant

**Affiliations:** Computer Science, Illinois State University, Normal, Illinois, USA; Department of Health Metrics Sciences, University of Washington, Seattle, Washington, USA; Knox College, Galesburg, Illinois, USA

**Keywords:** dengue, probabilistic forecasting, satellite embeddings, geospatial foundation models, location confounding, spatial validation, equivalence testing

## Abstract

**Background:** Some recent disease forecasts use satellite embeddings (numbers summarising places). A model may use them only as a placeholder, recognising districts and their usual dengue levels without learning about the environment. We asked whether AlphaEarth embeddings improve weekly district dengue forecasts and whether gains go beyond such recognition.

**Methods:** We retrospectively forecast cases one and four weeks ahead in 6,909 districts of Brazil, Colombia, Peru and Taiwan (OpenDengue, 2020–2024) with gradient boosting. In 14 comparisons across future years and held-out places, we measured how embeddings changed the weighted interval score, with and without a location control (an input that only identifies a district), to test whether location alone explains gains. We used Diebold–Mariano tests, the Holm correction and a ±1% equivalence margin set after the first Brazil and Peru results.

**Findings:** No comparison showed a significant gain after correction. Six showed losses: five small (−0.42% to −0.68%) and one with Taiwan held out (−18.3%; absolute −0.0037). Only Colombia’s held-out-states loss (−0.52%) stayed significant under two stricter checks and a check for cases hidden in zero-filled weeks. In 8 comparisons (5 under the stricter checks), any gain or loss was shown to be smaller than 1% of forecast error. This margin is roughly Brazil’s gain from all non-case inputs combined. In exploratory analyses, embeddings alone predicted district burden in unseen states. Without case history, neither coordinates nor embeddings helped detectably in Brazil’s held-out states; both hurt in Colombia’s. In Brazil’s future years, coordinates cut error by 3.3% (1.7 to 4.9); embeddings added nothing detectable.

**Interpretation:** Embeddings partly showed which districts usually have more dengue, but in Brazil and Colombia other non-case inputs already held most of it. With case history they added no detectable accuracy; without it, evidence was inconclusive. Embeddings should be tested against location-only controls in held-out places.

## 1 Introduction

Dengue causes a large burden of illness in the tropics and subtropics. An estimated 390 million dengue infections occur each year, and 96 million of them cause symptoms [1]. Cases reported to the World Health Organization reached a record of more than 14.6 million in 2024, with more than 12,000 deaths [2]. Peru had the largest dengue outbreak on its record in 2023 [3], and Brazil its largest epidemic to date in 2024 [4]. Short-term forecasts for small areas can help health services time vector control and prepare clinical care. Dengue forecasts that give a range of likely values, rather than a single number, are well established [5–8].

Dengue transmission depends partly on climate and the environment [6,7] and on human movement [9]. Forecasters therefore often add descriptors of place from separate data products. Geospatial foundation models are trained on large archives of satellite data and other observations. For any location, they compress these observations into a fixed-length list of numbers, called an embedding. AlphaEarth Foundations provides a 64-number embedding for every 10 m pixel and every year from 2017 [10,11]. It is computed from optical and radar satellite imagery by a model that was also trained to reconstruct climate, elevation and other layers. The Population Dynamics Foundation Model (PDFM) provides place embeddings built from search, map and activity data [12]. Even generic image features predict many outcomes [13,14]. Because embeddings cover the whole globe, they are attractive where local data are sparse.

An embedding can also improve a forecast without telling the model anything about the environment. A district’s embedding changes little from year to year, so a flexible model can use it to recognise the district and learn its usual dengue level. For example, it can learn that one district usually has more dengue than another without learning why. The embedding is then only a placeholder: a label that tells the model which district it is looking at. When every test district also appears in training, this labelling role alone can improve scores. We call this the identity confound: an apparent gain that comes from knowing which district is which. In spatial machine learning, coordinates and similar stand-ins for place help on places a model has seen but work poorly on new ones [15,16]. In England, much of the gain from satellite-image embeddings for house prices, but not for air pollution, reflected coarse location [17].

The identity confound can be tested with location-only features and with places never seen in training. Dengue adds a complication. Our models see a district’s cases in recent weeks and in the same weeks of the previous three years. These show roughly its usual dengue level, which is much of what a label could tell a model. Holding districts out of training stops a model from memorising which embedding belongs to which district. When it forecasts for a held-out district, however, it still receives that district’s own recent case counts. The cleanest test of whether embeddings carry environmental information or merely identify a district therefore needs a model without case history, which cannot read a district’s usual level from its counts. It also needs held-out places, where the model cannot have memorised the district.

Published dengue evaluations of place embeddings have not yet applied these tests. Brittain et al. added one PDFM snapshot to monthly forecasts for 5,570 Brazilian municipalities as a residual adjustment, with temporal hold-outs only [18]. Forecast error fell in 19 of 20 combinations of model and horizon. In the Mexico dengue case study of Muslim et al., a static embedding reduced the mean weighted interval score by 0.0051 at one month, and the gain was not significant at three or six months [19]. For two other outcomes, malaria and respiratory infection, Nazir et al. reported AlphaEarth gains [20,21]. In none of these analyses did we find a location-only control, such as coordinates, district labels or shuffled embeddings, or a test in held-out places, although other case studies in the Muslim et al. paper had such checks. The closest earlier study, of dengue early warning in one Indian city, found that the embeddings of a geospatial foundation model could not be distinguished from open map layers, and that the difference “fell to 0.001 once the open layers were given the cell’s own coordinates” [22]. We extend that test to four countries, to weekly forecasts that give a range of values, and to places held out of training.

In a retrospective study, we tested whether AlphaEarth embeddings improve weekly dengue forecasts in 6,909 scored second-level administrative units of Brazil, Colombia, Peru and Taiwan. These are municipalities in Brazil and Colombia, provinces in Peru and townships in Taiwan; we call them all districts. A location control is an input that tells the model where a district is, which district it is, or how much dengue it usually has, without describing its own environment directly.

Our main question was whether the embeddings improve forecasts. Location controls and analyses without case history bear on a second question, whether a gain reflects the environment or only district recognition, but cannot fully settle it. We turned these into four questions. When the test covers only future years, do embeddings reduce forecast error beyond the other inputs? Is any gain left once a location control is present? Does any gain appear in districts, states or countries left out of training, although their recent case counts remain inputs? Finally, in exploratory analyses, do embeddings predict a district’s average dengue burden, and without case history, do they add anything beyond location?

## 2 Methods

### 2.1 Study design

This retrospective study used one snapshot of final, revised data and followed the EPIFORGE 2020 guidelines

[23] (Supplementary Methods, section G; design in Figure 1). Automated AI agents wrote the code and ran the downloads, checks, model runs and analyses (see Use of AI tools). Codes for models, controls and splits are defined in the glossary (Supplementary Methods, section H). Each forecast is made at a forecast origin: the end of the last week whose data it may use. From the end of week 6, for example, the forecasts one and four weeks ahead are for weeks 7 and 10. We call one country with one evaluation split a comparison. Brazil on future years is one comparison and Brazil on held-out states another.

**Figure 1.**
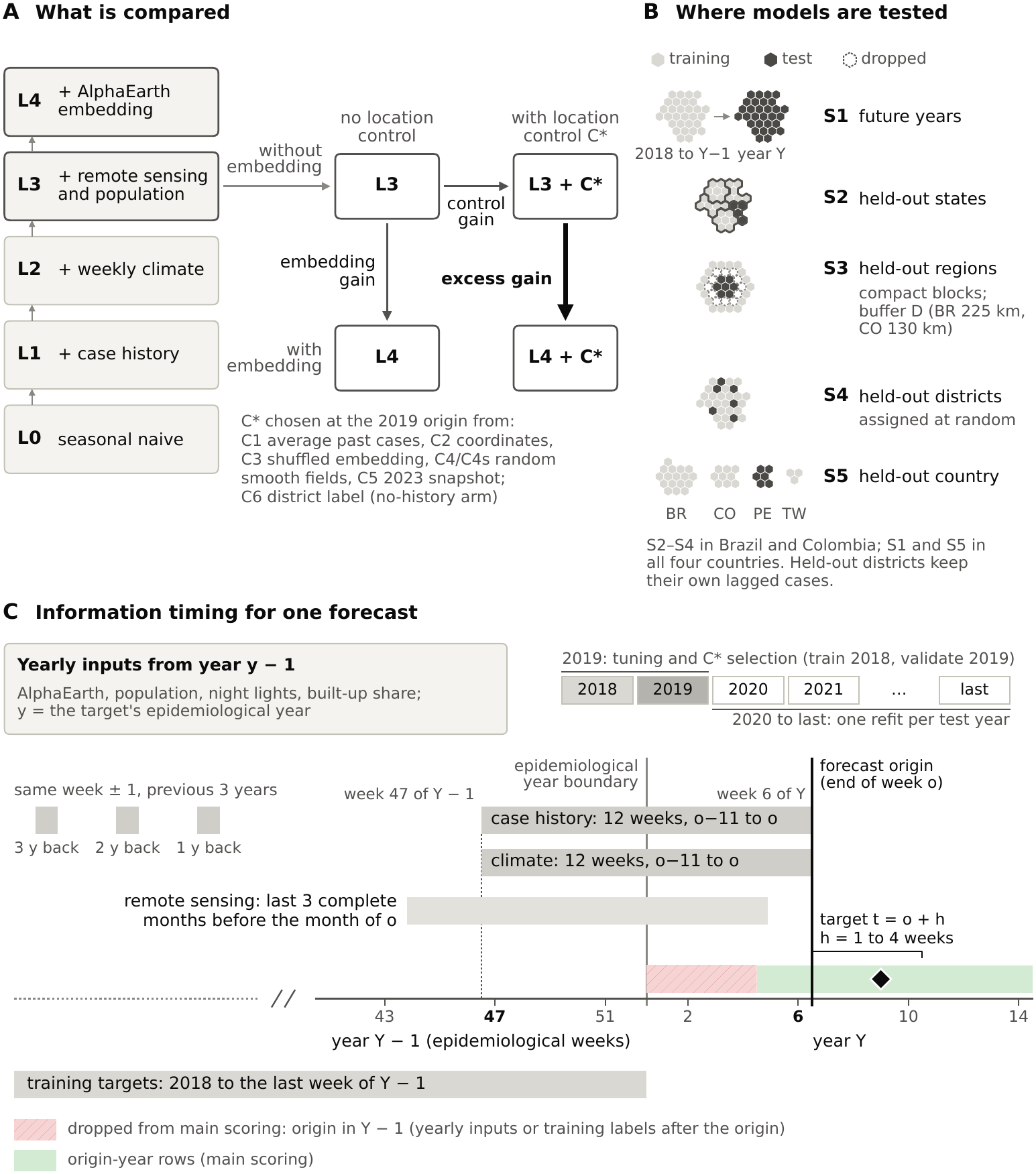
Study design and information timing. Each comparison changes one block of inputs while the model type, scored rows and settings stay fixed; scores use only forecasts whose inputs and training labels end at the origin. (A) The model ladder L0 to L4 and, from the reference model L3, the 2 × 2 grid of models behind the three gains. In the grid, each arrow adds one input (the embeddings or the location control) to the model it starts from, and the gain is how much forecast error falls along that arrow: embedding gain (L3 to L4), control gain (L3 to L3 + C*) and excess gain (L3 + C* to L4 + C*). The location control C* is chosen on 2019 data from C1 to C5, the last being the 2023 snapshot; the district label C6 is a candidate only without case history. (B) The five splits, drawn on hexagons standing for districts (light: training; dark: test; dashed: dropped buffer): S1, future years; S2, held-out states; S3, held-out regions; S4, held-out districts; S5, held-out country. S2 to S4 were run in Brazil and Colombia, S1 and S5 in all four countries. (C) Information timing for a forecast whose origin o is the end of epidemiological week 6 of year Y. The 12-week case-history and climate windows run from week 47 of year Y − 1 to week 6 of year Y; remote sensing uses the last three complete months before the month of o; yearly inputs come from the year before the target’s epidemiological year; training targets end in the last week of Y − 1. Targets in the first h weeks of Y (hatched) are dropped from scoring. Horizons are 1 to 4 weeks; main estimates pool horizons 1 and 4.

### 2.2 Data

We used weekly district case counts from OpenDengue version 1.3.1 [24,25] (Table 1; Tables S1 and S2). Because countries define cases differently, each percentage compares models within one country.

**Table 1.**
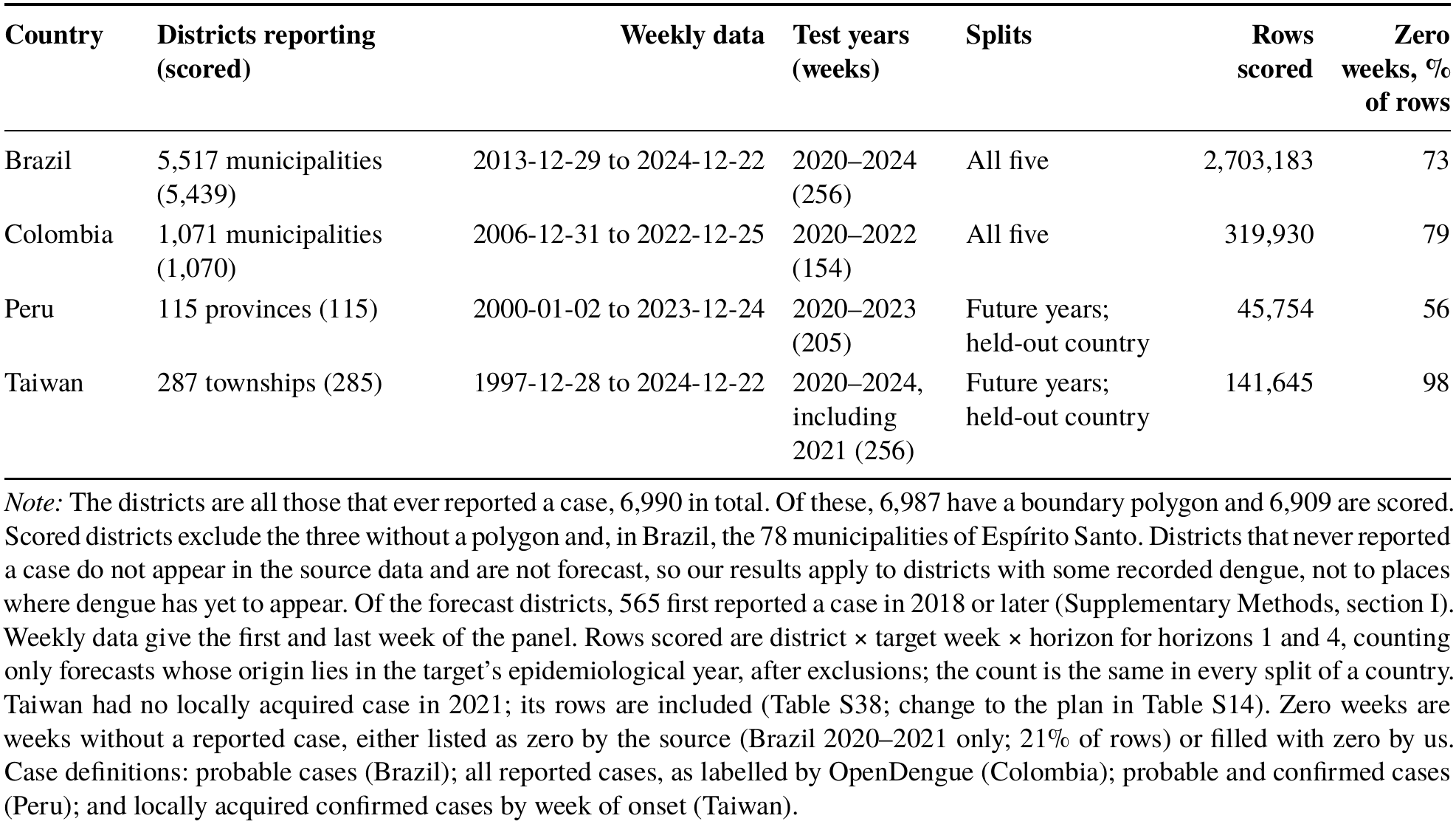
Data and evaluation design by country.

| Country | Districts reporting (scored) | Weekly data | Test years (weeks) | Splits | Rows scored | Zero weeks, % of rows |
|---|---|---|---|---|---|---|
| Brazil | 5,517 municipalities (5,439) | 2013-12-29 to 2024-12-22 | 2020–2024 (256) | All five | 2,703,183 | 73 |
| Colombia | 1,071 municipalities (1,070) | 2006-12-31 to 2022-12-25 | 2020–2022 (154) | All five | 319,930 | 79 |
| Peru | 115 provinces (115) | 2000-01-02 to 2023-12-24 | 2020–2023 (205) | Future years; held-out country | 45,754 | 56 |
| Taiwan | 287 townships (285) | 1997-12-28 to 2024-12-22 | 2020–2024, including 2021 (256) | Future years; held-out country | 141,645 | 98 |

*Note:* The districts are all those that ever reported a case, 6,990 in total. Of these, 6,987 have a boundary polygon and 6,909 are scored. Scored districts exclude the three without a polygon and, in Brazil, the 78 municipalities of Espírito Santo. Districts that never reported a case do not appear in the source data and are not forecast, so our results apply to districts with some recorded dengue, not to places where dengue has yet to appear. Of the forecast districts, 565 first reported a case in 2018 or later (Supplementary Methods, section I). Weekly data give the first and last week of the panel. Rows scored are district × target week × horizon for horizons 1 and 4, counting only forecasts whose origin lies in the target's epidemiological year, after exclusions; the count is the same in every split of a country. Taiwan had no locally acquired case in 2021; its rows are included (Table S38; change to the plan in Table S14). Zero weeks are weeks without a reported case, either listed as zero by the source (Brazil 2020–2021 only; 21% of rows) or filled with zero by us. Case definitions: probable cases (Brazil); all reported cases, as labelled by OpenDengue (Colombia); probable and confirmed cases (Peru); and locally acquired confirmed cases by week of onset (Taiwan).

OpenDengue omits most weeks without cases, and outside Brazil nearly all of them (Table S2). Within each country’s coverage window, we therefore set every missing district-week to zero and call these weeks zero-filled. A missing week usually means no case was recorded, but it can hide unreported cases (Section 4.5; Table S56). In samples of Brazilian municipalities, at most 2.1% of filled weeks in a year had at least one probable case recorded in InfoDengue [26,27] (Table S4). In Taiwan, no filled week had a local case recorded in the Taiwan Centers for Disease Control daily file of confirmed cases [28]. In Colombia and Peru, only totals could be checked: district counts summed to within about 5% of national totals in Colombia and to 85–94% in Peru (Supplementary Methods, section A4).

For every model, we excluded from scoring the three districts without a boundary polygon (Table S3), two Peruvian department-weeks flagged as incomplete by a rule fixed beforehand and, from 29 December 2019, Espírito Santo’s 78 municipalities. OpenDengue has almost no rows for this state after 2019, so our zero-filling would create false zeros there while InfoDengue shows cases (Supplementary Methods, section A6). This removed 38,766 of 2,741,949 Brazilian rows (1.41%); the models were not refitted.

### 2.3 Forecasting task

At each origin, a model forecasts the cases h weeks ahead (the horizon). The forecast is a set of nine quantiles of log(1 + cases), the deciles 0.1 to 0.9; these give a median and ranges of likely values. Horizons were 1 to 4 weeks in the future-years split and 1 and 4 weeks elsewhere, as in forecast hubs [29]. A week’s epidemiological year is the calendar year of its Wednesday. For each test year Y, one model per horizon was trained on targets from epidemiological years 2018 to Y − 1, the same rows for every model. Because AlphaEarth begins in 2017 and each embedding is used for the next year, training starts in 2018. The 2020 models therefore learned from two years of data and the 2024 models from six.

### 2.4 Models and inputs

We built a ladder of models (Figure 1A; Table S6). Each model keeps the previous model’s inputs and adds one type of information: recent cases (L1), climate (L2), remote sensing and population (L3, the reference model) and embeddings (L4, the embedding model). The seasonal naive baseline (L0) repeats the cases of the same week a year earlier. The case-history model uses cases in the 12 weeks to the origin, the same weeks in the previous three years and the week of the year. The climate step adds population-weighted ERA5-Land temperature, precipitation and humidity, lagged up to 12 weeks [30]. The remote-sensing and population step adds MODIS vegetation greenness (NDVI) and land-surface temperature for the last three complete months [31,32], and night-time lights, built-up share and log population for the year before the target year [33–35]. The embedding model adds the 64-number AlphaEarth embedding for the year before the target year [10,11]. It enters as a population-weighted district average rescaled to unit length (recomputed independently in Table S5).

The forecasting model was LightGBM quantile regression [36], which combines many small decision trees. We fitted one set of trees per quantile and horizon (learning rate 0.05; a fixed 400 rounds, no early stopping). A random seed sets which rows and inputs each tree samples and how the random controls are drawn. Each fit used all training rows of its split (for example, 1,439,937 for the Brazilian future-years reference model, 2023, horizon 1). Baselines were the seasonal naive forecast, a negative binomial regression and, in Brazil, the pretrained time-series model TimesFM 2.5 without further training [37,38]. Each country used one set of LightGBM settings for every model, control and split, tuned once on the case-history model (train 2018, validate 2019). Within-country models had 127 leaves per tree in Brazil and 31 in Colombia, Peru and Taiwan, with at least 100 rows per leaf. Held-out-country models used Brazil’s settings (127 leaves), or Colombia’s when Brazil was held out. The protocol planned to tune each model separately; we report that as a sensitivity analysis. All runs used about 4,700 core-hours on one 64-thread server.

### 2.5 Location controls

Location controls test whether location information alone could reproduce an apparent benefit of the embeddings. We used six candidate controls (Table S7): five location controls and a 2023 embedding snapshot. We call the control selected for a comparison C*.

- **Average past cases (C1)**: the district’s mean log(1 + cases) before 2018 (2014–2017 in Brazil). A held-out district gets the distance-weighted mean of its five nearest training districts. With a whole country held out, this control cannot be computed.
- **Coordinates (C2)**: latitude, longitude and 80 smooth wave patterns over the globe, which help the model represent broad regional gradients (spherical harmonics up to degree 8) [39].
- **Shuffled embeddings (C3)**: each district receives another district’s AlphaEarth series from the same country, fixed across years (a few keep their own).
- **Smooth spatial noise (C4)**: 64 random smooth maps that vary over the same distance as the embeddings. A short-range version (C4s) varies over the buffer distance of the held-out-regions split.
- **District label (C6)**: the district’s ID as one categorical input, usable only when the test covers future years. It was a candidate only in the analysis without case history; with case history it appears only in the fixed control panel (Table S36).

The 2023 snapshot (C5) was never selected.

With a whole country held out, shuffled embeddings carry no district-level location but, being drawn within each country, still mark the country. They then act mainly as a noise control, showing what 64 extra columns cost.

We chose each comparison’s control on 2019 data. Controls whose 2019 score was within two standard errors of the best were treated as tied, and we took the first tied control in this order: average past cases, shuffled embeddings, coordinates, short-range noise, long-range noise and the snapshot. The rule always picks a control, even when none beats the reference model, because the excess gain needs one to compare against. When that control makes forecasts worse, the excess gain is measured against a weaker model (Section 2.9). A sensitivity rule added after review, the “no control” rule, keeps a control only if it beat the reference model in 2019 by more than two standard errors; otherwise the excess gain equals the embedding gain. In the future-years split we also reran each comparison with each of four controls in turn (a fixed control panel).

### 2.6 Evaluation splits

Five splits define the training and test sets (Figure 1B). The future-years split (S1) tests all districts in later years. The held-out-states split (S2) leaves out whole states (departments in Colombia) in five folds. The held-out-regions split (S3) leaves out compact blocks of neighbouring districts and drops training districts within a buffer distance [40–42]. The buffer (225 km in Brazil, 130 km in Colombia) is the distance beyond which the correlation between two districts’ case counts, with the seasonal pattern removed, fell below 0.1 in 2014– 2019 (Supplementary Methods, section D3). The held-out-districts split (S4) assigns districts at random to five folds within states, and the held-out-country split (S5) trains on three countries and tests on the fourth.

The three within-country spatial splits were run only in Brazil and Colombia, because Peru has too few provinces; with the other two splits in all four countries, this gives 14 comparisons. Held out did not mean unknown to the model. Held-out districts were never used to fit a model, but their own recent case counts were still inputs to their forecasts, so they were only partly unseen.

### 2.7 Information timing

Weekly inputs ended at the origin, monthly inputs used complete months before it, and yearly inputs came from the year before the target’s epidemiological year (Figure 1C). Without a further rule, a forecast made in late December for a January week would use yearly inputs from a year that had not yet ended, and a model trained on cases reported after the forecast date. After review we therefore scored only forecasts whose origin lies in the target’s epidemiological year. This removed both leaks and dropped 4.8% of rows; nothing was refitted.

No test-year outcome of a held-out district entered any fit, but earlier outcomes informed tuning and control selection (Tables S8 to S11).

### 2.8 Scoring

The primary score was the weighted interval score (WIS) [43] on log(1 + cases) [44]. It is a proper scoring rule [45]. It adds the width of each forecast interval to a penalty when the observation falls outside it. Lower is better, and for a single-value forecast it equals the absolute error. For an observation *y* and a central interval from *l* to *u* with coverage 1 − *α*, the interval score and the WIS are

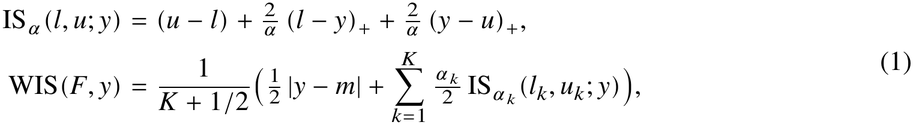

where *F* is the forecast, *y* is the observed log(1 + cases), *m* is the forecast median, and (*x*)_+_ equals *x* when *x* is positive and 0 otherwise. Our nine quantiles give the median plus *K* = 4 central intervals, with bounds *l_k_* and *u_k_* and *α_k_* = 0.8, 0.6, 0.4 and 0.2 for coverage 20%, 40%, 60% and 80%. Our WIS values are therefore not comparable with the 23-quantile WIS of forecast hubs.

Secondary measures were the WIS on the case scale and the coverage of the 40% and 80% intervals. We also split rows into three types of week: weeks with at least one case, zeros listed by the source (Brazil 2020–2021 only) and zero-filled weeks. Because these groups are defined by the observed outcome, they are descriptive only. A forecast that widens its upper tail tends to lose on zero weeks and gain on case weeks whether or not it is better overall (the forecaster’s dilemma [46]). We therefore also report measures computed from the forecasts alone.

### 2.9 Gains

Throughout, a positive gain means that the added input reduced forecast error, and a negative gain, which we call a loss, means that it increased error.

We write *W̅_M_* for the mean WIS of model *M* on the shared rows, with each target week weighted equally (Section 2.10). The embedding gain is how much the embeddings improve the reference model:

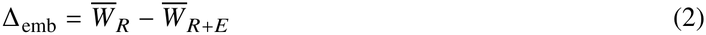

where *R* is the reference model (L3) and *R* + *E* is the same model with the embeddings added (L4). The control gain is how much the selected location control alone improves the reference model:

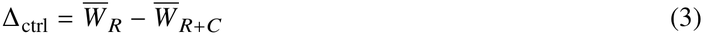

where *R*+*C* is the reference model with the selected control (L3 + C*). The excess gain is what the embeddings add once the model already has the selected control:

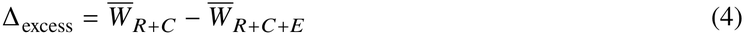

where *R* + *C* + *E* has both the control and the embeddings (L4 + C*). The embedding gain asks whether the embeddings help, the control gain whether location alone helps, and the excess gain whether the embeddings still help once location is known.

The protocol named the excess gain as the primary measure. After review we treated the embedding gain and the excess gain as both main measures, because the selected control sometimes made the reference model worse (Section 3.3). In those comparisons, the excess gain measured the embeddings inside a weakened model. Gains are reported both as absolute differences and as percentages of *W_R_*, which differs widely between countries. The protocol also planned an identity share, Δ_ctrl_/Δ_emb_, for embedding gains whose 95% confidence interval (CI) excluded zero and that were at least 1% of *W_R_*; none qualified.

### 2.10 Statistical analysis

For each target week, we averaged the WIS difference over districts and then over horizons 1 and 4; the estimate is the mean of these weekly values. We compared models with the Diebold–Mariano test [47], which asks whether the average weekly difference in score between two models is larger than chance would produce. Because errors in neighbouring weeks are correlated, we used a variance estimate built for correlated series, the equal-weighted cosine (EWC) estimator [48], and the small-sample correction of Harvey, Leybourne and Newbold [49]. With Holm’s correction for multiple testing [50], this is the main test. Because the test treats each week as one observation, its conclusions apply to these districts, these seasons and the models as fitted. The main test can reject too often when differences drift across seasons (Section 4.5), so we rely most on results that also hold under the two stricter checks below.

Each of the two main measures forms its own family of 14 comparisons for the Holm correction; only the excess-gain family was fixed before the held-out-place results were seen.

To show that two models differ by very little, we used two one-sided tests (TOST) [51]. Equivalent does not mean identical scores, only that the evidence supports a difference smaller than ±1%. A comparison was called equivalent when both one-sided null hypotheses were rejected after Holm adjustment over the 14 comparisons. Without the adjustment, a comparison is equivalent when the 90% CI of its gain lies inside ±1% of the reference model’s WIS. Table 2 marks comparisons that were equivalent before adjustment only. The 1% value was the protocol’s threshold for a material embedding gain; we adopted it as the margin after the Brazil and Peru future-years results had been seen. We report ±0.5% alongside and ±2% as a sensitivity analysis [52]. A loss or gain requires a Holm-adjusted p below 0.05; a loss is small when its estimate lies inside the margin.

**Table 2.** Gains from adding AlphaEarth embeddings, by comparison (positive values mean lower forecast error).

| **Comparison** | **Conclusion (excess gain)** | **Excess gain, % (95% CI)** | **Excess gain, absolute (×10<sup>−4</sup>)** | **Holm-adjusted p** | **Embedding gain, % (95% CI)** | **Selected control** |
|---|---|---|---|---|---|---|
| Brazil, future years | No significant difference; within ±1% | +0.23 (−0.33 to 0.79) | +3.9 | 1.00 | +0.26 (−0.31 to 0.82) | Average past cases (C1)† |
| Brazil, held-out states | No significant difference; within ±1% | 0.00 (−0.34 to 0.34) | 0.0 | 1.00 | −0.02 (−0.34 to 0.30) | Average past cases (C1) |
| Brazil, held-out regions | No significant difference; within ±1% | −0.16 (−0.34 to 0.03) | −2.7 | 0.53 | −0.13 (−0.35 to 0.09) | Average past cases (C1) |
| Brazil, held-out districts | No significant difference; within ±1% | +0.20 (−0.32 to 0.71) | +3.3 | 1.00 | +0.22 (−0.32 to 0.76) | Average past cases (C1) |
| Brazil, held-out country | Small loss, only with the noise control; within ±1% before adjustment only | −0.68 (−0.94 to −0.42) | −13.0 | <0.001 | −0.39 (−1.34 to 0.55) | Shuffled embeddings (C3)* |
| Colombia, future years | Small loss; within ±1% | −0.42 (−0.64 to −0.21) | −5.2 | 0.014 | −0.38 (−0.61 to −0.16) | Average past cases (C1) |
| Colombia, held-out states | Small loss; within ±1% | −0.52 (−0.74 to −0.30) | −6.4 | 0.003 | −0.60 (−0.81 to −0.39) | Average past cases (C1)‡ |
| Colombia, held-out regions | Small loss; not shown within ±1% | −0.68 (−1.08 to −0.29) | −8.5 | 0.030 | −0.82 (−1.14 to −0.49) | Average past cases (C1)‡ |
| Colombia, held-out districts | Small loss; within ±1% | −0.45 (−0.68 to −0.21) | −5.4 | 0.019 | −0.54 (−0.78 to −0.30) | Average past cases (C1)‡ |
| Colombia, held-out country | No significant difference; within ±1% | +0.22 (−0.19 to 0.63) | +2.7 | 1.00 | −0.04 (−0.37 to 0.28) | Coordinates (C2)†‡ |
| Peru, future years | Inconclusive | −0.82 (−1.49 to −0.14) | −19.7 | 0.18 | −0.91 (−1.62 to −0.20) | Average past cases (C1) |
| Peru, held-out country | Inconclusive | +0.93 (−0.16 to 2.02) | +22.0 | 0.53 | +0.68 (−0.59 to 1.96)§ | Shuffled embeddings (C3)*‡ |
| Taiwan, future years | Inconclusive | −0.71 (−1.40 to −0.03) | −2.2 | 0.29 | −0.95 (−1.71 to −0.18) | Average past cases (C1) |
| Taiwan, held-out country | Loss, large relative to a low baseline; not significant under the stricter checks | −18.29 (−29.80 to −6.78) | −36.9 | 0.035 | −21.60 (−31.82 to −11.39)§ | Shuffled embeddings (C3)*‡ |

*Note:* Gains are reductions in WIS on log(1 + cases) as a percentage of the reference model's WIS on the same rows (horizons 1 and 4 pooled; forecasts whose origin lies in the target's epidemiological year; each target week weighted equally). Positive values mean that the embeddings reduced forecast error. The excess gain adds the embeddings to the reference model with the selected control; the embedding gain adds them to the reference model alone. CIs and p values come from the main test (Methods); Holm adjustment is over the 14 comparisons. C1, average past cases; C2, coordinates; C3, shuffled embeddings.
*Seeds:* values average seeds 0 to 2 where three seeds exist (all comparisons except Brazil's held-out states, regions and districts and Colombia held out; Table S52). § Embedding gain from seed 0 only.
*Taiwan:* includes 2021. Without 2021 (seed 0), the excess gain was −0.44% (−1.01 to 0.14) for future years and −11.16% (−15.66 to −6.65) with Taiwan held out. Part of the second change is a smaller denominator (reference WIS 0.0242 without and 0.0202 with 2021; Tables S37 and S38).
*Symbols:* † The selected control beat the reference model in the 2019 selection (Section 2.5). ‡ The control gain's 95% CI lay below zero in the test years (the control made the reference model worse). \* With a whole country held out, shuffled embeddings are a noise control of the same size as the embeddings. They are shuffled within each country, so they still mark the country, but they carry no district-level location.
*Labels:* Small loss, an excess gain below zero with a Holm-adjusted p below 0.05 and an estimate inside ±1%. Within ±1%, equivalence shown by Holm-adjusted two one-sided tests (or before adjustment only, where marked), assuming no drift across seasons. Not shown within ±1%, the 90% CI crosses −1%. Inconclusive, neither a significant difference nor shown equivalence. A comparison can be both a small loss and within ±1%: the loss is statistically clear but smaller than 1%.

Two stricter checks repeat the tests (results in Table S26; their calibration in Table S12). The strict-variance check uses fewer variance terms (B = 4), which is safer when differences drift slowly between seasons. The surrogate-series check calibrates p values on simulated series that keep the pattern of our data (IAAFT surrogates). Further checks treat the reference WIS as an estimate, score on the case scale, or score all rows. A post hoc hidden-case check rescored the stored forecasts after adding cases to some zero-filled test weeks (Table S56). Unless stated, the checks and sensitivity analyses (Table S13) are not adjusted for multiplicity.

### 2.11 Forecasts without case history

An exploratory analysis dropped all 21 case-derived inputs, giving a reference model without case history (N3) and its embedding version (N4). We specified it for Brazil after the future-years results with case history and before scoring any of its own runs. Its main design is the held-out-states split, where knowing which district is which cannot help; the future-years split is a second design (horizons 1 and 4). Its control was chosen from controls without case data (shuffled embeddings, coordinates, district label, short-range and long-range noise, in that order); average past cases entered as a separate step. The model settings are those tuned with case history. After review we added a district-label panel in Brazil’s future years and a repeat in Colombia’s held-out states. We specified each before it produced any result.

### 2.12 Changes to the protocol

The original protocol (15 November 2026) fixed the model ladder, splits, gains and scoring. Table S14 dates all 54 later changes; each was recorded after at least some test results had been seen. The first 23 were recorded before any held-out-place result, and future-years results bore on 11 of them. The rest followed external and internal reviews of earlier drafts, when at least 13 of the 14 comparisons had been seen. Three planned items were not carried out: a long training window, a 23-quantile WIS and per-outbreak reporting for Taiwan.

Seven changes made after results were seen alter the primary numbers (Table S14):

- one set of model settings per country, adopted after Brazil showed that tree size moved the score several times more than the embeddings did;
- the rule for selecting the control;
- pooling horizons 1 and 4 in the future-years comparisons;
- averaging three seeds, first in Brazil’s future years and then wherever extra seeds exist;
- scoring only forecasts whose origin lies in the target’s epidemiological year;
- including Taiwan’s 2021, which moved the seed-0 Taiwan estimates from −11.2% to −17.3% (held out) and from −0.44% to −0.60% (future years);
- excluding Espírito Santo.

Table S43 sets the protocol’s original settings beside the reported results. We did not register the protocol with an outside service; the dated commit history, public from posting, records what was specified and when.

### 2.13 Analyses added in response to review

After a further review, and after the main results had been seen, we specified more analyses before computing them (Tables S43 to S55). From existing forecasts, we added a positive control (does the design detect each step of the future-years ladder?) and checks of the control rule, trends by test year, equivalence, forecast width and embedding stability.

An exploratory information probe asked whether embeddings, coordinates and static covariates (2017– 2018 means of the reference model’s non-case inputs) predict each district’s average weekly log(1 + cases) in 2014–2019. A small LightGBM model was fitted with whole states left out in turn; we report R², the share of between-district variation explained in the left-out states, with CIs from resampling states.

Exploratory new runs, with Holm correction within each family, tested four other forms of the embeddings (Section 3.9). They also added seeds 1 and 2 in nine comparisons, and the three-seed mean became the primary estimate, as decided before any new seed was scored. Table S13 lists the remaining runs.

## 3 Results

### 3.1 Data and baseline performance

In the future-years split, the case-history model had 0.36 to 0.43 times the WIS of the seasonal naive baseline (lower is better). The negative binomial regression and, in Brazil, zero-shot TimesFM did worse (Table S17). The full model, with embeddings and a location control, changed this ratio by at most 0.01.

In the positive control, non-case inputs improved forecasts detectably only in Brazil, where climate, remote sensing and population together gave a gain of +1.09% (95% CI 0.44 to 1.74). In Colombia only remote sensing and population helped detectably, and in Peru and Taiwan no step did (Table S44; unadjusted).

Across all comparisons, the 80% intervals of the full model contained 83% to 99% of observations (Table S20). On weeks with cases, however, they contained only 61% to 75%, and 23% in Taiwan’s future years (Table S20).

### 3.2 Primary results

Adding AlphaEarth embeddings did not significantly improve forecasts in any of the 14 comparisons, by either main measure after Holm correction (Table 2; Figure 2). WIS has no exact translation into case counts, because it also scores how wide the forecast ranges are. As a rough guide only, moving every forecast quantile 0.2% (a factor of 1.002) further from the observed value of 1 + cases would raise WIS by about 0.002. The largest loss within a country (Peru, future years) was about 0.0020.

**Figure 2.**
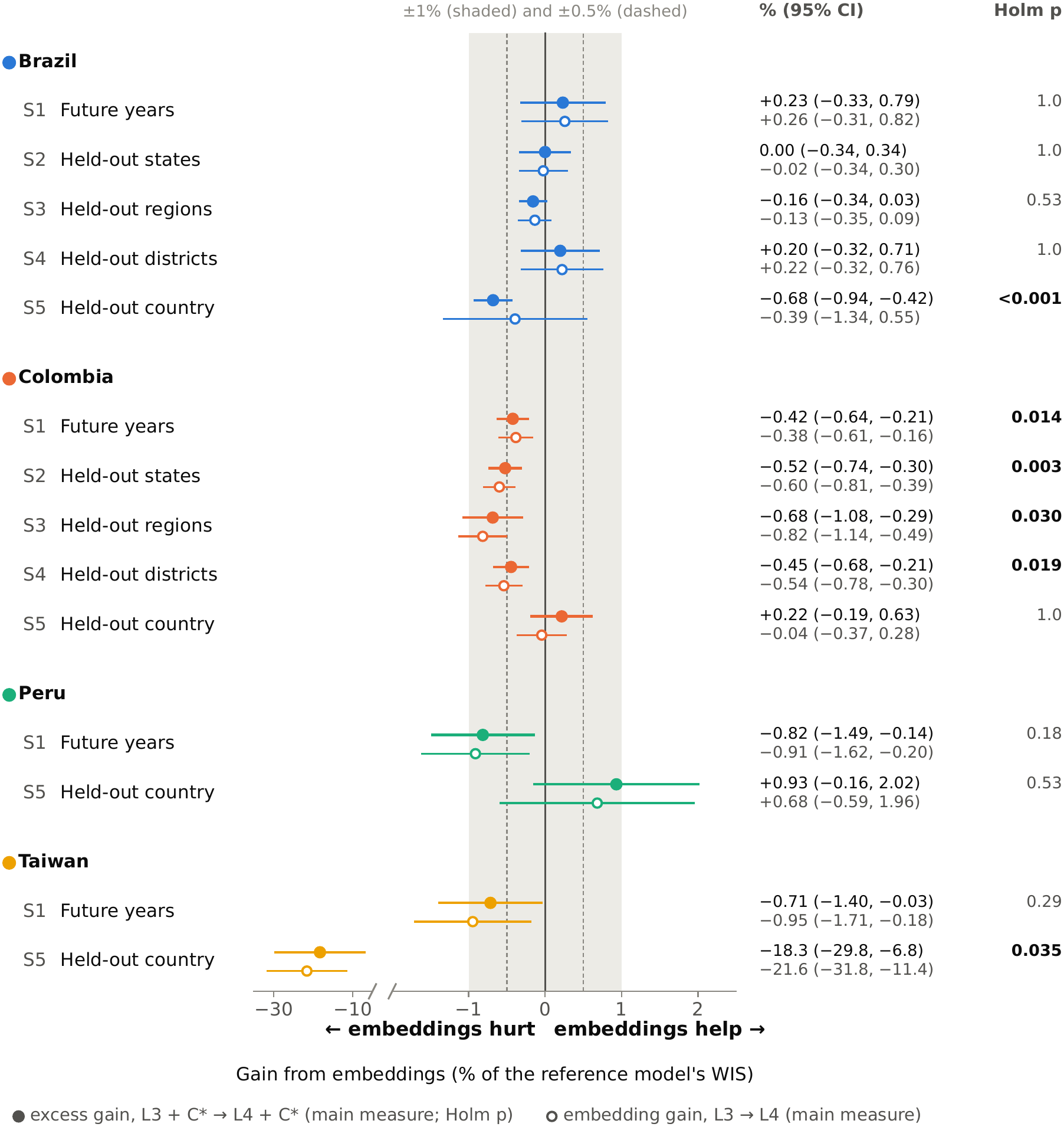
Excess and embedding gains from AlphaEarth embeddings in each comparison. For each country and split, the filled marker is the excess gain (L3 + C* to L4 + C*) and the open marker, just below it, the embedding gain (L3 to L4), both as a percentage of the reference model’s WIS (horizons 1 and 4 pooled; forecasts whose origin lies in the target’s epidemiological year; target weeks weighted equally). Positive values mean that the embeddings reduced forecast error. Lines are 95% CIs from the main test. The shaded band is the ±1% equivalence margin and dashed lines mark ±0.5%. The right-hand columns give each gain with its 95% CI (embedding gain in grey) and the Holm-adjusted p value of the excess gain, over all 14 comparisons (bold: p < 0.05). Taiwan held out is drawn on its own scale after an axis break: excess gain −18.3% (absolute −0.0037) and embedding gain −21.6% (absolute −0.0044). Taiwan includes 2021. S1, future years; S2, held-out states; S3, held-out regions; S4, held-out districts; S5, held-out country.

For the excess gain, six of the 14 comparisons showed a loss after Holm correction, meaning that the embeddings increased forecast error. The most robust loss was in Colombia’s held-out states (−0.52%, 95% CI −0.74 to −0.30; Section 3.4). With Brazil held out, the loss (−0.68%) appeared only when shuffled embeddings were added to both models as a noise control. Without any control, the embedding gain in this comparison was −0.39% (p = 0.39; Table 2). With Taiwan held out, the loss was small in absolute terms (−0.0037) but large as a percentage (−18.29%), because the reference WIS was low (0.0202). It did not stay significant under the stricter checks (Section 3.4).

The largest positive estimate came with Peru held out (+0.93%, 95% CI −0.16 to 2.02; Holm-adjusted p = 0.53). For this comparison, the Holm-adjusted p was 0.050 under the surrogate-series check, too close to 0.05 to call given simulation error, and 0.18 under the strict-variance check.

The embedding gain’s 95% CI lay below zero in seven comparisons: the four Colombian comparisons within the country, Peru and Taiwan in future years, and Taiwan held out (−21.60%, −31.82 to −11.39). In Table 2, the Conclusion column first says whether the excess gain differed significantly from zero. The label “within ±1%” adds that any difference was shown to be smaller than 1%.

### 3.3 Equivalence and the location controls

With the main test, effects larger than ±1% were ruled out in 9 of 14 comparisons before Holm adjustment and in 8 after it, none of them in Peru or Taiwan. With the strict-variance check and Holm adjustment, 5 comparisons were equivalent within ±1% and none within ±0.5% (Table S47). Three of those five, all in Colombia, also showed losses. For them, equivalence means only that the loss was smaller than 1%. Under the surrogate-series check, run without Holm adjustment, every comparison that was equivalent before adjustment stayed equivalent except Brazil’s future years (calibrated 90% CI −0.62% to 1.08%).

The ±1% margin is 0.92 times Brazil’s gain from all non-case inputs, 2.3 times Colombia’s and 5.0 times Peru’s (Table S44).

With case history, the control gain was never significantly positive, and in six comparisons the control made forecasts worse (marked ‡ in Table 2). Such a control can make the excess gain exceed the embedding gain, because the embeddings may partly offset the harm. With Colombia held out, coordinates worsened forecasts by 0.81% (95% CI 0.46 to 1.16), and the excess gain was +0.22% against −0.04% for the embeddings alone. Under the “no control” rule, a control was kept only in the two comparisons where the selected control had beaten the reference model in 2019 (marked † in Table 2). The Colombian losses within the country and the loss with Taiwan held out remained significant, and the loss with Brazil held out did not (Table S45). The selected control was usually average past cases, which adds little once recent cases are known, so in most comparisons the excess gain was close to the embedding gain.

### 3.4 Robustness

Each of the six losses had a 95% CI below zero at horizons 1 and 4 separately, and each was significant before correction under both stricter checks (Tables S25 and S26). After Holm correction, one stayed significant under both checks (Colombia’s held-out states). Three stayed significant under the surrogate-series check only (Brazil held out; Colombia’s future years and held-out regions), and two under neither (Colombia’s held-out districts; Taiwan held out). On the case scale, the excess gain was negative in 13 of 14 comparisons (Table S27).

In every comparison with a loss, each seed was negative on its own, and averaging three seeds gave slightly larger losses than seed 0 alone (Table S52).

Restoring the planned settings (Table S43) left the Brazilian and Colombian conclusions unchanged. Three results were fragile. Peru’s future-years estimate changed sign when each model was tuned separately (−0.82% to +0.65%). The Taiwan future-years loss, significant only before correction, disappeared under most planned settings. With Peru held out, the gain reached p < 0.05 before correction under the strict-variance check, and p = 0.051 on all rows.

### 3.5 Where the losses arise

Figure 3 splits each excess gain by type of week (descriptive; Tables S28 and S29). Zero-filled weeks contributed negatively in all 14 comparisons (95% CI below zero in 12). Weeks with cases contributed positively in 12 of 14 (CI above zero only with Colombia or Peru held out). Zeros listed by the source contributed little. Restricted to district-years with at least one case, the Colombian excess gain stayed negative (−0.29% to −0.33%; p < 0.01 in three comparisons). With Taiwan held out, the loss shrank from −18.3% to −3.3% on district-years with cases.

**Figure 3.**
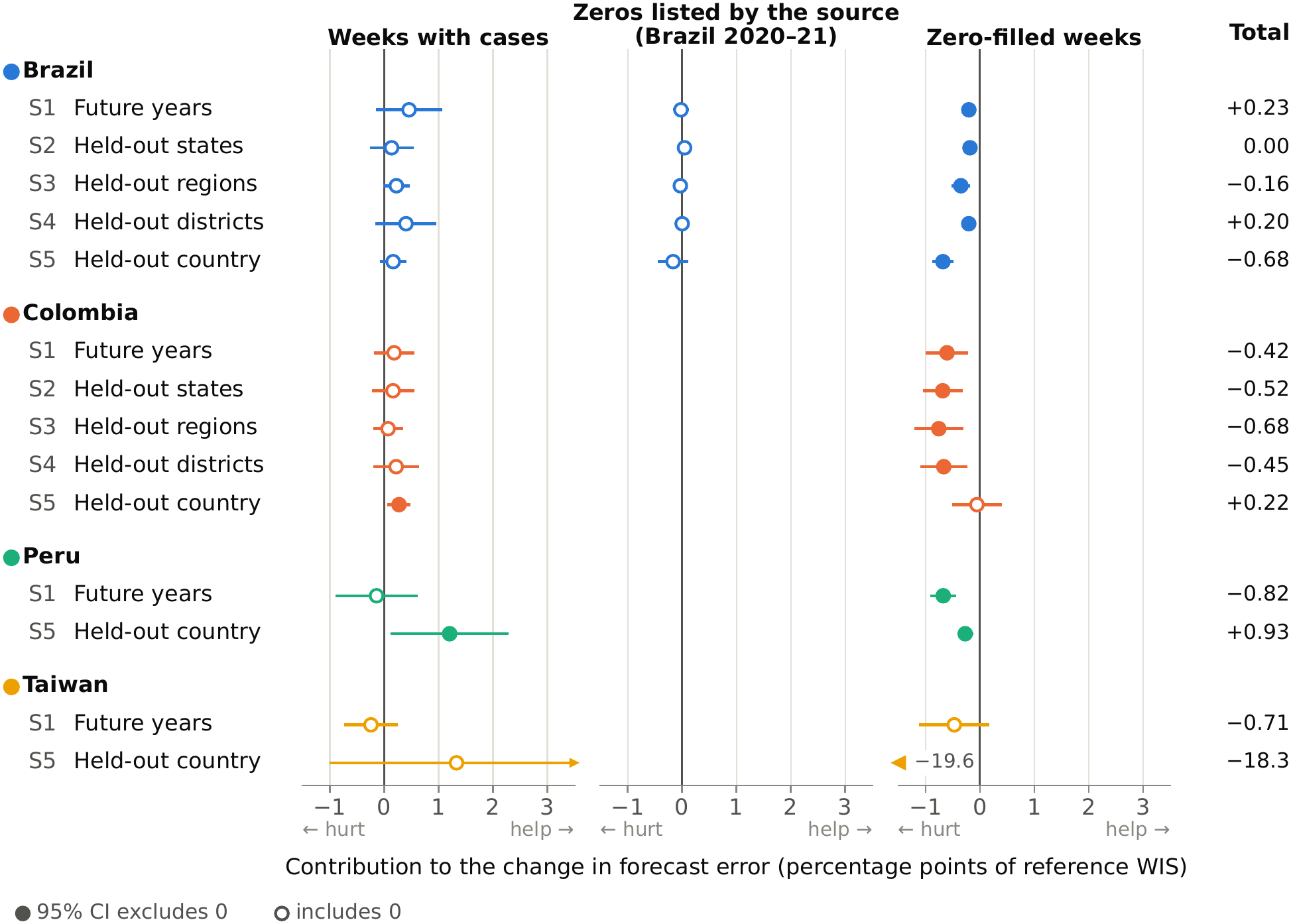
Contributions of three types of target week to the excess gain (descriptive). For each comparison, the excess gain is split additively into contributions from weeks with cases, zeros listed by the source (Brazil 2020–2021 only) and weeks we filled with zero, each in percentage points of the comparison’s reference WIS, with unadjusted 95% CIs (filled marker: CI excludes zero). The three contributions add up exactly to the total printed in the right-hand column. Interval ends beyond the axis are cut with an arrowhead, and an estimate beyond the axis (Taiwan held out, zero-filled weeks) is drawn as an arrow at the panel edge with its value. The strata are defined by the outcome and are descriptive. Taiwan includes 2021. S1 to S5 as in Figure 2.

On weeks without cases, the embeddings raised the average 0.9 quantile, the upper end of the forecast range, in all 14 comparisons (CI excluding zero in 13). Quantiles above the median accounted for at least 63% of the loss in those weeks (Table S31). The embeddings also widened the 80% intervals on zero weeks in all 14 comparisons. Only with Peru held out did they bring the share of case weeks inside these intervals significantly closer to 80% (Table S48).

The hidden-case check (Table S56) assumed that some zero-filled test weeks hid unreported cases. It gave one or three cases to a random share of these weeks, in 20 draws per share, and rescored the stored forecasts; a draw is one random choice of which weeks hide cases. At 2.1% of filled weeks, the highest yearly rate in Brazil’s check (Section 2.2), the six losses shrank by 13% to 22%. After Holm correction, two stayed significant in every draw: the loss with Brazil held out and the loss in Colombia’s held-out states. At 10%, none did, and no comparison became a significant gain.

The Brazilian future-years excess gain was −0.22% in 2020 and −0.67% in 2021, and rose to +0.88% in 2024 (Table S30). With year-to-year variation allowed for, however, no comparison showed a detectable trend in the gain with training length (smallest p = 0.096; Table S46; Figure S2). The shortest training windows fell in COVID-19 pandemic years [53,54], so training length and pandemic-era reporting cannot be separated.

No consistent gain appeared in outbreak weeks, under population or case weighting, after recalibrating the intervals, or in other subgroups (Tables S32 to S35).

### 3.6 Fixed location controls in the future-years split

We reran each future-years comparison with four fixed controls in turn (Table S36; seed 0). None of the 16 excess gains had a 95% CI above zero, and in Colombia, Peru and Taiwan all 12 were negative. In Brazil they were small and positive: +0.02% with coordinates and +0.25% to +0.30% with the other three controls, against an embedding gain of +0.29% (seed 0) without a control. The district label lowered the reference model’s accuracy in all four countries (not significant in Taiwan).

In Brazil, real embeddings did better than shuffled ones of the same size (+0.29% against −0.04%). In Colombia, Peru and Taiwan, shuffled embeddings worsened the reference model by 0.12% to 0.76%, against 0.36% to 1.06% for the real embeddings. We did not test these differences.

### 3.7 Taiwan and transfer between countries

Training on the other three countries instead of within the country improved the reference model for Peru and Taiwan but worsened it for Brazil and Colombia (Table S21).

Measured by cosine similarity to the closest training district’s embedding, Taiwan’s districts were much less similar to the training districts than Brazil’s held-out states were (Tables S39 and S40; Figure S3). All 285 Taiwanese districts had a loss, and more similar districts did not lose less: the most similar fifth lost the most (Spearman rank correlation −0.25).

### 3.8 Forecasts without case history

Without case history (Brazil, future years), the reference model’s error roughly doubled (WIS 0.1675 to 0.3520, target weeks weighted equally; Figure 4B; Tables S41 and S42). The selected control, coordinates, gave a control gain of 3.30% (95% CI 1.68 to 4.93; p < 0.001). The embedding gain without a control was 2.54%, but its CI included zero (−0.82 to 5.89; p = 0.13). With coordinates in the model, the excess gain was −0.43% (−2.06 to 1.19; p = 0.58; minimum detectable effect 2.29%). The district label alone gave a gain of 1.98% (0.76 to 3.21), and the excess gain beyond the label was −0.04% (−1.24 to 1.16).

**Figure 4.**
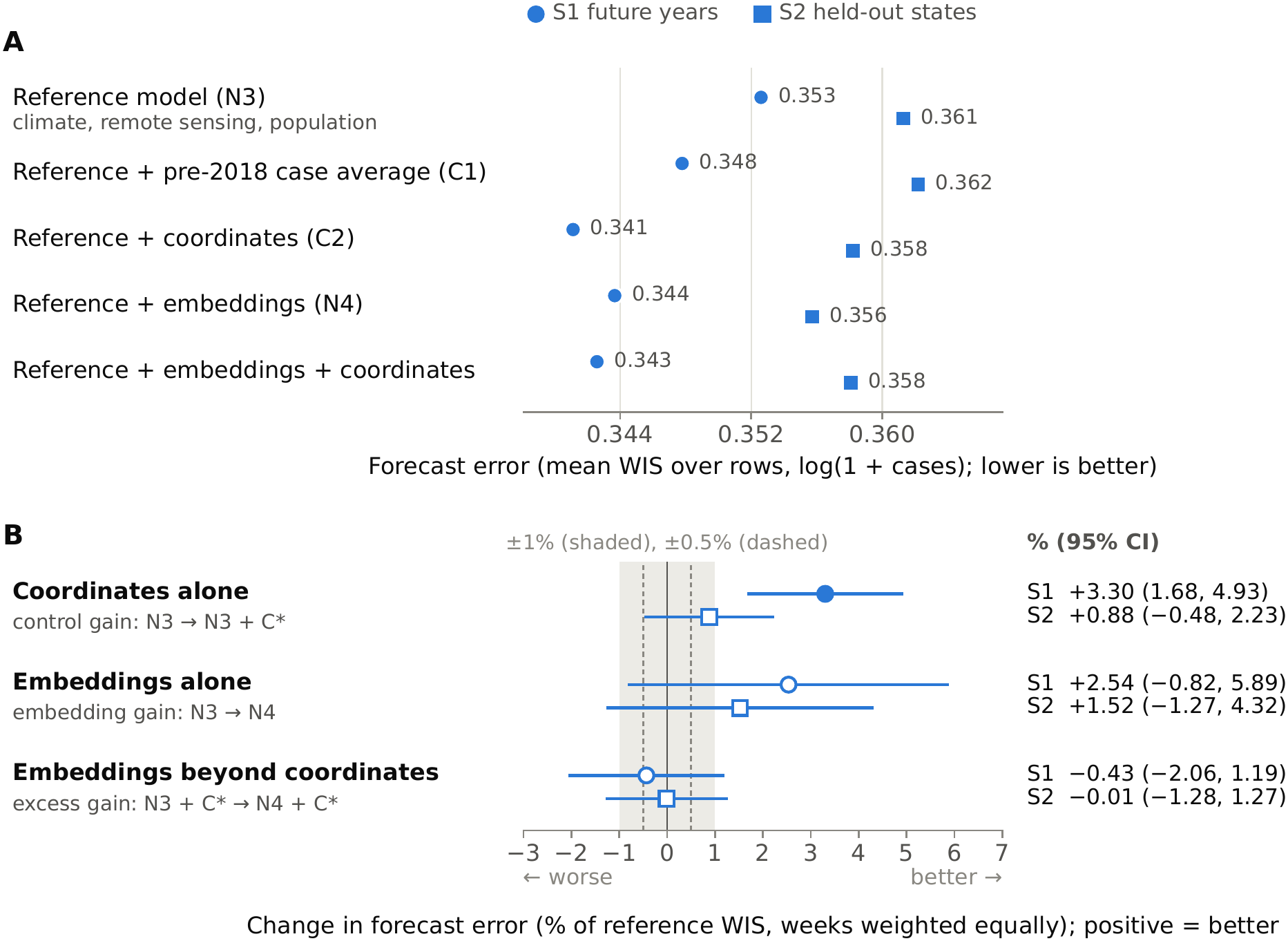
Forecasting without case history (Brazil, future years and held-out states; exploratory). Models use season, climate, remote sensing and population only (reference model N3), plus a location input. (A) Mean WIS over rows for each model, horizons 1 and 4 pooled (seed 0; the same 2,703,183 rows in every model). The selected control is coordinates (C2) in both splits, chosen on 2019 data; average past cases (C1, the district’s pre-2018 case average) is a separate rung. (B) Control gain (N3 to N3 + C2), embedding gain (N3 to N4) and excess gain (N3 + C2 to N4 + C2) as a percentage of the reference WIS. Panel B weights each target week equally and panel A averages over rows, so panel B’s percentages cannot be computed exactly from panel A. Intervals are 95% CIs (filled marker: CI excludes zero). The shaded band is ±1% and dashed lines ±0.5%.

In Brazil’s held-out states, the main design of this analysis, no effect was detected and the intervals were wide (Figure 4; Table S42). With coordinates again selected, the control gain was +0.88% (95% CI −0.48 to 2.23) and the embedding gain +1.52% (−1.27 to 4.32). The excess gain beyond coordinates was −0.01% (−1.28 to 1.27; minimum detectable effect 1.80%). Average past cases, entered as its own step, did not help either. We had specified in advance the pattern that a pure district label would produce: the embeddings would help on their own (a significant embedding gain) but add almost nothing once the model had the district’s location (an excess gain equivalent to zero). It appeared in neither split.

The result in Colombia’s held-out states was different (Table S54). Coordinates were again selected, but here they made forecasts worse (control gain −2.08%, 95% CI −3.47 to −0.69). The embeddings also made forecasts worse, alone (−5.30%, −8.19 to −2.41) and beyond coordinates, as did average past cases.

### 3.9 Analyses added in review

In the exploratory information probe, the embeddings alone explained 24% to 52% of the between-district variation in 2014–2019 burden in held-out states. That is less than the static covariates explained (43% to 66%) but far more than coordinates (R² −0.38 to 0.06; a negative R² means a fit worse than predicting the average). A tree model cannot carry coordinates over to states it has not seen.

Added to coordinates and static covariates, the embeddings raised R² by 0.010 in Brazil (95% CI −0.002 to 0.025) and 0.010 in Colombia (−0.018 to 0.042). Both intervals include zero, as do the wider ones for Peru and Taiwan. Test-year burden gave the same picture, and embeddings predicted burden poorly across countries (Table S50; Figure S4).

No other form of the embeddings gave a significant gain in 28 tests (largest estimate +0.15%; Table S51). Two compact forms, an embedding-similarity version of average past cases and eight principal components, shrank the Colombian future-years embedding loss without reversing it. With a static 2017–2018 embedding, the excess gain stayed negative. Embeddings in place of the remote-sensing and population block did worse than that block in all four comparisons.

In Colombia’s future years, tuning both models separately chose the shared settings, leaving the excess gain unchanged (Table S55).

## 4 Discussion

### 4.1 Principal findings

With case history in the model, AlphaEarth embeddings did not detectably improve weekly district dengue forecasts in any of the 14 comparisons, with or without a location control. Of six losses after correction, only the small one in Colombia’s held-out states held up under both stricter checks and the “no control” rule; the others, including the loss with Taiwan held out (absolute 0.0037), depended on the control or the check.

In unseen states, the embeddings predicted which districts carry a high dengue burden, less well than standard covariates but far better than coordinates; in Brazil and Colombia they added little once both were in the model, even without case history (Section 3.9).

At one and four weeks ahead, recent case counts provided most of the forecasting information, which left little for other inputs, embeddings included, to add. The design detected known covariates only in Brazil and partly in Colombia (Section 3.1). In Peru and Taiwan, where no other non-case input helped detectably, the null results say little about whether the embeddings hold environmental information.

The earlier dengue evaluations of place embeddings that we found used neither location-only controls nor held-out places [18,19]. We applied both across four countries, extending a coordinate control used in one Indian city [22]. Equivalence tests ruled out effects larger than 1% of forecast error in 8 comparisons under the main test, and in 5 of these under both stricter checks.

### 4.2 Why the embeddings can make forecasts worse

Most rows are weeks filled with zero, and the losses concentrate there. In Colombia, however, the loss persisted in district-years with reported cases (Table S48), so it is not only an artefact of filled zeros. We did not analyse how the trees use the embeddings; the following are hypotheses. Annual embeddings do not change within a season, so one possibility is that the model uses them mainly to move its forecasts up or down. They did raise the upper forecast quantiles on zero weeks everywhere (Section 3.5). Such a shift costs accuracy on zero weeks and gains it on case weeks, whether or not it helps overall (Section 2.8).

Part of the loss may come simply from adding many columns. In Colombia, Peru and Taiwan, shuffled embeddings of the same size cost one-third to four-fifths as much as the real ones; in Brazil they cost almost nothing, while the real ones gained slightly (Section 3.6). Compact summaries also shrank the loss (Section 3.9). Sixty-four columns that barely change within a district may give the trees many weak, correlated inputs to choose between, whatever the columns contain. AlphaEarth was trained partly to reconstruct climate reanalysis and elevation [10], which may explain why the embeddings repeat information already in the climate and remote-sensing inputs.

Tree size also matters. In Brazil, tuning each model separately changed WIS by about eight times the embedding gain (Supplementary Methods, section C), so comparisons that tune each input set separately partly measure the tuning. Tuning does not explain the Colombian loss (Section 3.9).

The comparison with Taiwan held out was a stress test. With 98% of its rows zero-filled, the reference error was very low, so a small absolute loss became a large percentage. Its embeddings also lay far from those of the training districts, so the model had to extrapolate. Every Taiwanese district had a loss, whatever its similarity to the training districts [55]. Most of that loss came from district-years without cases.

One possible explanation is that embeddings differ systematically between countries, so a model can use them to learn each country’s case definition and reporting level. Taiwan counts only locally acquired confirmed cases, so a level learned from the other countries would be wrong for every Taiwanese district. This fits the embedding gain better than the excess gain, because both excess-gain models here contain shuffled embeddings, which already mark the country. We did not test embeddings centred within each country, which would block this route. Dengue is also not endemic in Taiwan: imported cases can start local epidemics, and local cases are concentrated in Tainan, Kaohsiung and Pingtung in the south [56]. Environmental similarity to endemic districts therefore says little about local risk.

### 4.3 Forecasts without case history

In these exploratory analyses without case history, coordinates and a district label both reduced error in Brazil’s future years, and the embeddings added nothing detectable beyond either. In that setting, the embeddings may have worked partly as a placeholder, telling the model where a district was or which district it was. In held-out states, where a label cannot help, nothing was detected, but intervals were wide. The labelling pattern we had specified in advance (Section 3.8) appeared in neither split. Other results argue against a pure labelling role. Shuffled embeddings, which identify districts as stably as the real ones, did not help in Brazil’s future years (Table S36) or at the 2019 selection without case history (Table S22). In Brazil, a district’s embedding also changes from year to year about as much as it differs from its neighbours (Table S49). Coordinates also follow climate zones, which makes a placeholder role hard to separate from the environment. These analyses used one seed and settings tuned with case history, and they included no shuffled-embedding run in the test years.

### 4.4 Comparison with other studies

Our results do not contradict the PDFM studies, which used another embedding, monthly targets and a residual design [18,19], but small gains under temporal hold-outs need a location-only control. Like Pradhan [22], we found no detectable embedding advantage, with or without location inputs. In Brazil the point estimate moved from +0.29% without a control to +0.02% with coordinates (seed 0; Table S36), well within the uncertainty. AlphaEarth gains for malaria and respiratory infection [20,21] come from settings without weekly case histories.

### 4.5 Strengths and limitations

The study’s strengths are its four countries, several location controls, a positive control, equivalence tests and a dated log of protocol changes. We could not test PDFM, the embedding used in both earlier dengue studies, because it requires approved access [12]; our results apply to AlphaEarth only.

When the difference between models drifted across seasons, the main test found effects too often. In five comparisons, the surrogate-series check showed that the main test, set at 5%, rejected 14% to 18% of the time (Table S47; earlier simulations in Table S12). In the loss comparisons we checked, the rate was close to the intended level (1% to 9%). The inflated rates weaken the equivalence results in those five comparisons. They do not weaken the finding of no gain: a test that rejects too often finds gains too easily, and the main test still finds none.

Every model used the same LightGBM settings, tuned without embeddings over a small grid. This removes tuning as a confounder but may not suit 64 extra correlated inputs. In Brazil, separate tuning favoured smaller trees, most strongly for the embedding model. Both models were re-tuned only in Colombia’s future years, and the held-out-country settings, Taiwan’s included, were not tuned for pooled data. The embeddings also entered as 64 raw inputs. They are designed for linear and nearest-neighbour use, and because their axes are arbitrary, tree splits may use them inefficiently. A nearest-neighbour risk feature and eight principal components, which suit that design better, also gave no gain (Section 3.9). Other learners, residual designs and learned location encoders [39,57,58] may behave differently.

We used one snapshot of final, revised counts. In practice the latest weeks are incomplete when a forecast is made, so all models would be less accurate in real time, and place inputs might matter more under delayed reporting. Shifting the case inputs back by two weeks in Brazil’s future years gave an embedding gain of +0.60% (95% CI −0.56 to 1.76; Table S37), but this does not reproduce real reporting delays. Test years 2020–2021 fell in the COVID-19 pandemic, when dengue notification declined in parts of Latin America [53,54]. AlphaEarth was trained on imagery that overlaps our test years [10], which, if anything, favours the embeddings.

Filled zeros were checked against independent district-level data only in Brazil and Taiwan, so some in Colombia and Peru may hide unreported cases. If they do, the losses may be overstated: the embedding model gave higher upper forecasts on these weeks (Section 3.5), and these would then score better. The hidden-case check, which changed only the scored weeks and not the models’ inputs or training targets, left only two of the six losses significant after correction in every draw at rates like Brazil’s (Table S56). In Espírito Santo, the false zeros created by our zero-filling were removed from scoring but remain in the training targets.

Because training began in 2018, inputs that are constant within a year had few district-years to learn from in early test years. Held-out districts were only partly unseen (Sections 2.6 and 2.7). Had we tuned tree size within each fold, we would have chosen smaller trees in 12 of 15 Brazilian folds. Removing all case history does not mimic sparse or delayed surveillance.

Our conclusions apply only to the seasons we studied. With Peru or Taiwan held out, only the excess gain was averaged over three random seeds. Some decisions, including the ±1% margin, one set of settings per country and the inclusion of Taiwan’s 2021 season, followed results (Section 2.12). Further limitations are in Supplementary Methods, section I.

### 4.6 Implications and future work

For a dengue programme, the largest loss within a country is, as a rough guide, what forecasts 0.2% further from the observed counts would cost (Section 3.2), whereas case history cut forecast error by about 60% relative to the seasonal naive forecast (Section 3.1). We did not evaluate vector-control or clinical decisions, but differences this small seem unlikely to change them. In the settings we tested (one to four weeks ahead, gradient boosting, four countries), a programme whose weekly forecasts already use recent case counts should not expect much from adding AlphaEarth embeddings. Small gains cannot be ruled out: the upper ends of the 95% CIs reach +0.79% in Brazil’s future years, +0.71% in its held-out districts and +2.02% with Peru held out.

For outbreak preparedness, forecast ranges probably matter more than these small differences. Every model was overconfident in weeks with cases: the full model’s 80% intervals contained only 61% to 75% of those weeks (23% in Taiwan’s future years). A team planning from these ranges should expect counts to fall outside them more often than the 80% label suggests. Embeddings did not fix this, and recalibration helped mainly in Peru (Table S34).

Follow-up work will test PDFM in Brazil, a second model class, embeddings centred within each country, and settings with reporting delays and longer horizons.

### 4.7 Conclusions

Across four countries, AlphaEarth embeddings carried information about where dengue burden is high, but in Brazil and Colombia standard climate, remote-sensing and population inputs already held most of it. With each district’s recent case counts in the model, the embeddings did not detectably improve weekly district dengue forecasts. Under the main test, effects larger than ±1% were ruled out in 8 comparisons, and in 5 of these under both stricter checks as well. That margin is about Brazil’s combined gain from all non-case inputs. With Taiwan held out, the embeddings worsened forecasts, though not robustly. One possible reason is that Taiwan’s case definition differs from that of the training countries. Without case history, our results do not settle whether the embeddings act only as a placeholder: the estimates were imprecise, and the pattern we had specified in advance did not appear (Section 4.3). A gain from foundation-model embeddings is evidence of more than district recognition only if it exceeds the gain from location-only controls in places left out of training. Even then, the extra information need not be environmental. Such a test should use the same settings for every model and fix in advance an equivalence margin, the smallest difference that would matter.

## Supporting information

Supplementary Information (Supplementary Methods and Supplementary Results)

## Data Availability

All input data are public (OpenDengue V1.3.1, Google Earth Engine datasets, IBGE and geoBoundaries boundaries, InfoDengue API, Taiwan CDC open data). The code is available at https://github.com/thesantoshpant/place-or-placeholder. Derived data (weekly district panel, district features, district embeddings, forecasts and scores) are available from the authors on request and will be archived on Zenodo with the journal version.

https://doi.org/10.6084/m9.figshare.24259573.v5

https://data.gov.tw/dataset/21025

https://info.dengue.mat.br/services/api/doc

## Declarations

### Ethics statement

This study used only openly available human data. Weekly dengue case counts aggregated by administrative unit came from the OpenDengue data release V1.3.1 (https://github.com/OpenDengue/master-repo/tree/main/data/releases/V1.3.1; doi:10.6084/m9.figshare.24259573.v5) [24,25]. We also used Taiwan’s public daily file of confirmed cases, which gives counts by onset date and township of residence (“Dengue fever daily confirmed cases since 1998”, Dengue_Daily.csv) [28]. It comes from the Taiwan Centers for Disease Control through the Taiwan government open data platform (https://data.gov.tw/dataset/21025); we used the Internet Archive snapshot of 29 July 2025 (https://web.archive.org/web/20250729004227/https://od.cdc.gov.tw/eic/Dengue_Daily.csv). Weekly municipal dengue counts for Brazil came from the InfoDengue public API (https://info.dengue.mat.br/services/api/doc) [26,27].

The Taiwan CDC file is a public open-government dataset without names or other direct identifiers. We used it only to build and check weekly township totals, and we report only aggregated weekly counts. No new data were collected from people. Ethics approval and consent were not needed, because only openly available, de-identified data were used. The study is not a clinical trial and was not registered.

### Reporting guideline

We followed the EPIFORGE 2020 guidelines for epidemic forecasting and prediction research [23]; the completed checklist is in Supplementary Methods, section G. This is a retrospective forecasting study: all forecasts were produced after the fact from one snapshot of final, revised data.

### Data availability

All input data are public. Dengue cases are from the OpenDengue V1.3.1 Temporal extract (release commit 99b6a3e; figshare version 5), licensed CC BY 4.0 [24,25]. The embeddings are the AlphaEarth Foundations Satellite Embedding dataset, V1 annual (GOOGLE/SATELLITE_EMBEDDING/V1/ANNUAL), from the Google Earth Engine Data Catalog under CC-BY 4.0 [10,11]; this dataset is produced by Google and Google DeepMind.

All covariates were obtained through Google Earth Engine [59]: ERA5-Land daily aggregates (Copernicus licence; contains modified Copernicus Climate Change Service information) [30]; MODIS Terra vegetation indices (MOD13Q1) and land surface temperature (MOD11A2), Collection 6.1 [31,32]; VIIRS annual night-time lights [33]; and the GHSL P2023A population and built-up surface grids [34,35]. Boundaries are the 2022 IBGE municipal mesh for Brazil [60] and geoBoundaries gbOpen for Colombia (DANE MGN 2020, CC BY 4.0), Peru (IGN/OCHA, CC BY 3.0 IGO) and Taiwan (OpenStreetMap 2017, CC BY-SA 2.0) [61,62]. Validation sources were the InfoDengue API (accessed November 2026) [27] and the Taiwan CDC file `Dengue_Daily.csv` (Open Government Data License v1.0) [28].

The following derived data are available from the authors on request and will be archived on Zenodo with the journal version: the weekly district panel with zero-filled weeks; the district features; the population-weighted AlphaEarth embeddings for every district and extracted year from 2017 (64 values each, before and after rescaling to unit length, with the population sums); and all forecasts, scores and result tables. They will be released under CC BY-SA 4.0, as the share-alike terms of the Taiwan boundaries require.

### Code availability

The code is available under the MIT licence at https://github.com/thesantoshpant/place-or-placeholder. It reproduces the whole study, from data download to every table and figure, and comes with unit tests, a locked software environment and manifests that list every data file with its checksum. The public history includes the dated commits cited for pre-specification in the Supplementary Methods (d8c7944, fcc3c38, bd3ba50, e9df1f1).

### Funding

This work received no specific funding. Google Earth Engine was used through its free noncommercial access.

### Competing interests

The authors declare no competing interests.

### Author contributions

All authors contributed equally to the design, analysis and writing, and approved the final version.

### Use of AI tools

The authors designed the study and take full responsibility for its content. Large language model agents (Claude, Anthropic) were used to write and run the analysis code and to help draft and edit the text.

## Acknowledgements

We thank the OpenDengue, InfoDengue and Taiwan Centers for Disease Control teams for making their data public, and Knox College for access to its computing server.

## Preprint licence

This preprint is made available under a CC BY 4.0 licence.

## Notes

### Competing Interest Statement

The authors have declared no competing interest.

### Author Declarations

All data were openly available before the study began. Weekly dengue case counts: OpenDengue V1.3.1 (https://github.com/OpenDengue/master-repo/tree/main/data/releases/V1.3.1; https://doi.org/10.6084/m9.figshare.24259573.v5). Taiwan CDC daily confirmed dengue cases (https://data.gov.tw/dataset/21025; snapshot used: https://web.archive.org/web/20250729004227/https://od.cdc.gov.tw/eic/Dengue_Daily.csv). Brazil validation counts: InfoDengue API (https://info.dengue.mat.br/services/api/doc). Satellite embeddings and environmental covariates: Google Earth Engine Data Catalog (https://developers.google.com/earth-engine/datasets/catalog/GOOGLE_SATELLITE_EMBEDDING_V1_ANNUAL).

