## Supplementary Information (Supplementary Methods and Supplementary Results) for "Place or placeholder? AlphaEarth satellite embeddings did not detectably improve weekly dengue forecasts in four countries"

**References** **64**

### Supplementary Methods

Contents: A. Data sources, processing and verification. B. Features and controls. C. Models and hyperparameters. D. Information available to each model. E. Inference, size simulation, supporting and sensitivity analyses. F. Deviations from the original protocol. G. EPIFORGE 2020 checklist. H. Glossary (Table S16). I. Further limitations.

Automated AI agents (see Use of AI tools) wrote the code and ran the downloads, the verification scripts, the Earth Engine extractions, the model runs, the scoring, the statistical analyses and the internal audits described in this supplement. A check called separate or independent below was written and run by a different automated AI agent from the one that produced the output being checked.

The supplement uses short codes in tables, and the glossary (Supplementary Methods, section H) defines each of them. There are five splits. Future years (S1) forecasts the same districts in later years. The other four hold out states (S2), regions made of compact blocks of districts with a buffer around them (S3), districts assigned at random (S4) and a whole country (S5). One country with one split is a comparison. The models form a ladder of steps, L0 to L4. The reference model (L3) uses case history, climate, remote sensing and population, and the embedding model (L4) adds the satellite embeddings. A location control is an input that tells the model where a district is, which district it is, or how much dengue it usually has, without describing its own environment directly. The six controls are C1 to C6 (Table S7), and C* is the control selected for a comparison.

We report three measures. The embedding gain compares the reference model with the embedding model. The control gain compares the reference model with the reference model plus the selected control; earlier drafts called it the identity gain, and the code and result files still call it `G_id`. The excess gain compares the reference model with the control against the embedding model with the same control. A positive embedding or excess gain means that the embeddings reduced forecast error, and a positive control gain means that the control reduced it. Tables S17 to S56 and Figures S1 to S4 are in the Supplementary Results.

#### A. Data sources, processing and verification

##### A1. Sources and licences

**Table S1. Data sources, versions and licences.**

| Source | Use | Version / product | Licence as recorded |
|---|---|---|---|
| OpenDengue [1,2] | Weekly Admin2 cases | V1.3.1, Temporal extract (release commit 99b6a3e; figshare v5) | CC BY 4.0 (figshare v5) |
| InfoDengue API [3,4] | Brazil zero-fill validation | `alertcity`, column `casprov` (probable) | Public API |
| Taiwan CDC `Dengue_Daily.csv` [5] | Taiwan validation and crosswalk | Internet Archive snapshot of 2025-07-29 | Open government data |
| IBGE municipal mesh [6] | Brazil polygons | 2022 | IBGE open data |
| geoBoundaries gbOpen [7,8] | Colombia, Peru, Taiwan polygons | COL ADM2 (DANE 2020); PER ADM2; TWN ADM2 (OpenStreetMap 2017) | CC BY 4.0; CC BY 3.0 IGO; CC BY-SA 2.0 |
| ERA5-Land daily aggregates [9] | Weekly climate | `ECMWF/ERA5_LAND/DAILY_AGGR` | Copernicus licence |
| MODIS [10,11] | NDVI; land-surface temperature | MOD13Q1 and MOD11A2, collection 6.1 | Provider terms |

*Table S1 (continued)*

| Source | Use | Version / product | Licence as recorded |
|---|---|---|---|
| VIIRS night lights [12] | Night lights | VNL annual V2.1 (2013–2021), V2.2 (2022–2024) | Provider terms |
| GHSL [13,14] | Population, weights, built-up share | P2023A GHS_POP and GHS_BUILT_S, 100 m | Provider terms |
| AlphaEarth Foundations [15,16] | Embeddings | `GOOGLE/SATELLITE_EMBEDDING/V1/ANNUAL`, 64 bands, 10 m | CC BY 4.0 |

All gridded inputs were downloaded from Google Earth Engine [17] and averaged over each district on our own server, weighting each grid cell by the share of it that falls inside the district [18]. Before use, a separate script checked every downloaded file: it compared checksums with the source, tested archive integrity and parsed the whole file. The OpenDengue archives matched the GitHub blob hashes of release commit 99b6a3e and the figshare v5 MD5 sums and parsed without error. The boundary files and the 2024 InfoDengue downloads were checked in the same way.

#### A2. Weekly district panel

We used the weekly, district-level (Admin2) rows of the OpenDengue Temporal extract. All weekly rows are 7-day spans starting on Sunday, and no district appears twice in the same week. The epidemiological year of a week is the calendar year of its Wednesday. Districts are identified by the 7-digit IBGE code in Brazil and by their state and district names elsewhere. The FAO GAUL codes could not be used because they are not unique: Taiwan's 287 districts share two codes.

**Table S2. Weekly district panel after zero-filling.**

| Country | Districts | Weeks | Panel rows | Source weekly rows | Source rows reporting 0 cases | Panel rows zero-filled (whole window) |
|---|---|---|---|---|---|---|
| Brazil | 5,517 | 574 | 3,166,758 | 1,895,538 | 63% | 40.1% (2014–2021: 29.3%; 2022 on: 69.2%) |
| Colombia | 1,071 | 835 | 894,285 | 199,458 | 0.4% | 77.7% |
| Peru | 115 | 1,252 | 143,980 | 30,991 | 0% | 78.5% |
| Taiwan | 287 | 1,409 | 404,383 | 7,673 | 0% | 98.1% |

District counts include only districts that ever reported a case. Peru has 115 of about 196 provinces, and Taiwan 287 of about 368 townships, namely those that ever had a locally acquired case.

Three district counts appear in the paper. The panel has 6,990 districts (Table S2). Of these, 6,987 have a boundary polygon (Table S3; one Colombian and two Taiwanese districts have none). The scored count, used in the main text, is 6,909. The 78 Espírito Santo municipalities have no scored row, because their exclusion (Supplementary Methods, section A6) covers every test year. Brazil therefore contributes 5,439 scored districts, Colombia 1,070, Peru 115 and Taiwan 285.

#### A3. Boundaries

Brazil was joined by IBGE code to the 2022 municipal mesh. Colombia, Peru and Taiwan were matched by name within each state, department or county. Matches were tried in this order: manual fixes, then the Taiwan CDC township list, then exact, normalised and finally approximate (fuzzy) name matches. A fuzzy match needed a rapidfuzz similarity score of at least 85, or 90 in Taiwan, and each district could match only one polygon.

**Table S3. Matching of districts to boundary polygons.**

| Country | Districts matched | Cases 2014 on matched | Match methods |
|---|---|---|---|
| Brazil | 5,517 / 5,517 | 100.000% | IBGE code |
| Colombia | 1,070 / 1,071 | 99.994% | exact 1,005, normalised 30, fuzzy 28, manual 7 |
| Peru | 115 / 115 | 100.000% | exact 113, manual 2 |
| Taiwan | 285 / 287 | 100.000% | CDC crosswalk 271, manual 6, exact 4, normalised 3, fuzzy 1 |

Three districts have no polygon. Belén de Bajirá in Colombia had 40 cases since 2014, and its municipality status was annulled. Taiwan has two “other” categories that are not places and had no cases since 2014. All three are excluded from scoring. The official Taiwan township file returned HTTP 403, so Taiwan polygons are the 2017 OpenStreetMap-derived geoBoundaries file.

*Taiwan crosswalk*

We matched OpenDengue townships to Taiwan CDC townships by choosing, within each county, the pairing that best lines up their weekly case counts. Agreement in weekly counts is therefore partly built in and does not validate the match. A check that does not use counts compared the romanised (pinyin) spelling of every Chinese township name with the OpenDengue name. It agreed for 287 of 287 districts: 281 exactly, 5 by close spelling and 1 after an individual check. In addition, 40 rows of the match were reviewed one by one. Automated AI agents made both checks (see Use of AI tools).

#### A4. Zero-fill validation

*Rule*

The district universe is every district in a country’s weekly file. A country’s coverage runs from its first to its last week with any district row. Within that period, a missing district-week is set to 0 and flagged as zero-filled.

*Brazil vs InfoDengue*

OpenDengue matches InfoDengue's count of probable cases (`casprov`) rather than its count of notified cases.

**Table S4. OpenDengue compared with InfoDengue probable cases in samples of Brazilian municipalities.**

| Year | Municipalities sampled | Present weeks: Σ OpenDengue / Σ probable | Absent weeks with ≥ 1 probable case | Maximum in an absent week |
|---|---|---|---|---|
| 2017 | 40 | 1.008 | 0 of 902 | 0 |
| 2019 | 40 | 1.003 | 0 of 573 | 0 |
| 2022 | 40 | 1.020 | 14 of 1,285 (1.1%) | 4 |
| 2023 | 40 | 0.939 | 26 of 1,231 (2.1%) | 8 |
| 2024 (excluding Espírito Santo) | 58 | 0.964 | 25 of 1,658 (1.5%) | 3 |

The weekly correlation was at least 0.97 in every year and group, and 0.999 in 2024. In weeks present in both sources the differences were small, though larger in later years. This fits revisions to the surveillance data made after the OpenDengue snapshot.

*Taiwan vs Taiwan CDC*

OpenDengue's Taiwan series counts locally acquired confirmed cases by week of onset and township of residence. Imported cases are left out. All 7,673 township-weeks present in OpenDengue match the CDC data exactly, and no absent township-week holds a local case. In 2021 there were no local cases and 12 imported ones. National weekly totals match exactly in 83.7% of weeks. The rest of the difference is 410 early local cases from 1998–2010, 217 of them in 2002, that have no county of residence and so cannot be placed in a township.

*Colombia and Peru*

In Colombia, district sums agree with national totals within about 5%. The weekly ratio of the district sum to the national total is about 1 and stable, with a 10th percentile of at least 0.79 in 2020–2022. This leaves little room for unreported cases in filled weeks. In Peru, district sums cover 85–94% of national totals in 2020–2023. With the same case definition, they are 0.84–0.94 of department totals in 2007–2013. Only 0.4–1.6% of the department-level cases fall in department-weeks in which every district was zero-filled. We could not check absent weeks at province level, so Peru's zero-filling is only partly validated.

#### A5. Completeness rule and exclusions

*Completeness rule*

The rule was fixed after the future-years results but before it was applied to the test years. The primary threshold is k = 0.5, with 0.4 and 0.6 as sensitivity values.

1. Country-weeks (Brazil, Colombia and Peru, in weeks with at least 20 national cases). A week is flagged when the district total is unusually low compared with the national total. Unusually low means that the ratio of the two is below k times its usual value for that time of year and also below k times the median ratio of the same year. The usual value is the median ratio over the previous three years, within ±2 weeks of the same epidemiological week.

2. State-weeks. A week is flagged when its cases fall below k times the smaller of two means: the mean of the two weeks before and the mean of the two weeks after. That smaller mean must be at least 50. If the national series dips in the same way, the flag is used for sensitivity analyses only.
3. Weeks flagged at k ≤ 0.5 by the primary rule are excluded from scoring. All other flags are used only in sensitivity analyses.

*Flags in the test years*

No test week was flagged in Brazil, Colombia or Taiwan. In Peru, two department-weeks were flagged, Junín 2020-12-27 and Ucayali 2023-07-23; they hold 0.03% of rows and 0.02% of cases. In Brazil, 8–22 December 2024 carries a flag used only in sensitivity analyses (1.1% of Brazilian test rows, 0.4% of cases). Taiwan has no national reference, but the state-week check of rule 2 flagged 0 of 96 evaluable weeks. Removing the incomplete-week exclusions restores 16 Peruvian rows and changes no result.

*Gap flag in training*

A country-week is gap-flagged when the national count is at least 20 and the ratio of the district sum to the national total is below 0.25 times that country-year's ratio of sums. Only one week is flagged, Peru on 2018-08-12, and it is dropped from the training targets.

##### A6. Espírito Santo false zeros (D34)

OpenDengue V1.3.1 has no source data for Espírito Santo after 2020. Its district rows end on 2020-12-27, and 63 of the 78 municipalities have their last row on 2019-12-22. The 15 municipalities with 2020 rows report far fewer cases than InfoDengue (7,562 vs 27,077 probable). In 2024 every Espírito Santo week is zero-filled, while InfoDengue shows probable cases in 2,925 of 4,056 of those weeks (72%), 166,407 cases in total. A scan of every state, department and county in the four countries found no other gap of this kind.

*Rule*

Every Espírito Santo municipality, that is, every district whose IBGE code matches `^32\d{5}$`, is excluded from scoring for target weeks from 2019-12-29 (epidemiological year 2020) onward. The exclusion applies to every Brazilian comparison and every model. It removes 38,766 primary rows per comparison, which is 1.41% of the about 2.74 million Brazilian primary rows before the exclusion; 2,703,183 rows remain. It moves every Brazilian excess gain by at most 0.03 percentage points. The rule does not depend on the outcomes of other states, because it follows from the source no longer reporting this state. Models were not refitted, so their 2020–2023 training targets still contain these false zeros. Espírito Santo makes up 1.9–4.5% of Brazil's zero-filled rows in each year.

##### A7. Covariates

*Climate (L2)*

ERA5-Land daily values were summed or averaged over the panel's Sunday-start weeks. The variables are mean, minimum and maximum temperature at 2 m, total precipitation and relative humidity. Humidity was computed from daily mean temperature and dew point with the Magnus formula and capped at 100%. Each grid cell is weighted by GHSL population from the nearest available year and by the share of the cell inside the district, on a 0.01° grid aligned with ERA5-Land's 0.1° grid. Ocean cells get zero weight. We downloaded the weekly grids from Earth Engine and averaged them on our own server. The result agreed with Earth Engine's own district averages to r > 0.999999995 at about 1/20 of the computing cost (Brazil 1.76 EECU-hours). Twenty-three districts, mostly islands, have no ERA5-Land land cell (Brazil 3, Colombia 2, Taiwan 18); they use the nearest land cell.

*Remote sensing and population (L3)*

- NDVI: MOD13Q1 (16-day, 250 m), keeping pixels with quality flag SummaryQA ≤ 1. Values were read from the coarser, about 1 km, copies that Earth Engine stores for large areas. In a pilot this gave r = 0.998 against the native 250 m data at about a quarter of the cost.
- Land-surface temperature (LST): MOD11A2 day and night (8-day, 1 km), without the satellite's quality mask. A version that used the quality flags (v2) did not remove the cold readings caused by clouds, and it changed Brazil's month-to-month pattern (correlation with v1 about 0.85–0.88), so we did not use it. Instead, for Colombia, Peru, Taiwan and every held-out-country model (S5), an LST value more than 10 °C below the district's median for that calendar month over years ≤ 2017 is set to missing (D7). The share of district-months removed by this screen, day and night, was 0.08% and 0.02% in Brazil, 0.06% and 0.57% in Colombia, 0.06% and 0.18% in Peru, and 0.05% and 0.06% in Taiwan. The screen is off for Brazil's within-country splits (S1–S4).
- Month assignment: each composite image is assigned to the month in which it ends, so each month uses only data available by its end. A missing district-month is filled only from the same month of the previous year.
- Night lights: VIIRS annual, from 2013 (missing before). Built-up share: GHSL GHS_BUILT_S, interpolated linearly between the GHSL reference years.
- Population: GHSL GHS_POP, interpolated on a log scale between the GHSL reference years, with no extrapolation beyond them. It exceeds the 2022 Brazilian census by 4–10%, so we use it only as a relative scale.
- Remote-sensing extraction used 8.9 EECU-hours in total.

*AlphaEarth (L4)*

- Image: the annual mosaic of `GOOGLE/SATELLITE_EMBEDDING/V1/ANNUAL` (64 bands A00–A63, 10 m, each pixel vector of unit length), read at a coarser resolution that Earth Engine builds by averaging pixels.
- District mean: a population-weighted mean at 100 m on the GHSL population grid. Each pixel is weighted by its share inside the district times its GHSL population. Negative no-data values were set to 0, and population was interpolated linearly between 2015, 2020 and 2025. Only blocks with people were downloaded. An unweighted 300 m mean is stored but not used in the primary models.
- In a 10-municipality subset of the Brazilian pilot, 100 m gave practically the same means as 10 m (mean cosine 0.99994), while simple resampling to 300 m did not (0.993). Downloading blocks and summing them on our own server matched Earth Engine's own district sums exactly at about 40% of the cost.
- Image years extracted: Brazil 2017–2024, Colombia 2017–2021 and 2023, Peru and Taiwan 2017–2023. Target year y uses image year y − 1.
- District means are shorter than unit length. In the median district, the length of the population-weighted mean is 0.87–0.92 by country (range 0.70–0.98). In Brazil the median is 0.92 and the range 0.74–0.98.

Each mean is rescaled to unit length. Rescaling removes the length of the mean, which is shorter when a district's land cover is more varied; we did not test that length as a separate input.

- The source data have no AlphaEarth pixels for Fernando de Noronha (Brazil) in 2017–2021. Its embedding inputs are therefore missing for target years 2018–2022, and LightGBM treats them as missing values. The missing values are the same in every model that contains the embeddings (L4 and L4 + C*), so they do not differ between the two sides of the excess gain.
- Extracting one state-year again with a different number of parallel workers gave exactly the same sums.

**Table S5. Independent recomputation of AlphaEarth district means.**

| Country | Districts covered | District-years recomputed | Minimum cosine, weighted | Minimum cosine, unweighted | Median cosine, neighbours vs random pairs |
|---|---|---|---|---|---|
| Brazil | 5,517 / 5,517 | 33 | 0.9999999987 | 0.9999999969 | 0.973 vs 0.710 |
| Colombia | 1,070 / 1,071 | 33 | 0.9999999982 | 0.9999999975 | 0.926 vs 0.644 |
| Peru | 115 / 115 | 32 | 0.99999999975 | 0.9999999996 | 0.91 vs 0.44 |
| Taiwan | 285 / 287 | 33 | 0.99999998 | 0.99999989 | 0.975 vs 0.88 |

*Note:* The recomputation used a separate script with its own Earth Engine code; it did not reuse the extraction code. Population sums agree within 0.03%. Districts not covered are those without a polygon. The agreement shows that the extraction can be reproduced. It says nothing about the quality of the embeddings themselves.

#### A8. Computation

LightGBM 4.7.0 ran on CPUs in its deterministic mode with row-wise histograms. Every pair of compared runs used the same number of threads, seed rule and software version. For one model we checked, the predictions were exactly the same with 2, 4, 8 and 16 threads. TimesFM ran on one GPU.

*Training and randomness*

Every model (booster) is trained for a fixed 400 boosting rounds. There is no early stopping and no validation set inside the test-year fits. The seed of a run sets LightGBM's random number generators. These affect the model only through the rows and columns each tree sees: a random 0.8 share of rows at every round and a random 0.9 share of columns. The seed also sets the random draws of two controls, the source map of the shuffled embeddings (C3) and the smooth random fields (C4 and C4s). Nothing else in a run is random. The features, folds, test years and scoring are fixed.

*Size of each fit*

One fit is one model for one quantile, horizon, test year and fold. Each job in the run fits 9 such models, for the median and the 8 other deciles, on the same training rows. Table S5b gives the range of training rows per fit for the full model (L4 + C*) over test years and folds; the smallest is at the first test year, 2020. Within a comparison, all models have the same inputs apart from the embedding and control columns (Table S6). The tuning and selection fits at the 2019 origin trained on the 2018 targets: Brazil 286,884 rows, Colombia 55,692, Peru 5,865 and Taiwan 14,924 (horizon 1).

**Table S5b. Training rows per fit for the full model (L4 + C*), horizons 1 and 4.**

| Comparison | Training rows per fit (range over test years and folds) | Input columns |
|---|---|---|
| Brazil, future years | 573,768 – 1,726,821 | 151 |
| Brazil, held-out states | 456,872 – 1,385,651 | 151 |
| Brazil, held-out regions | 377,624 – 1,290,812 | 151 |
| Brazil, held-out districts | 458,952 – 1,381,582 | 151 |
| Brazil, held-out country | 153,077 – 405,242 | 214 |
| Colombia, future years | 111,384 – 223,839 | 151 |
| Colombia, held-out states | 89,024 – 179,113 | 151 |
| Colombia, held-out regions | 54,912 – 216,942 | 151 |
| Colombia, held-out districts | 89,024 – 179,113 | 151 |
| Colombia, held-out country | 615,461 – 1,236,956 | 232 |
| Peru, future years | 11,845 – 29,900 | 151 |
| Peru, held-out country | 715,000 – 1,794,375 | 214 |
| Taiwan, future years | 29,848 – 89,831 | 151 |
| Taiwan, held-out country | 696,997 – 2,042,232 | 214 |
| Brazil, future years, without case history | 573,768 – 1,726,821 | 211 |
| Brazil, held-out states, without case history | 456,872 – 1,385,651 | 211 |

*Note:* Input columns of the full model (L4 + C*) as recorded during the runs. They are the 150 columns of L4 in Table S6 plus the control: 1 column for C1, 64 for C3 and 82 for C2. Held-out-country fits pool the three other countries.

##### *Total compute*

All LightGBM and negative binomial runs took about 4,700 core-hours, counted as wall time times threads and summed over the 6,180 completed jobs that were stored. This total includes the 2019 tuning and control-selection runs, the sensitivity analyses, the runs added after the third review and some superseded runs kept for the record. It does not include runs discarded after the control fixes (rows 13–15 and 18). The primary and secondary runs with case history took about 2,400 core-hours (3,326 jobs). The analysis without case history took about 900 (804 jobs), the runs added after the third review about 810 (1,722) and the review sensitivity analyses and fixed control panels about 510 (328). TimesFM used 32 minutes of one GPU. Earth Engine extraction used about 182 EECU-hours for AlphaEarth and 8.9 for remote sensing. All runs used one server (AMD EPYC 7551P, 32 cores, 125 GiB RAM).

##### *Derived data to be released*

The district-year AlphaEarth table (population-weighted and unweighted means, before and after renormalisation, with population sums and norms), the weekly district panel, the covariate tables, the split and fold definitions, all stored forecasts and the scoring code are available from the authors on request and will be archived on Zenodo with the journal version; the code is at https://github.com/thesantoshpant/place-or-placeholder. The embedding table is derived from a CC BY 4.0 product; the case panel is derived from OpenDengue (CC BY 4.0), and the Taiwan polygons are CC BY-SA 2.0.

#### B. Features and controls

**Table S6. Inputs added at each step of the model ladder.**

| Level | Block added | Columns | Time alignment |
|---|---|---|---|
| L0 | log(1 + cases) in target week − 52 | 1 | Year before the target |
| L1 | log(1 + cases) at lags 0–11 from the origin (12); target week − 52j and ±1 week, j = 1, 2, 3 (9); sine and cosine of the target's epidemiological week (2) | 23 | Lags end at the origin week |
| L2 | ERA5-Land lags 0–11 of mean temperature, precipitation and humidity (36); 4-, 8- and 12-week rolling means of mean, minimum and maximum temperature, precipitation and humidity (15); population-weighted | 74 | Ends at the origin week |
| L3 | NDVI, day LST and night LST, population-weighted, for the 3 latest complete months (9); night lights and built-up share (2) and log(1 + population) (1) for year y − 1 | 86 | Months ending before the origin month; year before the target's epidemiological year |
| L4 | AlphaEarth population-weighted mean for year y − 1, renormalised (64) | 150 | Year before the target's epidemiological year |
| N3 | L3 without the 21 case-derived columns | 65 | As L3 |
| N4 | L4 without the 21 case-derived columns | 129 | As L4 |

*Note:* Column counts are cumulative and are given for Brazil.

A software check confirms that L0 to L2 contain no district-specific input. Weekly inputs are read through a function that checks that the source week is at or before the forecast origin. Monthly and yearly inputs are read through a function that checks that the source period ends before the allowed limit.

**Table S7. Location controls.**

| Control | Type | Columns | Definition | Where defined |
|---|---|---|---|---|
| C1 | Average past cases | 1 | Mean log(1 + cases) over panel years before 2018 (Brazil 2014–2017), gap-flagged weeks dropped. S1: the district's own value, identical for training and test rows and at every origin. S2–S4: inverse-distance (1/(d + 1 km)) mean of the 5 nearest training districts' values; training districts get a matched leave-out value (S2 excludes their own state, S3 neighbours within D, S4 only themselves) | S1–S4 |
| C2 | Geographic | 82 | Latitude, longitude and 80 orthonormal real spherical harmonics of degree 1–8, no constant term [19,20] | All |
| C3 | Identifier (S1–S4); noise of matching size (S5) | 64 | One fixed source map per seed and country, used in every year: district i gets the real year y − 1 AlphaEarth vector of its source district. S1 and S5: a random permutation of the country's districts; the identity permutation is rejected, but the permutation is not a derangement, so occasionally a district keeps its own vector (about one per country and seed). S2–S4: training districts are permuted among themselves, and each held-out district gets a training district drawn at random (with replacement), never itself | All |
| C4 | Geographic | 64 | 64 Gaussian random fields per country, exponential covariance exp(−d/r) in great-circle distance, standardised within country; r fitted to the AlphaEarth semivariogram (Brazil 8,483 km, Colombia 705 km, Peru 148 km, Taiwan 27 km); drawn only on districts with coordinates | All |
| C4s | Geographic | 64 | C4 with r = min(D, fitted range): Brazil 225 km, Colombia 130 km; equal to C4 in Peru and Taiwan, so not a separate candidate there | Brazil, Colombia, S5 |

*Table S7 (continued)*

| Control | Type | Columns | Definition | Where defined |
|---|---|---|---|---|
| C5 | Snapshot | 64 | AlphaEarth 2023 for every target year | All |
| C6 | District label | 1 | District ID as one LightGBM categorical input | S1 only |

*Note:* Each control takes the place of the AlphaEarth columns (L3 + Ck) or is added to them (L4 + Ck). In the held-out-country split, C3 gives each district the embedding of another district of the same unseen country. It therefore measures the cost of 64 extra columns of realistic values rather than district identity, and we call it a noise control of matching size there. Districts without a polygon or point get missing values for C1, C2, C4 and C4s, which LightGBM treats as missing. These are Belén de Bajirá in Colombia and two "other" categories in Taiwan, and they are excluded from scoring.

Controls are rebuilt for every split, fold, origin and seed. Each district is located by a point guaranteed to lie inside its polygon. We tested that C1 does not change when every row from 2018 on, or every row of a test district, is replaced by an extreme value.

##### B1. Selection of the location control

In plain terms, we keep every control whose selection score is within about two standard errors of the best one, and then take the first of these in a fixed order of preference. In detail, for each comparison every candidate control k is added to the reference model and scored at the 2019 origin: trained on 2018 targets, scored on 2019 targets, at horizons 1 and 4. The score $\mathrm{Score}_k$ is the mean WIS on log(1 + cases) over rows (not week-weighted), averaged over seeds. C3, C4 and C4s use three seeds and the other candidates one. A candidate whose forecasts exactly match the reference model's adds nothing and is dropped. With $\sigma_{\mathrm{seed}}$ the within-candidate seed standard deviation pooled over the multi-seed candidates of the comparison and $n_k$ the number of seeds of candidate k,

$$\mathrm{SE}_k = \frac{\sigma_{\mathrm{seed}}}{\sqrt{n_k}}, \qquad \mathscr{T} = \left\{ k : \mathrm{Score}_k - \mathrm{Score}_{\mathrm{best}} \le 2\sqrt{\mathrm{SE}^2_{\mathrm{best}} + \mathrm{SE}^2_k} \right\},$$

so single-seed candidates (C1, C2, C5, C6) have $\mathrm{SE}_k = \sigma_{\mathrm{seed}}$. C* is the first member of the tie set $\mathscr{T}$ in the priority order C1 > C3 > C2 > C4s > C4 > C5. Without case history the order is C3 > C2 > C6 > C4s > C4. If $\sigma_{\mathrm{seed}}$ is zero or undefined, the comparison is marked uninformative and C* is the first remaining candidate in priority order.

The priority order puts first the controls that most directly encode a district's own dengue history or identity. These are average past cases (C1) and then the shuffled embeddings (C3), which keep the size and distribution of the real embeddings. Next come the smooth geographic controls, from the most flexible (coordinates with harmonics) to the least. The 2023 snapshot comes last because it uses later imagery. The plain reference model was not a candidate, so a control was always selected, even where every candidate scored worse than the reference model at 2019. This happened in 11 of 14 comparisons (Table S22). A rule that allows "no control" is reported as a sensitivity analysis (Supplementary Results, Table S45).

#### C. Models and hyperparameters

*LightGBM*

LightGBM was fitted with a quantile loss on log(1 + cases), with one model (booster) per quantile (9) and horizon. Settings: learning rate 0.05; 400 rounds; feature fraction 0.9; bagging fraction 0.8 at every iteration;

L2 penalty 1; deterministic mode with row-wise histograms. The forecast quantiles were converted back to case counts, set to at least 0 and put in order. A scan of the 77,187,144 stored forecasts found no quantiles out of order and none missing. We did not log how often quantiles were out of order before sorting.

*Tuning*

The search grid was leaves per tree {31, 127} × minimum rows per leaf {100, 1,000}. The criterion was the mean pinball loss, a standard score for quantile forecasts, on log(1 + cases) at quantiles 0.1, 0.5 and 0.9 and horizons 1 and 4. We trained on 2018 and validated on 2019, using level L1 and seed 0. Brazil chose 127/100 (validation 0.07406 vs 0.07428 for 31/100), and Colombia, Peru and Taiwan chose 31/100. One configuration per country is used for every level, control and split. The held-out-country split (S5) uses Brazil's configuration (127 leaves), or Colombia's (31 leaves) when Brazil is held out. Taiwan held out, the comparison with the largest loss, therefore used 127 leaves. The analysis without case history uses Brazil's L1 configuration, which was tuned on a model with case history.

*Per-level tuning (sensitivity)*

Tuning each level separately in Brazil gave 127/100 for L1 and L2 and 31/100 for L3 and L4. The gap between the best and second-best validation loss was 0.29% for L1, 0.03% for L2, 0.06% for L3 and 1.36% for L4. In the Brazilian future-years test years (S1), the reference model with 31 leaves differed from the same model with 127 leaves by about eight times the embedding gain under the shared configuration, pooled over horizons 1 and 4. Per horizon the ratio was 6.5–14 times on the primary rows. This difference motivated one shared configuration (D2). Before the origin-year filter the ratio was about seven.

*Taiwan*

At the 2019 tuning origin Taiwan had 95 non-zero training targets in 33 townships and 56 non-zero validation targets in 26 townships. Its tuning and its choice of control therefore carry almost no information. As a sensitivity analysis, we also ran Taiwan's future-years comparison (S1) with Brazil's configuration and Brazil's selected control.

*Baselines*

Seasonal naive model (L0): the quantile $\tau$ is expm1(log1p(y at t − 52) + the $\tau$-quantile of the training errors log1p(y_t) − log1p(y_{t−52})), set to at least 0. Negative binomial: an NB2 regression on raw counts with the L1 inputs and an intercept, fitted by L-BFGS from a Poisson start, with quantiles taken from the fitted distribution; it was not tuned. TimesFM 2.5 (checkpoint `google/timesfm-2.5-200m-pytorch`) was used as released, without training on our data (zero-shot). It received each Brazilian district's log(1 + cases) history up to the origin, at most 573 weeks, and its deciles were sorted and set to at least 0. It was run for Brazil's future-years comparison (S1) only.

#### D. Information available to each model

"Outcomes" means case counts. "Held-out" means the districts scored in a split's test fold. For test year Y the training targets are epidemiological years 2018 to Y − 1.

**Table S8. Information used at each step of the analysis, by split.**

| Step | Future years (S1) | Held-out states (S2) | Held-out regions (S3) | Held-out districts (S4) | Held-out country (S5) |
|---|---|---|---|---|---|
| Final model fit | All districts, targets 2018 to Y − 1 | Training districts only | Training districts minus the buffer | Training districts | The other three countries |
| Hyperparameters | Country L1 tuning at 2019 (train 2018, validate 2019), all districts | The same S1 tuning, so held-out districts' 2018–2019 outcomes informed the choice | As S2 | As S2 | Brazil's (Colombia's when Brazil is held out); no held-out-country outcome used |
| C* selection | All districts, 2019 outcomes | Train on training districts (2018), score held-out districts' 2019 outcomes | As S2, held-out blocks | As S2, held-out districts | Held-out country's 2019 outcomes |
| C1 value | Own mean log(1 + cases) before 2018 | Held-out: distance-weighted mean of the 5 nearest training districts' pre-2018 means. Training: the same, excluding their own state | As S2; training districts exclude neighbours within D | As S2; training districts exclude only themselves | Not defined |
| C3 value | Fixed random permutation of the country's districts (a district can occasionally keep its own vector) | Held-out district gets a random training district's series (never its own) | As S2 | As S2 | Fixed permutation within the held-out country (a district can occasionally keep its own vector) |
| C4/C4s range | Fitted on AlphaEarth 2017–2018 of all districts (covariates only, no outcomes) | As S1 | As S1 | As S1 | Per country, as S1 |
| Buffer D | – | – | Correlograms of deseasonalised 2014–2019 case series of all districts, held-out included (pre-test; Supplementary Methods, section D3) | – | – |
| Fold construction | – | Admin1 units assigned greedily by district count | k-means on centroids | Random, stratified by Admin1 | – |
| District universe | Districts that ever reported a case, through the end of the data (hindsight) | Same | Same | Same | Same |

*Table S8 (continued)*

| Step | Future years (S1) | Held-out states (S2) | Held-out regions (S3) | Held-out districts (S4) | Held-out country (S5) |
|---|---|---|---|---|---|
| LST screen | Per district and calendar month, median of its own LST over years ≤ 2017 (off in Brazil S1–S4) | Same | Same | Same | Same (on for all countries) |
| Test-time inputs of held-out districts | Own lagged cases, climate, remote sensing, population, AlphaEarth y − 1 | Same (own lagged cases are used) | Same | Same | Same |
| Scoring exclusions | No-polygon districts, incomplete weeks (rule uses ±2 weeks and the same year, i.e. later weeks), Espírito Santo from 2019-12-29 | Same | Same | Same | Same |

##### D1. Where held-out outcomes enter

Outcomes of held-out districts enter in three places, all before the test years. They informed the choice of model settings in S2–S4 (2018–2019 outcomes), the selection of the control in S2–S5 (2019 outcomes) and the buffer distance of the held-out-regions split, S3 (2014–2019 outcomes). The district universe uses outcomes from every year. No outcome of a held-out district from a test year (≥ 2020) enters any fit.

##### D2. Information dated after the forecast origin, removed by the origin-year filter (D24)

Yearly inputs are linked to the target's epidemiological year, and each annual model is trained on labels up to the last week of Y − 1. Rows whose origin falls in Y − 1 could therefore use information from after the origin.

**Table S9. Share of scored rows that could use information from after the origin, around the turn of the year.**

| Horizon | Yearly inputs after the origin (first h target weeks of Y) | Training labels after the origin (first h − 1 weeks) | Maximum weeks of labels after the origin |
|---|---:|---:|---:|
| 1 | 1.91% | 0 | 0 |
| 2 | 3.83% | 1.91% | 1 |
| 3 | 5.74% | 3.83% | 2 |
| 4 | 7.66% | 5.74% | 3 |
| Pooled h 1 and 4 (primary) | 4.79% | 2.87% | |

*Note:* h is the forecast horizon in weeks. The shares are identical in every comparison.

The filter keeps a row only if the epidemiological year of its origin week (the calendar year of the week's Wednesday) equals the test year. On the rows kept, yearly inputs are the same whether they are linked to the target or to the origin, and every training label is dated at or before the origin. In the three within-country Brazilian comparisons the dropped rows slightly favoured the embedding model (L4), so the later information did not cancel out between models.

**Table S10. Information dated after the forecast origin that the origin-year filter does not remove.**

| Item | Why it is later than the origin | Models affected | Direction |
|---|---|---|---|
| Case counts | One snapshot of final revised counts; no reporting delay except the delay-2 sensitivity | All | Unknown; flatters all models |
| ERA5-Land | Reaches Earth Engine about 3 months after real time | L2 and above | Shared |
| GHSL population and weights | Product released 2023; epochs 2015, 2020, 2025 (2025 a projection); climate weights use the nearest epoch, so 2023+ weeks use 2025 | L2 and above | Shared |
| AlphaEarth model | Trained on imagery "covering 2017 to present", production model with labels through 2023–2024; annual layers released 30 July 2025. The imagery period overlaps every test year (2020–2024) | L4, C3, C5 | Could favour the embeddings |
| C5 (2023 static embedding) | 2023 imagery used for 2020–2022 targets | C5 only (never selected) | Favours the control |
| District universe | Districts that ever reported a case; 565 enter in 2018 or later (Brazil 507, 111 first in 2024; Peru 28; Taiwan 18, all in 2023; Colombia 12) | All | Shared |
| Incomplete-week exclusion | Rule uses later weeks of the same year | Scoring only | Shared; test-year effect 2 Peruvian department-weeks |

A sensitivity analysis for late entry, which would restrict the universe at origin Y to districts first seen before Y, is supported by the code but was not run.

##### D3. Buffer geometry of the held-out-regions split

The buffer distance D was estimated on 2014–2019 only, with no test year, from districts with at least 10 weeks with cases (Brazil 4,249; Colombia 806). For each distance band we computed the mean correlation between pairs of districts' weekly log(1 + cases) residuals. We used two kinds of residual. The protocol residuals remove the district mean and week-of-year effects. The second kind also removes each calendar week's national mean. D is the larger of the two distances at which these correlation curves (correlograms) first fall below 0.1. In Brazil the protocol curve levels off at about 0.11 out to 2,000 km. This reflects national co-movement that no buffer can remove, so in Brazil only the second curve drops below that level, giving D = 225 km. In Colombia the two curves drop below it at 130 km and 80 km, so D = 130 km. Every training district within D of any held-out district of the fold is dropped from training.

The buffer is measured from polygon centres (centroids), while C1, C2 and C4 use a point inside each district. Measured between these inside points, 44 Brazilian and 13 Colombian training districts in a fold lie within D of a held-out district; these are about 0.23% and 0.38% of training memberships. The minimum distance between training and test districts in a fold is 161–218 km in Brazil (D = 225 km) and 122–146 km in Colombia (D = 130 km).

##### D4. Tuning within folds

The held-out states, regions and districts splits (S2–S4) reuse the country's future-years tuning (S1), which saw the held-out districts' 2018–2019 outcomes. An automated AI agent repeated the same search within each fold's training districts only, so no held-out or buffer district contributed. Repeating the search over all districts reproduced the stored S1 tuning to every printed digit.

**Table S11. Tuning repeated within each fold's training districts.**

| Country | Split | Folds choosing the S1 configuration | Validation cost of the S1 configuration vs the fold's best |
|---|---|---|---|
| Colombia | S2 | 5 of 5 | 0 |
| Colombia | S3 | 6 of 6 | 0 |
| Colombia | S4 | 5 of 5 | 0 |
| Brazil | S2 | 1 of 5 | 0.17–0.56% |
| Brazil | S3 | 0 of 5 | 0.29–1.05% |
| Brazil | S4 | 2 of 5 | 0.11–0.26% |

In Colombia every fold chose the same settings, so the use of held-out outcomes made no difference. In Brazil, 12 of 15 folds would have chosen 31 leaves. The S1 configuration raised the fold validation loss by 0.11–1.05% (median 0.32%), and in S1 itself the two settings differed by only 0.29%. The Brazilian S2–S4 comparisons were not rerun.

#### E. Inference, size simulation, supporting and sensitivity analyses

##### E1. Primary estimator and test

For a gain and a comparison, let $g$ be the seed-averaged WIS difference of one row. The weekly value $d_{h,t}$ is the mean of $g$ over districts at horizon $h$ and target week $t$, and $d_t$ is the mean of $d_{h,t}$ over the horizons present (1 and 4). The estimate $\bar{d}$ is the mean of $d_t$ over the $T$ test weeks. The main test is a Diebold–Mariano (DM) test. It uses the equal-weighted cosine (EWC) estimate of the long-run variance with $B$ cosine terms and a $t$ reference distribution with $B$ degrees of freedom. $B$ is 16 in Brazil and Taiwan, 11 in Colombia and 13 in Peru. The test also applies the Harvey–Leybourne–Newbold (HLN) small-sample factor at $h = 4$, and Holm correction across the comparisons of the primary family. Ten comparisons average seeds 0 to 2: Brazil S1 and S5, Colombia S1 to S4, Peru S1 and S5, and Taiwan S1 and S5. For Peru and Taiwan held out, the seed average applies only to the excess gain (row 53). The other comparisons use one seed, so their intervals are conditional on one fit. The statistic and its parts are

$$\mathrm{DM} = \frac{\bar{d}}{\mathrm{SE}}, \qquad \mathrm{SE} = \frac{\mathrm{SE}_{\mathrm{EWC}}}{c_{\mathrm{HLN}}},$$
$$B = \lfloor 0.4\, T^{2/3} \rfloor, \quad c_{\mathrm{HLN}} = \sqrt{\frac{T + 1 - 2h + h(h-1)/T}{T}}.$$

The 95% and 90% intervals are $\bar{d} \pm t_B\,\mathrm{SE}$, so a 95% interval excludes zero exactly when $p < 0.05$. The HLN factor at $h = 4$ is applied to the series pooled over horizons 1 and 4. Because the correction grows with $h$, this is the conservative choice. Equivalence is tested with two one-sided tests (TOST). They show equivalence when the 90% interval lies inside ±1% of WIS(L3), where WIS(L3) uses the same week-pooled mean. The minimum detectable effect (MDE), the smallest effect the test could reliably detect, is $(t_{B,0.975} + t_{B,0.80})$ SE without adjustment. Holm-adjusted thresholds are about 1.34 times larger.

The WIS uses the median and the nine deciles, that is K = 4 central intervals (20%, 40%, 60% and 80%) plus the median, normalised by K + ½. It is therefore not on the same scale as the 23-quantile WIS of the forecast hubs (row 36). Unless a table says "row mean", a mean WIS in this paper is week-weighted. That is, we first average over districts and horizons within each target week and then average over weeks.

*Labels and counting*

The label of each comparison in Table 2 combines the main test with the Holm-adjusted TOST. Examples are "Small loss; within ±1%" and "Inconclusive", which means neither a significant difference nor shown equivalence. A loss is an excess gain below zero with Holm-adjusted $p < 0.05$, and a gain is one above zero with the same condition. A loss or gain is called small when it lies inside ±1%. A $p < 0.05$ without Holm adjustment is never counted.

*Margin*

The original protocol used 1% as the smallest embedding gain worth reporting, before any share of it could be attributed to identity (row 9). We decided to use it as an equivalence margin after seeing the Brazil and Peru future-years results (S1). WIS also scores the width of the forecast ranges, so it has no exact translation into case counts. As a rough guide only, moving every forecast quantile roughly 0.2% further from the observed value of 1 + cases would raise WIS by 1% of Brazil's WIS(L3), about 0.0017. Two comparisons give a sense of scale. Within our own study, TimesFM used without training on our data had 13% higher WIS than LightGBM L1 in Brazil's future-years comparison (S1). Their WIS relative to the seasonal naive model (L0) was 0.411 and 0.362. In the US COVID-19 Forecast Hub, neighbouring leading models differed by 0.01–0.02 in relative WIS, about 1.6–3.3% of the leader's WIS [21]. That comparison is on the case scale, while our margin is a share of WIS on log(1 + cases), so it is only a rough guide. Margins of ±0.5% and ±2% are also reported (Table S24).

*Inference population*

Weeks are the sampling unit, for this fixed set of districts, fitted models and test seasons. The folds of the spatial splits divide the districts into groups; they are not random samples. Inference about future seasons would need the variance between seasons. A test at the level of seasons would have only 2–4 degrees of freedom and could barely estimate it.

**E2. Robustness checks**

- The strict-variance check: the main test with only B = 4 cosine terms. With fewer terms the test is less affected by slow drift in the weekly differences, at the cost of power.
- The surrogate-series check. We simulated 2,000 series that have the same distribution of values and the same pattern of ups and downs as the observed weekly differences $d_t$, but a true mean of zero, and asked how often they give a test statistic as large as the observed one. Technically, these are windowed iterative amplitude-adjusted Fourier transform (IAAFT) surrogates [22]. Each is a circular series of length 8T with the real distribution of values and the real spectrum smoothed over a span of 3 (held flat below frequency 1/T), centred at zero, from which a random window of length T is taken. The p value is (1 + k)/(1 + 2,000), where k counts surrogates whose |DM statistic| is at least the observed one.
- Together these are the two stricter checks.
- Delta-method interval for 100 × G / WIS(L3), which allows for the uncertainty in WIS(L3) (EWC + HLN on the linearised weekly series).
- Case scale: the excess gain with WIS on raw counts (expm1 of the log-scale quantiles, set to at least 0).
- All rows: without the origin-year filter.
- Old exclusions: without the Espírito Santo exclusion.

- Resampling of the weekly differences $d_t$ in 52-week blocks (circular block bootstrap). This is descriptive only, because T = 154–256 weeks gives only three to five blocks.

#### E3. Size simulation

The size of a test is how often it finds an effect when there is none; it should match the nominal level. We measured the size of the main test and its alternatives on simulated series with no effect, built to resemble each comparison's own centred weekly differences $d_t$ (2,000 replications per comparison and generating process). We used four ways of generating the series:

- surrogate series with the real pattern of ups and downs, which treats swings over several years as random noise (windowed IAAFT);
- the same with extra slow swings, as a stress test (IAAFT with doubled power below frequency 2/T);
- yearly seasonal cycles plus noise that carries over from week to week, with heavy-tailed residuals resampled from the data (annual harmonics plus AR(1), called ARX);
- a simple Gaussian AR(1) series with $\varphi = 0.5$ and T = 104.

As real placebos we used differences between two seeds of the same model: 21 series from Brazil's future-years comparison (S1; T = 261) and 33 series from the 2019 control runs (T = 52).

**Table S12. Empirical size of the tests at a nominal 5% level.**

| Generating process | Main test (EWC + HLN) | Strict-variance check (EWC + HLN, B = 4) | Newey–West (NW94) | 52-week block bootstrap |
|---|---|---|---|---|
| AR(1), $\varphi = 0.5$, T = 104 | 0.054 | 0.046 | 0.110 | 0.294 |
| ARX, median over comparisons | 0.026 | 0.034 | 0.10 | 0.18–0.21 |
| IAAFT, median over comparisons | 0.048 | 0.036 | 0.20 | 0.18–0.21 |
| IAAFT low-frequency stress, median over comparisons | 0.120 | 0.048 | 0.30 | 0.18–0.21 |
| IAAFT, worst comparisons (BR S1, BR S2, BR S3, CO S5, PE S1) | 0.10–0.22 | ≤ 0.15 | 0.10–0.50 (any process) | 0.15–0.31 (any process) |
| Seed placebo, Brazil S1 (21 series) | 2 of 21 rejected | 1 of 21 | 4 of 21 | 3 of 21 |
| Seed placebo, 2019 controls (33 series, T = 52) | 4 of 33 | 5 of 33 | 13 of 33 | undefined |

*Note:* Size is the share of simulated series with no effect in which the test found one. The Monte Carlo standard error is about 0.5 percentage points. Computed before the origin-year filter and the Espírito Santo exclusion. BR, Brazil; CO, Colombia; PE, Peru.

*Conclusions*

The main test is acceptably calibrated when the weekly differences $d_t$ have short memory around a steady mean, as in most comparisons (size ≤ 8% under ARX). The HLN correction changes the size by at most 1 point. In this earlier simulation, run before the origin-year filter, the test finds effects too often (size 10–22%, up to 28% under the stress test) in comparisons whose gain drifts across seasons, if that drift is treated as noise. These are Brazil S1, S2 and S3, Colombia S5 and Peru S1. Equivalence statements for Brazil S1, Brazil S2 and Colombia S5 therefore hold only if there is no drift across several seasons. The Newey–West and $h - 1$ variance estimators and both block bootstraps found effects far too often.

In this simulation analysis, which came before the origin-year filter and the Taiwan 2021 runs, only Colombia S4 lost Holm significance under the strict-variance check (B = 4; Holm-adjusted p 0.10). On the final primary rows (Table 2 and Table S26; Holm over 14 comparisons), Brazil S5, Colombia S1, S3 and S4 and Taiwan S5 lose Holm significance under the strict-variance check (B = 4), so only Colombia S2 keeps it. Under the surrogate-series check, Colombia S4 and Taiwan S5 lose it. The direction of all six losses is unchanged. In the final surrogate-series calibration of the primary rows (Supplementary Results, Table S47), which replaces the earlier simulation for the final results, the size of the nominal 5% test was 14% to 18% in Brazil S1, S2, S3 and S4 and Colombia S5. Equivalence statements for these comparisons also assume no drift across several seasons. The chance of detecting a gain of 1% of WIS(L3) (the power) is 1.00 in Colombia S5 and about 1.00 in Brazil S2, S3 and S5 and Colombia S1–S4. It is 0.83–0.88 in Brazil S1, Taiwan S1 and Peru S1, 0.53 in Peru S5 and 0.08 in Taiwan S5.

**E4. Supporting analyses (exploratory, no multiplicity adjustment; D33)**

- Baselines in the future-years split (S1): WIS on log(1 + cases) of the seasonal naive model (L0), negative binomial regression, LightGBM L1–L4 and L3/L4 + C*, and TimesFM (Brazil only). WIS relative to L0 is computed on rows scored by every model.
- Additive stratum contributions: for each horizon and week, d = Σ_s w_s d_s, where w_s is the stratum's share of rows. The contributions, as % of the comparison's WIS(L3), therefore add up exactly to the total. We also report each stratum's estimate on its own rows as % of that stratum's own WIS(L3); these are not shares of the total.
- Outbreak weeks: cases ≥ max(5, the district's 90th percentile of weekly cases over 2013–2019).
- Weighted means: within each mean over a horizon and week, districts are weighted by their population in the target year, or by their mean weekly cases over 2013–2017 plus 1.
- Causal recalibration: the forecast ranges are adjusted using only earlier test years, separately for L3 + C* and L4 + C*. Each quantile is shifted by a fixed amount on the log scale, fitted on the earlier test years; the first test year is left unadjusted. There is either one shift per quantile (global) or one shift per group of forecasts with a similar median (called Mondrian).
- Mechanism: the change in the mean 0.9 quantile between L4 + C* and L3 + C* on rows with zero cases, and the share of the loss on those rows that comes from quantiles above the median.
- Embedding similarity (S2–S5): for each held-out district, the cosine similarity between its rescaled year y − 1 embedding and the most similar training district's vector, and the mean over the 5 most similar [23], set against the district's gain.
- Per-year gains with context flags (COVID-19 years 2020–2021, Peru 2023, Brazil 2024).
- Exploratory subgroups (research question 4, where embeddings help or hurt): population tercile in the year before the test year; number of years in 2014–2017 with at least one case (0, 1–2, 3–4); and tercile of the change in GHSL built-up share over 2010–2015.
- Coverage of the 40% and 80% intervals overall and on weeks with cases.

#### E5. Sensitivity analyses

**Table S13. Sensitivity analyses and their status.**

| Analysis | Comparisons | Estimands | Status |
|---|---|---|---|
| Three seeds (primary) vs seed 0 | Brazil S1 | All three | Done |
| Per-level tuning (original protocol) | Brazil S1 | All three | Done |
| Fixed control panel C1, C2, C3, C6 | S1, all countries | Control and excess gains | Done (Table S36; Brazil C3 and C6 also in Table S37) |
| Yearly inputs keyed to the origin year (strictly causal) | Brazil S1 | Embedding gain only | Done (Table S37) |
| Two-week reporting delay in case inputs | Brazil S1 | Embedding gain only | Done (Table S37) |
| Taiwan with Brazil's configuration and C* | Taiwan S1 | All three | Done |
| Taiwan without 2021 | Taiwan S1, S5 | All three | Done |
| Margins ±0.5% and ±2% | All comparisons | Excess gain | Done |
| All rows; old exclusions; case scale; B = 4; IAAFT; delta method | All comparisons | Excess gain | Done |
| Late-entry district universe | All comparisons | Excess gain | Not run |
| Long training window for L0–L3 (2013 to Y − 1; protocol) | Brazil S1 | Ladder | Not run (row 35) |
| 23-quantile WIS (protocol secondary score) | All comparisons | All | Not computed (row 36) |
| As planned vs as reported (Table S43) | All comparisons | Excess and embedding gains | Done (row 44; Table S43) |
| Gains from known predictors along the ladder, as a scale for the margin (Table S44) | S1, all countries | Ladder gains | Done (row 41; Table S44) |
| "No control" selection rule (Table S45) | All comparisons | Embedding or excess gain | Done (row 40; Table S45) |
| Per-year trend test (Table S46) | All comparisons | Excess and embedding gains | Done (row 42; Table S46) |
| Equivalence at ±0.5/1/2% with EWC and B = 4 SEs (Table S47) | All comparisons | Excess gain | Done (row 43; Table S47) |
| Checks that do not select rows by outcome (Table S48) | All comparisons; Colombia district-years | Excess gain, coverage, width | Done (row 45; Table S48) |
| Embedding stability (Table S49) | All countries | Cosine similarity | Done (row 46; Table S49) |
| Information probe (Table S50, Figure S4) | All countries | Cross-validated skill | Done (row 47; Table S50) |
| Representation variants V1–V4 (Table S51) | CO S1, CO S2, PE S1, TW S1, BR S2 | Variant gains | Done (row 48; Table S51) |
| Seeds 0–2 (Table S52) | Colombia, Peru, Taiwan comparisons; Brazil S5 | All three | Done (rows 49 and 53; Table S52) |
| Taiwan future-years ladder and panel with 2021 (Table S53) | Taiwan S1 | Ladder, control panel | Done (row 50; Table S53) |
| No case history, Colombia held-out states (Table S54) | Colombia S2 | All three | Done (row 51; Table S54) |
| Equal-budget tuning (Table S55) | Colombia S1 | Excess gain | Done (row 52; Table S55) |
| District-label panel without case history, N3 + C6, N4 + C6 (exploratory) | Brazil S1 without case history | Control and excess gains | Done (row 38; results below) |
| Hidden-case check: a share of zero-filled test weeks changed to 1 or 3 cases, as if they had hidden unreported cases; stored forecasts rescored (Table S56) | All comparisons | Excess gain | Done (row 54; Table S56) |

The reporting-delay sensitivity analysis only shifts the case-history inputs two weeks back. It does not use real data on reporting delays. The runs with origin-linked yearly inputs and with a delay cover L3 and L4 only, so they are compared with the embedding gain rather than the excess gain. None of the Brazilian sensitivity analyses gave a significant gain. The embedding gain was +0.26% (95% CI −0.29 to 0.82) with strictly causal yearly inputs. It was +0.60% (−0.56 to 1.76) with a two-week reporting delay, against +0.29% for seed 0 of the primary runs. The excess gain was +0.28% (−0.27 to 0.82) with shuffled embeddings (C3) in place of the selected control, and +0.30% (−0.12 to 0.71) with the district label (C6). Row 38 added an exploratory comparison without case history, in Brazil's future-years split (S1) with seed 0. There, the district label (C6) gave a control gain of +1.98% over N3 (95% CI 0.76 to 3.21; $p = 0.003$). This is less than the gain from coordinates (+3.30%). With the district label in the model, the excess gain from the embeddings was −0.04% (−1.24 to 1.16).

#### F. Deviations from the original protocol

We logged each change to the protocol with the date it was recorded and the results that had already been seen. The original protocol is the evaluation plan and its design review of 15 November 2026. The changes were recorded as follows.

- Rows 1–10 were committed in d8c7944. The analysis without case history was pre-specified in bd3ba50. Rows 11–17 were recorded in fcc3c38, and the control fixes of rows 13–15 were implemented in 9de7386. All of this happened on 16 November 2026, before any result of a spatial split had been read.
- Rows 18–23 were committed in e9df1f1 (16 November, 20:29 CST), about three hours before the first spatial-split result (3ca02d7, 23:21 CST).
- Rows 24–34 were committed in f107238 (18 November, 02:16 CST) after external review. By then the S1, S2, S3 and S5 results and Colombia S4 had been seen. Only Brazil S4 and the comparisons without case history were still running.
- Rows 35–38 were added on 18 November 2026 after review of the full draft and recorded in the deposit commit. The Brazil S1 result without case history had been read, but the Brazil S4 result and the Brazil S2 result without case history had not. Rows 35–37 record protocol commitments that were not carried out.
- Rows 39–53 were added on 19 November 2026 after the third review, when all 13 completed comparisons had been read; the result for Brazil's held-out districts had not yet been seen. Each was specified, and recorded in commit 8001a92, before it was scored, and row 53 before any new seed was seen.
- Row 54, the hidden-case check, was added on 22 November 2026, during the authors' review of the full draft, when all 14 comparisons had been read. It rescores stored forecasts against changed targets; nothing was refitted and no primary number changes. Its specification and its results were recorded together in commit 40d7faf (22 November, 19:01 CST).

All 54 rows can be grouped by what had been seen when they were decided. Rows 1–23 (23 rows) were recorded after some future-years test results had been seen but before any held-out-place result. The future-years results for Brazil, Peru and Taiwan bore on the decisions in rows 2, 8, 9, 11, 13, 16 and 18–22 (column "Decided after seeing" of Table S14). The baselines (L0–L2, TimesFM) were also looked at on test years while the analysis code was being developed. Rows 24–34 (11 rows) came after every completed comparison except Brazil's held-out districts and the analyses without case history. Rows 35–38 (4 rows) came after the Brazilian future-years result without case history, rows 39–53 (15 rows) after all 13 completed comparisons, and row 54 after all 14.

Four rows changed the quantity being estimated after results it affects had been read. Row 2 introduced one shared configuration, row 21 pooled horizons 1 and 4 in the future-years comparisons, and row 24 restricted scoring to origin-year rows. Row 25 included Taiwan 2021 and was decided after both Taiwan comparisons had been read. It moved the seed-0 excess gain with Taiwan held out from −11.16% to −17.30%, and the Taiwan future-years gain from −0.44% to −0.60%. With seeds 0 to 2 the final values are −18.29% and −0.71%. Row 53 (seed averaging) changes the primary estimates of the comparisons that gained seeds (Table S52). It changed no conclusion after Holm adjustment over 14 comparisons, although the loss with Brazil held out is no longer significant under the strict-variance check (B = 4) with Holm adjustment (Table S26).

**Table S14. Deviations from the original protocol.**

| # | Change | Reason | Decided after seeing | Commit |
|---|---|---|---|---|
| 1 | C1 under the temporal split = the district's mean log(1 + cases) over the years before the first training target year (Brazil 2014–2017), identical for training and test rows | The first definition (mean of the training targets) leaked each training row's own target: 2019 WIS 0.263 vs 0.175 for plain L3 | 2019 selection-origin results only | d8c7944 |
| 2 | One LightGBM configuration per country, tuned once on L1 at the 2019 origin, for all levels and controls; per-level tuning becomes a sensitivity analysis | Per-level tuning on a nearly flat grid (validation differences 0.03–1.4%) gave L1/L2 127 leaves and L3/L4 31 leaves; on Brazil S1 test years L3 with 127 leaves beat L3 with 31 at every horizon (Holm p ≤ 0.012), a capacity effect several times the embedding gain (about eight times on the primary rows; Supplementary Methods, section C) | Brazil S1 test years | d8c7944 |
| 3 | S2 = grouped leave-states-out in 5 folds (was one state per fold: 27 in Brazil, 33 in Colombia); Brazil's S3 groups 10 blocks into 5 folds; old versions kept in the code, not run | Compute (the 27-fold S2 was 54% of the total); held-out districts are still in states unseen in training | No | d8c7944 |
| 4 | Spatial splits score horizons 1 and 4 only (S1 keeps 1–4); the random district-week split S0 is dropped; final C* runs use 1 seed (selection uses 3 seeds for random controls; the protocol said 10); no re-tuning per split | Compute; Brazil S1 selection seed SD about 0.0001 WIS | No | d8c7944 |
| 5 | C1 in S2/S3: training districts get a matched leave-out value (exclude own state in S2, neighbours within D in S3) | With plain leave-one-out, training districts' neighbours were far closer than held-out districts' (median 23 km vs 105–394 km), so training and test values differed in distribution | No | d8c7944 |
| 6 | S5 configuration = the L1 configuration of the largest training country (Brazil; Colombia when Brazil is held out) | No S5-specific tuning origin exists | No | d8c7944 |

*Table S14 (continued)*

| # | Change | Reason | Decided after seeing | Commit |
|---|---|---|---|---|
| 7 | LST outlier screen for Colombia, Peru and Taiwan (value > 10 °C below the district's calendar-month median from years ≤ 2017 set to missing); off for Brazil S1–S4 | Cloud-contaminated cold MODIS LST (Colombia minimum −42 °C); a QC-bit mask did not remove it | No | d8c7944 |
| 8 | Secondary no-case-history arm added, pre-specified in `docs/methods/nohist_arm.md` before it ran | With case history neither embeddings nor controls helped (Brazil and Peru S1), so identity and place information could not be told apart | Yes (S1 nulls); labelled exploratory | d8c7944; bd3ba50 |
| 9 | Smallest effect of interest = 1% of WIS(L3) (the protocol's ID-share gate); TOST on 90% intervals; minimum-detectable-effect table | A null result needs an explicit margin | Partly (Brazil and Peru S1 seen) | d8c7944 |
| 10 | Deferred: TimesFM + LightGBM residual hybrid, monthly replication arm, PDFM (L5), reporting-lag analysis on data vintages | Time and scope | No | d8c7944 |
| 11 | Inference: DM test on the weekly cross-district mean loss difference with EWC variance, fixed-b t and HLN; interval $\bar{d} \pm t_B \cdot SE$; TOST on the 90% version; the bootstrap becomes a sensitivity analysis with 52-week (not 8-week) blocks | Weekly loss differences stay autocorrelated at lags 8 and 52 (about 0.42–0.43); the 8-week bootstrap excluded 0 where DM p was 0.3–0.5 | Yes (Brazil S1); before any spatial result | fcc3c38 |
| 12 | Primary family = pooled-horizon excess gain, with case history, 14 country × split comparisons, Holm; embedding and control gains, per-horizon and per-year results secondary (since row 39 the embedding gain is also a main measure) | The family had not been fixed | Fixed before any spatial result | fcc3c38 |
| 13 | C3 uses one fixed source map per seed across all years | A new permutation each year meant C3 was not a stable identifier (0–0.05% of districts kept their source) | Found by an automated AI-agent audit (see Use of AI tools) after Brazil S1 test results; C3 had been scored at the 2019 origin only | fcc3c38; code 9de7386 |
| 14 | C1 in S2–S4 = matched neighbour mean of the neighbours' pre-training-year mean log(1 + cases) (was their 2018 to Y − 1 targets) | In-period neighbour targets are contemporaneous with training targets (a diluted leak) and differ from the S1 definition | No spatial result read | fcc3c38; code 9de7386 |
| 15 | C4 at two ranges: the fitted range (Brazil 8,483 km) and a short range (C4s) | At 8,483 km C4 is close to a planar trend, not an identifier | No | fcc3c38; code 9de7386 |
| 16 | Brazil S1 primary runs get 2 extra seeds at horizons 1 and 4; seed SD reported | Per-horizon seed SD (about 0.0001–0.0003 WIS) is of the same order as the excess gain | Yes (Brazil S1) | fcc3c38 |
| 17 | Documented, not changed: yearly inputs keyed to the target year; annual refits train through the end of Y − 1; an origin-keyed sensitivity planned (superseded by row 24) | Automated AI-agent audit | – | fcc3c38 |
| 18 | C4/C4s fields fitted only on districts with coordinates; affected Colombia, Taiwan and S5 runs and the Colombia S1 and Brazil S5 C* runs were discarded and rerun | Three districts without polygons made every field missing in Colombia and Taiwan, so C4/C4s forecasts equalled L3 (Colombia S1 C* had in effect been plain L3) | Yes (Colombia S1 2019 selection, Taiwan S1 test results); no spatial result | e9df1f1 |

*Table S14 (continued)*

| # | Change | Reason | Decided after seeing | Commit |
|---|---|---|---|---|
| 19 | New C* selection rule: drop candidates whose forecasts equal L3's; tie set within 2 seed SEs of the best; first tie-set member in priority C1 > C3 > C2 > C4s > C4 > C5 (without case history: C3 > C2 > C6 > C4s > C4); zero seed SD = "uninformative" | C* was being decided inside seed noise (Colombia margin 0.00003 vs seed SD 0.00006; Taiwan exact ties broken by sort order) | Yes (Brazil, Peru, Taiwan S1); before any spatial result | e9df1f1 |
| 20 | C4s range = min(D, fitted range); in Peru and Taiwan C4s equals C4 and is not a candidate | C4s had run at a longer range than C4 there; it never won, so no C* changed | Yes (Peru, Taiwan S1) | e9df1f1 |
| 21 | The primary family pools horizons 1 and 4 in every comparison; S1 pooled over 1–4 is secondary | The S1 estimand differed from S2–S5 | Yes (S1); before any spatial result | e9df1f1 |
| 22 | Brazil S1 primary comparison uses seeds 0, 1 and 2 at horizons 1 and 4; each estimand uses the seeds common to both its models; seed SD computed where all seeds exist | The manifest listed one seed, and the old fill rule halved the seed SD | Yes (Brazil S1) | e9df1f1 |
| 23 | Scheduler thread budget counts other jobs on the server | Operations | No effect on results | e9df1f1 |
| 24 | Primary scored rows = origin-year rows (drops the first h epidemiological weeks of each test year); all rows reported as a sensitivity | Yearly inputs and annual training labels could postdate the origin (Supplementary Methods, section D) | Yes (after review) | f107238 |
| 25 | Taiwan 2021 is a test year in TW S1 and TW S5; Taiwan S1 explicitly in the family of 14 comparisons; the old test-year set reported as a sensitivity | Excluding a year because it had no cases is an outcome-based exclusion | Yes | f107238 |
| 26 | Three target strata (weeks with cases, zeros listed by the source, zero-filled), each with its own estimate and an exact additive contribution to the comparison total | "Observed" mixed weeks with cases and zeros listed by the source; stratum estimates do not add up under equal-week weighting | Yes | f107238 |
| 27 | Holm-adjusted TOST, plain labels, margin table at ±0.5/1/2%, and a loss-counting rule (Holm-adjusted $p < 0.05$; "small" inside ±1%) | Equivalence claims over 14 comparisons need the same multiplicity control; earlier class words were misleading | Yes | f107238 |
| 28 | Robustness columns: EWC with $B = 4$, IAAFT-calibrated p, delta-method interval, case scale, all rows | The primary test is oversized under multi-season drift in some comparisons; the percentage ignored denominator uncertainty | Yes | f107238 |
| 29 | Fixed control panel (S1: C1, C2, C3, C6 each in place of C*) | The conclusion should not rest on the selected C* alone | Yes | f107238 |
| 30 | WIS on incidence dropped (never computed); case-scale WIS is the secondary scale | The protocol listed a metric that no code computed | Yes | f107238 |
| 31 | The protocol's RQ4 (year-aligned vs static embeddings) deferred; RQ4 in this paper is the exploratory question on where embeddings help or hurt | Time and scope | Yes | f107238 |
| 32 | N2 (climate-only level without case history) not run | Compute; not needed for any estimand without case history | Yes | f107238 |
| 33 | Supporting analyses added on the primary rows (Supplementary Methods, section E), all exploratory | Review | Yes | f107238 |

*Table S14 (continued)*

| # | Change | Reason | Decided after seeing | Commit |
|---|---|---|---|---|
| 34 | Espírito Santo excluded from scoring from 2019-12-29 (Supplementary Methods, section A6); old exclusions reported as a sensitivity | OpenDengue holds false zeros for the state while InfoDengue shows cases | Yes (found in review) | f107238 |
| 35 | Long-window secondary analysis (L0–L3 trained on 2013 to Y − 1) not run | Compute before the preprint; it does not involve the embeddings (AlphaEarth starts in 2017). Limitation: at the 2020 origin every rung trains on two target years | Yes (after review of the full draft) | Deposit commit, 18 November 2026 |
| 36 | 23-quantile WIS (secondary score) not computed; all scores use the median and the 0.1–0.9 deciles | The fitted models output only the median and deciles, so a 23-quantile WIS would need refitting | Yes (after review of the full draft) | Deposit commit, 18 November 2026 |
| 37 | Taiwan per-outbreak and county-level reporting replaced by per-year results (Table S30) and Taiwan's two comparisons in the family of 14 comparisons | Taiwan's test years are almost all zeros, so per-outbreak estimates would rest on a handful of weeks | Yes (after review) | Deposit commit, 18 November 2026 |
| 38 | Exploratory district-label panel without case history: N3 + C6 and N4 + C6 on Brazil S1 test years, seed 0, horizons 1 and 4, primary rows (results in Supplementary Methods, section E5: control gain +1.98%, excess gain −0.04%) | Coordinates (the selected C*) also carry environmental gradients, so a pure identifier is needed to read the result without case history | Yes for other comparisons; no for these runs (added before they produced any result) | Deposit commit, 18 November 2026 |
| 39 | The embedding gain (L3 → L4) becomes a main measure alongside the excess gain and is shown in Table 2 and Figure 2; the "identity gain" is renamed the "control gain" (wording only; code and file names unchanged) | The selected control often scored worse than L3, so the excess gain alone can understate or overstate the embeddings' contribution | Post hoc, after all 13 comparisons were read; specified before scoring | 8001a92 |
| 40 | Sensitivity selection rule allowing "no control": C* = none when no candidate beat L3 at 2019 by more than 2 SE; such comparisons then use the embedding gain (Table S45) | The selection rule always selected a control, even when all candidates were worse than L3 (11 of 13 comparisons) | Post hoc, after all 13 comparisons were read; specified before scoring | 8001a92 |
| 41 | Gains from known predictors along the ladder (L1 → L2, L2 → L3, L1 → L3, L3 → L4) with CIs in every future-years comparison, used to anchor the ±1% margin; ±0.5% reported alongside (Table S44) | The ±1% margin was set after the S1 results and needs an empirical anchor | Post hoc, after all 13 comparisons were read; specified before scoring | 8001a92 |
| 42 | Per-year gains with a weighted trend test on training-window length, and a DM test on 2023–2024 for Brazil (Table S46) | Reviewers asked whether the null result is specific to short training windows | Post hoc, after all 13 comparisons were read; specified before scoring | 8001a92 |
| 43 | Equivalence at ±0.5/1/2% with both the EWC and the B = 4 standard errors (Table S47) | Equivalence claims should also hold under the strict-variance check | Post hoc, after all 13 comparisons were read; specified before scoring | 8001a92 |
| 44 | Supplementary "as planned vs as reported" table (Table S43) | One place to see the effect of every post-result choice on the primary family | Post hoc, after all 13 comparisons were read; specified before scoring | 8001a92 |
| 45 | Checks that do not select rows by outcome: Colombian district-years with at least one case; change in 80% coverage and interval width on case and zero weeks (Table S48) | Outcome-defined strata can reward biased forecasts (forecaster's dilemma) | Post hoc, after all 13 comparisons were read; specified before scoring | 8001a92 |
| 46 | Embedding stability: within-district year-to-year cosine against between-district cosine (Table S49) | Shows how far the yearly embedding acts as a fixed district signature | Post hoc, after all 13 comparisons were read; specified before scoring | 8001a92 |

*Table S14 (continued)*

| # | Change | Reason | Decided after seeing | Commit |
|---|---|---|---|---|
| 47 | Information probe: cross-sectional prediction of district burden from embeddings, coordinates and L3 statics (Table S50, Figure S4) | Separates "no dengue information in the embeddings" from "information redundant with case history at 1–4 weeks" | Post hoc, after all 13 comparisons were read; specified before scoring | 8001a92 |
| 48 | Representation variants V1–V4 (embedding-space risk encoding, PCA-8, causal static 2017–2018 mean, substitution for the L3 block) in five comparisons; Holm within the variant family only (Table S51) | Reviewers asked whether a different use of the embeddings would show a gain | Post hoc, after all 13 comparisons were read; specified before scoring | 8001a92 |
| 49 | Seeds 1–2 for L3, L4, L3 + C* and L4 + C* in Colombia (future years, held-out states, regions and districts), Peru and Taiwan (future years) and Brazil held out, and for L3 + C* and L4 + C* only in Peru and Taiwan held out (Table S52) | Seed SD is of the same order as several estimates | Post hoc, after all 13 comparisons were read; specified before any new seed was seen | 8001a92 |
| 50 | Taiwan future-years ladder (seasonal naive, negative binomial, L1, shared-configuration L2) and fixed control panel rerun with 2021 (Table S53) | Makes all Taiwan tables use the same rows (141,645 rather than 113,430 in some) | Post hoc, after all 13 comparisons were read; specified before scoring | 8001a92 |
| 51 | Analysis without case history repeated for Colombia held-out states, exploratory (Table S54) | Checks whether the Brazilian pattern without case history holds in a second country | Post hoc, after all 13 comparisons were read; specified before scoring | 8001a92 |
| 52 | Equal-budget per-level tuning of L3 + C* and L4 + C* in Colombia S1 (Table S55) | Checks whether one shared configuration favoured or penalised the embedding model | Post hoc, after all 13 comparisons were read; specified before scoring | 8001a92 |
| 53 | A comparison's primary estimate becomes the mean over seeds 0–2 wherever all three seeds exist for both models of an estimand; seed 0 alone becomes a sensitivity analysis (Table S43, Table S52) | Brings every comparison with extra seeds to the estimator already used for Brazil, future years (row 22) | Post hoc, after all 13 comparisons were read; decided 19 November 2026 before any new seed was seen | 8001a92 |
| 54 | Hidden-case check (Table S56), which assumes that some zero-filled weeks hid unreported cases: in each comparison, a share of zero-filled test weeks is changed to 1 or 3 cases, the stored forecasts of both models are rescored, and the main test is rerun; the share at which each loss stops being significant is compared with the 2.1% of filled weeks that held a probable case in Brazil's 2023 check (Table S4) | A co-author asked how sensitive the losses are to cases hidden by zero-filling, since most scored rows are zero-filled and the losses arise there | Post hoc, after all 14 comparisons were read; scoring only, no model refitted | 40d7faf |

##### *Documented, not changed*

(a) The buffer of the held-out-regions split (S3) is measured from district centres (centroids), while C1, C2 and C4 use a point inside each district (Supplementary Methods, section D). (b) The analysis without case history uses Brazil's L1 configuration. (c) Brazil's control selection under per-level tuning ranks the C3 and C4 controls as they were before their later fixes, and it is used only within the per-level sensitivity analysis. (d) Brazilian rows in the held-out-country split (S5) use the LST screen, while Brazil's S1–S4 runs do not. The screen removes 0.08% of daytime and 0.02% of night-time Brazilian district-months. (e) The window for the outbreak threshold, 2013–2019, includes the training years 2018–2019 but no test year.

#### G. EPIFORGE 2020 checklist

Table S15 maps each item of the EPIFORGE 2020 checklist [24] to the manuscript; items are paraphrased.

**Table S15. EPIFORGE 2020 checklist.**

| # | Item | Where | Status |
|---|---|---|---|
| 1 | Identify the study as forecasting research in the title or abstract | Title; Abstract | Present |
| 2 | Purpose and forecast targets | Introduction; Methods 2.3 (target: weekly cases per district, 1–4 weeks ahead) | Present |
| 3 | Methods in reproducible detail, including fitting and assumptions | Methods 2.2–2.12; Supplementary Methods, A–F | Present; code link in item 9 |
| 4 | Prospective, real-time or retrospective | Methods 2.1: retrospective, from one snapshot of final revised data | Present |
| 5 | Origin of input data, with references | Methods 2.2–2.4; Supplementary Methods, A1 | Present |
| 6 | Source data available, or why not | Data availability statement (OpenDengue and all covariate sources are public; derived data available from the authors on request, to be archived on Zenodo with the journal version) | Present |
| 7 | Data processing | Methods 2.2; Supplementary Methods, A2–A7 | Present |
| 8 | Model type and assumptions, with references | Methods 2.4; Supplementary Methods, B and C | Present |
| 9 | Code available, or why not | Code availability statement; protocol, deviation log and code, with the full commit history, at https://github.com/thesantoshpant/place-or-placeholder (no separate pre-results deposit was made) | Present |
| 10 | Validation approach, described and justified | Methods 2.6–2.7; Supplementary Methods, D | Present |
| 11 | Accuracy evaluation, described and justified | Methods 2.8–2.10; Supplementary Methods, E | Present |
| 12 | Comparison with benchmark models, justified | Methods 2.4 (seasonal naive, negative binomial, TimesFM in Brazil); Results 3.1; Tables S17 and S18 | Present |
| 13 | Forecast horizon, described and justified | Methods 2.3: 1–4 weeks, the range used by forecast hubs; horizons 1 and 4 bracket it and are used in S2–S5 to halve compute. Lead times for vector control can be longer; Discussion 4.1 limits the conclusion to one- and four-week forecasts and 4.6 names longer horizons as future work | Present (scope stated) |
| 14 | Forecast uncertainty presented and explained | Quantile forecasts, WIS and interval coverage (Methods 2.8; Results 3.1) | Present |
| 15 | Non-technical summary of results and uncertainty | Abstract; Discussion 4.1 | Partly present (no separate lay summary) |
| 16 | Time-stamped, versioned identifier for forecasts and data | Protocol and code with the full dated commit history at https://github.com/thesantoshpant/place-or-placeholder; forecasts and derived data on request, to be archived on Zenodo with the journal version | Partly present |
| 17 | Weaknesses, including data quality and methods | Discussion 4.5; Supplementary Methods, section I | Present |
| 18 | Implications for public-health action | Discussion 4.6 (scoped: no recommendation about operational use) | Present |
| 19 | Generalisability across populations | Discussion 4.2 and 4.5; four-country design and the held-out-country split | Present |

#### H. Glossary

**Table S16. Glossary of terms and abbreviations.**

| Term | Meaning |
|---|---|
| Admin1, Admin2 | First- and second-level administrative units. Admin1: states (Brazil), departments (Colombia, Peru), counties and cities (Taiwan). Admin2: municipalities (Brazil, Colombia), provinces (Peru), townships (Taiwan) |
| AlphaEarth | AlphaEarth Foundations Satellite Embedding V1: annual 64-dimensional embeddings of satellite imagery at 10 m [15] |
| Area of applicability | The part of feature space where training data are close enough for predictions to be trusted [23] |
| B | Number of cosine terms in the EWC variance; also the degrees of freedom of the t reference |
| Booster, leaves | A booster is one fitted gradient-boosted model, an ensemble of decision trees; here there is one per quantile and horizon. Leaves are the end nodes of each tree, and more leaves allow more complex trees |
| C1–C6, C4s | Location controls (Table S7) |
| C*, selected control | The location control chosen for a comparison at the 2019 origin by the selection rule of Supplementary Methods, section B1 |
| Comparison (cell) | One country with one split. "Cell" is the older name and is used only in some code and file names |
| Case scale | WIS computed on raw counts instead of log(1 + cases) |
| Correlogram | Mean correlation between districts' deseasonalised case series as a function of the distance between them; used to set the S3 buffer D (Supplementary Methods, section D3) |
| Cosine similarity | Similarity of the directions of two vectors, from −1 to 1; 1 means the same direction |
| Coverage | Share of observed values inside a central prediction interval (40% or 80%) |
| D | Buffer distance of the held-out-regions split (Brazil 225 km, Colombia 130 km; Supplementary Methods, section D3) |
| Noise control of matching size (dimension-matched) | C3 in the held-out-country split: 64 columns of real embeddings of other districts of the unseen country, which match the embedding block's size and distribution but carry no district identity the model could have learned |
| Delta method | An approximation for the uncertainty of a ratio (here a gain as a percentage of WIS(L3)) when numerator and denominator are both estimated |
| District | Our word for an Admin2 unit |
| DM test | Diebold–Mariano test of equal forecast accuracy [25] |
| EE, EECU | Google Earth Engine; Earth Engine compute units (a quota meter) |
| Embedding | A fixed-length numeric vector summarising an object; here, a place's satellite observations in one year |
| Embedding gain, $\Delta_{\text{emb}}$ | WIS(L3) − WIS(L4): how much adding the embeddings to the reference model reduced forecast error. One of the two main measures |
| Embedding model | L4: the reference model plus the satellite embeddings |
| Epidemiological week, year | Sunday-start week; its year is the calendar year of its Wednesday |
| EPIFORGE | Reporting guideline for epidemic forecasting studies (2020) |
| EWC | Equal-weighted cosine long-run variance estimator [26] |
| Equivalent within the margin | The interval used for equivalence testing lies inside the equivalence margin, so effects larger than the margin are ruled out (Supplementary Methods, section E) |
| Excess gain, $\Delta_{\text{excess}}$ | WIS(L3 + C*) − WIS(L4 + C*): how much the embeddings reduced error when the model already had the selected location control. One of the two main measures, and the primary measure of the protocol |
| Future years | The split that forecasts the same districts in years after the training years (S1) |
| Fixed-b | Critical values that treat the number of variance terms B as fixed, which keeps tests closer to their nominal size in short series [26] |
| GHSL | Global Human Settlement Layer (population and built-up surfaces) |
| h | Forecast horizon in weeks (target week minus origin week) |
| HLN | Harvey–Leybourne–Newbold small-sample correction [27] |

*Table S16 (continued)*

| Term | Meaning |
|---|---|
| Holm | Holm's step-down multiplicity correction [28] |
| IAAFT | Iterative amplitude-adjusted Fourier transform surrogates: random series with a given distribution of values and a given pattern of ups and downs (spectrum); used in the surrogate-series check |
| Identity confound | The possibility that an embedding improves forecasts because it identifies the district, not because of what it says about the environment |
| Control gain, $\Delta_{ctrl}$ | WIS(L3) − WIS(L3 + C*): how much adding the selected location control to the reference model reduced error. Earlier drafts called it the identity gain, and code and result files call it `G_id` |
| k-means | A clustering method that splits points into k compact groups; used on district centroids to form the S3 blocks |
| L0–L4 | Steps of the model ladder (Table S6) |
| LightGBM | Gradient-boosted decision trees [29]; here with a quantile loss |
| Location control | An input that tells the model where a district is, which district it is, or how much dengue it usually has, without describing its own environment directly (C1–C6). The controls include the district's average past cases, its coordinates, shuffled embeddings, smooth spatial noise and a district label |
| Main test | The Diebold–Mariano test with the EWC variance estimate and the HLN small-sample correction, with Holm correction across the comparisons of the primary family (Supplementary Methods, section E) |
| Location proxy | A feature that improves forecasts mainly by standing in for where a district is |
| LST | Land-surface temperature (MODIS) |
| MDE | Minimum detectable effect at 80% power and 5% two-sided level |
| MODIS | Moderate Resolution Imaging Spectroradiometer |
| Mondrian recalibration | Recalibration with a separate quantile shift for each bin of the forecast median |
| N3, N4 | Levels without case history: L3 and L4 without case-derived inputs |
| NB | Negative binomial regression |
| NDVI | Normalised difference vegetation index |
| Origin | The last week whose data a forecast may use; forecasts are made at its end |
| Origin-year rows | Scored rows whose origin lies in the same epidemiological year as the target (primary) |
| Outbreak week | A district-week with cases at least max(5, the district's 90th percentile of weekly cases over 2013–2019) |
| q0.9 | The 0.9 quantile of a forecast |
| Quantile | The value below which a given share of the forecast distribution lies; the 0.9 quantile is exceeded with probability 0.1 |
| Zero listed by the source | A week the source lists with 0 cases (Brazil 2020–2021 in the test years) |
| Reference model | L3: the model with case history, climate, remote sensing and population, but without embeddings |
| Strict-variance check | The main test repeated with fewer variance terms (a smaller B), which is less affected by slow drift (Supplementary Methods, section E) |
| Surrogate-series check | P values calibrated on simulated IAAFT series that share the data's distribution and pattern of ups and downs but have no true effect (Supplementary Methods, section E) |
| Row mean | A mean WIS over all scored rows, so weeks with more rows weigh more; used only where a table says so |
| S1–S5 | Evaluation splits: future years (S1), the same districts forecast in later years; held-out states (S2); held-out regions, compact blocks of districts with a buffer around them (S3); held-out districts, assigned at random (S4); and held-out country (S5) |
| SE | Standard error |
| Static history | The historical-risk control (C1) used as a separate step in the analysis without case history |
| Two stricter checks | The strict-variance check and the surrogate-series check together |
| With a location control | A model with the selected control added (+ C*), as in L3 + C* and L4 + C* |
| Spherical harmonics | Smooth functions of position on the globe, used as flexible coordinate features (C2) [19] |
| T | Number of target weeks in a comparison's weekly series |

*Table S16 (continued)*

| Term | Meaning |
|---|---|
| TimesFM | A pretrained time-series foundation model, used zero-shot as a baseline [30] |
| TOST | Two one-sided tests for equivalence [31] |
| VIIRS | Visible Infrared Imaging Radiometer Suite (night lights) |
| WIS | Weighted interval score [32]; lower is better. Here from the median and nine deciles (K = 4 intervals), normalised by K + ½ |
| Week-weighted | A mean over target weeks of the within-week mean over districts and horizons, so each week has equal weight (the default in this paper) |
| WIS(L3), reference WIS | Mean WIS of the reference model in the comparison; the denominator of every percentage |
| y − 1 | The year before the target year, the source year of yearly inputs |
| y, Y | y: the target's epidemiological year; Y: the test year of an annual refit |
| Zero-filled week | A district-week missing from the source and set to 0 |
| Zero-shot | Used as released, without training or fine-tuning on our data (TimesFM) |

#### I. Further limitations

- Calibration. The 80% intervals of the full model (L4 + C*) covered 61–75% of weeks with cases. In Taiwan's future-years comparison they covered 23%, or 24% without 2021. The other steps of the ladder were similar (56–75%). Adjusting the forecast ranges with earlier test years (causal recalibration) raised coverage mainly in Peru.
- Countries. Peru (115 provinces) was not shown to be equivalent within ±1%. The smallest effect the test could reliably detect was 0.95% for future years and 1.53% with Peru held out. Taiwan's data are almost all zeros, with 95 non-zero training targets at the 2019 tuning origin, so its tuning and control selection carry almost no information. Case definitions differ: probable cases in Brazil, all reported cases as labelled by OpenDengue ("Total") in Colombia, probable and confirmed cases in Peru, and locally acquired confirmed cases by week of onset in Taiwan. Percentages therefore compare models within a country.
- Covariates and geometry. The land-surface-temperature screen was off in Brazil's within-country runs. Population grids exceed Brazil's 2022 census by 4–10%. The buffer of the held-out-regions split is measured from district centres, and 0.2–0.4% of training districts in a fold fall inside it when distances are measured from points inside each district. The quality exclusions use neighbouring weeks, though dropping them changes no result.
- Information dated after the forecast. The AlphaEarth annual layers were released on 30 July 2025 by a model trained on data that extend into our test years [15]. Population is interpolated towards a 2025 projection. The district universe, districts that ever reported a case, uses hindsight: 565 districts first reported in 2018 or later, including 111 Brazilian municipalities first in 2024. Supplementary Methods, section D, lists every such item.
- Filled zeros. In Brazil in 2024, 1.5% of absent weeks held one to three probable cases, with Espírito Santo excluded. Taiwan's township match was inferred from counts, so agreement in counts does not validate it; a check of names matched all 287 townships (Supplementary Methods, section A3).
- Inference. The earlier size simulations were computed before the origin-year filter and the Espírito Santo exclusion. In them, the main test wrongly found an effect in 10–22% of simulated series with

no real effect, against a nominal 5%, in the comparisons with drift across several seasons. The rate reached 28% under a stress test and 10–12% in seed placebos (Supplementary Methods, section E3). On the final rows, the surrogate-series calibration gave a size of 14% to 18% in five comparisons: Brazil's future years, held-out states, held-out regions and held-out districts, and Colombia held out (Supplementary Results, Table S47). On the current rows, with Holm correction, the two stricter checks give the following picture (Table S26). The loss in Colombia's held-out states stays significant under both checks. The losses with Brazil held out and in Colombia's future years and held-out regions stay significant under one of them, the surrogate-series check. The losses in Colombia's held-out districts and with Taiwan held out stay significant under neither.

### Supplementary Results

Most WIS values in this supplement are week-weighted: we average over districts and horizons within each target week, then average over weeks. Tables marked "row mean" instead average over all rows, so their values differ slightly from the week-weighted ones (details below). Unless a table states otherwise, the following conventions apply.

- Score: the weighted interval score (WIS) on log(1 + cases), computed from the median and the deciles 0.1 to 0.9.
- Rows: one row is one district, target week and horizon, with horizons 1 and 4 pooled. Only origin-year rows are scored, that is, forecasts whose origin lies in the target's epidemiological year.
- Exclusions: districts without a boundary polygon, two incomplete Peruvian department-weeks, and Espírito Santo from 29 December 2019. Taiwan includes test year 2021 unless a row or table says "without 2021".
- Gains: percentages of the reference model's WIS on the same rows (L3, or N3 without case history). A positive value means that the added input reduced forecast error.
- Tests: confidence intervals (CIs) and p values come from the main test, a Diebold–Mariano (DM) test on the weekly mean loss difference with the equal-weighted cosine (EWC) variance and the Harvey–Leybourne–Newbold (HLN) correction. p values are unadjusted unless marked Holm.
- Rounding: percentages have two decimals, p values two significant digits (smallest shown <0.001) and WIS four decimals. A value that rounds to zero is written 0.00, without a sign. An interval bound that rounds to zero is given to one more decimal so that its side of zero stays visible (for example −0.003).

The codes are defined in the glossary of the Supplementary Methods. Model codes: L0, seasonal naive; L1, case history; L2, adds weekly climate; L3, the reference model, adds remote sensing and population; L4, the embedding model, adds the AlphaEarth embeddings; C*, the selected location control; N3 and N4, L3 and L4 without any case-derived input. Location controls: C1, average past cases; C2, coordinates; C3, shuffled embeddings, which in the held-out-country split act as a noise control of matching size rather than a location control; C4 and C4s, smooth random fields at the fitted range and at a short range; C5, the 2023 embedding snapshot; C6, a district label. Splits: S1, future years; S2, held-out states; S3, held-out regions; S4, held-out districts; S5, held-out country. A comparison is one country with one split. The embedding gain compares L3 with L4, and the control gain compares L3 with L3 + C*; earlier drafts called the control gain the identity gain, and the code and result files call it `G_id`. The excess gain compares L3 + C* with L4 + C*.

The reference WIS and every gain (Table 2 and Tables S21, S24 to S34, S36, S37 and S42) are week-weighted. Tables S17 to S20, S22, S23 and S41 report row means, marked in their titles, so their WIS values differ slightly from the week-weighted ones. For example, the reference model in Brazil's future-years comparison has a WIS of 0.1675 week-weighted and 0.1681 as a row mean.

#### A. Baselines and model ladder

**Table S17. Forecast error of the baseline and ladder models in the future-years split (row mean).**

| Model | Brazil | Colombia | Peru | Taiwan |
|---|---:|---:|---:|---:|
| L0, seasonal naive | 0.4695 (1.00) | 0.3272 (1.00) | 0.6596 (1.00) | 0.0743 (1.00) |
| Negative binomial regression (L1 inputs) | 0.2185 (0.47) | 0.1426 (0.44) | 0.3232 (0.49) | 0.0475 (0.64) |
| TimesFM 2.5, zero-shot | 0.1928 (0.41) | not run | not run | not run |
| LightGBM L1, case history | 0.1700 (0.36) | 0.1226 (0.37) | 0.2442 (0.37) | 0.0316 (0.43) |
| LightGBM L2, + climate | 0.1690 (0.36) | 0.1224 (0.37) | 0.2431 (0.37) | 0.0319 (0.43) |
| LightGBM L3, + remote sensing and population | 0.1681 (0.36) | 0.1220 (0.37) | 0.2438 (0.37) | 0.0320 (0.43) |
| LightGBM L4, + embeddings | 0.1677 (0.36) | 0.1225 (0.37) | 0.2460 (0.37) | 0.0323 (0.43) |
| LightGBM L3 + C* | 0.1680 (0.36) | 0.1221 (0.37) | 0.2443 (0.37) | 0.0321 (0.43) |
| LightGBM L4 + C* | 0.1676 (0.36) | 0.1226 (0.37) | 0.2463 (0.37) | 0.0323 (0.44) |
| Rows scored | 2,703,183 | 319,930 | 45,754 | 141,645 |

*Note:* Mean WIS over rows, with the WIS relative to the seasonal naive baseline (L0) in parentheses. Every model is scored on the same rows, whose number is given in the last row. TimesFM was run for Brazil only. The selected control (C*) is average past cases (C1) in all four countries. L3, L4, L3 + C* and L4 + C* average seeds 0 to 2 in all four countries; the other models use one run. Taiwan includes test year 2021, and its L2 uses the shared configuration (Table S53). The earlier Taiwan values without 2021 used a separately tuned L2. The step from L0 to L1 changes both the inputs and the method.

**Table S18. Coverage of 80% prediction intervals in the future-years split.**

| Model | Brazil | Colombia | Peru | Taiwan |
|---|---|---|---|---|
| L0, seasonal naive | 0.77 / 0.40 | 0.81 / 0.53 | 0.73 / 0.47 | 0.95 / 0.00 |
| Negative binomial regression (L1 inputs) | 0.94 / 0.77 | 0.96 / 0.81 | 0.93 / 0.84 | 0.99 / 0.42 |
| TimesFM 2.5, zero-shot | 0.82 / 0.60 | not run | not run | not run |
| LightGBM L1, case history | 0.91 / 0.68 | 0.93 / 0.72 | 0.84 / 0.70 | 0.98 / 0.24 |
| LightGBM L2, + climate | 0.90 / 0.68 | 0.93 / 0.72 | 0.83 / 0.68 | 0.98 / 0.23 |
| LightGBM L3, + remote sensing and population | 0.90 / 0.67 | 0.93 / 0.72 | 0.83 / 0.68 | 0.98 / 0.25 |
| LightGBM L4, + embeddings | 0.90 / 0.67 | 0.93 / 0.71 | 0.83 / 0.68 | 0.98 / 0.24 |
| LightGBM L3 + C* | 0.90 / 0.66 | 0.93 / 0.71 | 0.83 / 0.67 | 0.98 / 0.24 |
| LightGBM L4 + C* | 0.90 / 0.66 | 0.93 / 0.71 | 0.83 / 0.67 | 0.98 / 0.23 |

*Note:* Share of observations inside the 80% interval, first over all rows and then over rows with at least one case. Models, rows and seeds as in Table S17.

**Table S19. Mean forecast error along the model ladder in each comparison (row mean).**

| Comparison | C* | L0 | L1 | L2 | L3 | L4 |
|---|---|---:|---:|---:|---:|---:|
| Brazil, future years | C1 | 0.4695 | 0.1700 | 0.1690 | 0.1681 | 0.1677 |
| Brazil, held-out states | C1 | – | – | – | 0.1689 | 0.1690 |
| Brazil, held-out regions | C1 | – | – | – | 0.1706 | 0.1709 |
| Brazil, held-out districts | C1 | – | – | – | 0.1683 | 0.1679 |
| Brazil, held-out country | C3 | – | – | – | 0.1914 | 0.1921 |
| Colombia, future years | C1 | 0.3272 | 0.1226 | 0.1224 | 0.1220 | 0.1225 |
| Colombia, held-out states | C1 | – | – | – | 0.1225 | 0.1232 |
| Colombia, held-out regions | C1 | – | – | – | 0.1245 | 0.1255 |
| Colombia, held-out districts | C1 | – | – | – | 0.1223 | 0.1229 |

*Table S19 (continued)*

| Comparison | C* | L0 | L1 | L2 | L3 | L4 |
|---|---|---|---|---|---|---|
| Colombia, held-out country | C2 | – | – | – | 0.1238 | 0.1239 |
| Peru, future years | C1 | 0.6596 | 0.2442 | 0.2431 | 0.2438 | 0.2460 |
| Peru, held-out country | C3 | – | – | – | 0.2383 | 0.2367 |
| Taiwan, future years | C1 | 0.0743 | 0.0316 | 0.0319 | 0.0320 | 0.0323 |
| Taiwan, held-out country | C3 | – | – | – | 0.0205 | 0.0250 |
| Brazil, future years, per-level tuning (sensitivity) | C1 | 0.4695 | 0.1700 | 0.1690 | 0.1714 | 0.1710 |
| Taiwan, future years, Brazil parameters (sensitivity, without 2021) | C1 | – | – | – | 0.0398 | 0.0402 |

*Note:* Mean WIS over rows (lower is better), on rows common to every model. L0 to L2 were run in the future-years split only. L3, L4, L3 + C* and L4 + C* average seeds 0 to 2 where three seeds exist. That is the case in all future-years comparisons, with Brazil held out and in Colombia's four within-country splits. With Peru or Taiwan held out, only L3 + C* and L4 + C* have three seeds. Other comparisons use seed 0. Taiwan, future years, includes 2021 and uses the shared configuration for L2 (Table S53). There are two sensitivity rows. In Brazil, future years, each model was tuned separately, giving L3 and L4 with 31 leaves as in the original protocol, with the earlier control selection. In Taiwan, future years, without 2021, the models used Brazil's configuration and selected control.

**Table S20. Forecast error with the location control, and interval coverage, in each comparison (row mean).**

| Comparison | L3 + C* | L4 + C* | 80% coverage of L4 + C*, all rows / cases > 0 | Rows |
|---|---|---|---|---|
| Brazil, future years | 0.1680 | 0.1676 | 0.90 / 0.66 | 2,703,183 |
| Brazil, held-out states | 0.1690 | 0.1690 | 0.90 / 0.66 | 2,703,183 |
| Brazil, held-out regions | 0.1707 | 0.1710 | 0.90 / 0.66 | 2,703,183 |
| Brazil, held-out districts | 0.1683 | 0.1680 | 0.90 / 0.66 | 2,703,183 |
| Brazil, held-out country | 0.1911 | 0.1924 | 0.88 / 0.64 | 2,703,183 |
| Colombia, future years | 0.1221 | 0.1226 | 0.93 / 0.71 | 319,930 |
| Colombia, held-out states | 0.1225 | 0.1232 | 0.92 / 0.70 | 319,930 |
| Colombia, held-out regions | 0.1246 | 0.1254 | 0.92 / 0.70 | 319,930 |
| Colombia, held-out districts | 0.1224 | 0.1229 | 0.92 / 0.71 | 319,930 |
| Colombia, held-out country | 0.1249 | 0.1246 | 0.94 / 0.75 | 319,930 |
| Peru, future years | 0.2443 | 0.2463 | 0.83 / 0.67 | 45,754 |
| Peru, held-out country | 0.2392 | 0.2369 | 0.84 / 0.67 | 45,754 |
| Taiwan, future years | 0.0321 | 0.0323 | 0.98 / 0.23 | 141,645 |
| Taiwan, held-out country | 0.0216 | 0.0253 | 0.99 / 0.61 | 141,645 |
| Brazil, future years, per-level tuning (sensitivity) | 0.1714 | 0.1710 | 0.90 / 0.68 | 2,703,183 |
| Taiwan, future years, Brazil parameters (sensitivity, without 2021) | 0.0401 | 0.0403 | 0.98 / 0.24 | 113,430 |

*Note:* Mean WIS over rows. Rows and seeds as in Table S19.

#### B. Primary family: secondary gains, control selection and equivalence

**Table S21. Reference error, embedding gain and control gain in each comparison.**

| Comparison | Reference WIS (L3) | WIS of L4 + C* | Embedding gain, % (95% CI) | Control gain, % (95% CI) | DM p, excess gain | Holm TOST p (±1%) |
|---|---|---|---|---|---|---|
| Brazil, future years | 0.1675 | 0.1671 | +0.26 (−0.31 to 0.82) | +0.03 (−0.11 to 0.18) | 0.39 | 0.036 |
| Brazil, held-out states | 0.1683 | 0.1684 | −0.02 (−0.34 to 0.30) | −0.05 (−0.13 to 0.02) | 1.0 | <0.001 |
| Brazil, held-out regions | 0.1701 | 0.1704 | −0.13 (−0.35 to 0.09) | −0.03 (−0.14 to 0.08) | 0.090 | <0.001 |
| Brazil, held-out districts | 0.1678 | 0.1674 | +0.22 (−0.32 to 0.76) | +0.02 (−0.01 to 0.05) | 0.43 | 0.018 |
| Brazil, held-out country | 0.1907 | 0.1917 | −0.39 (−1.34 to 0.55) | +0.15 (−0.89 to 1.18) | <0.001 | 0.055 |
| Colombia, future years | 0.1222 | 0.1228 | −0.38 (−0.61 to −0.16) | −0.06 (−0.32 to 0.19) | 0.0012 | <0.001 |
| Colombia, held-out states | 0.1226 | 0.1233 | −0.60 (−0.81 to −0.39) | −0.06 (−0.09 to −0.03) | <0.001 | 0.0027 |
| Colombia, held-out regions | 0.1246 | 0.1255 | −0.82 (−1.14 to −0.49) | −0.08 (−0.14 to −0.02) | 0.0030 | 0.27 |
| Colombia, held-out districts | 0.1224 | 0.1231 | −0.54 (−0.78 to −0.30) | −0.10 (−0.12 to −0.08) | 0.0017 | 0.0018 |
| Colombia, held-out country | 0.1240 | 0.1247 | −0.04 (−0.37 to 0.28) | −0.81 (−1.16 to −0.46) | 0.27 | 0.0066 |
| Peru, future years | 0.2417 | 0.2442 | −0.91 (−1.62 to −0.20) | −0.20 (−0.92 to 0.52) | 0.022 | 0.85 |
| Peru, held-out country | 0.2362 | 0.2351 | +0.68 (−0.59 to 1.96) | −0.44 (−0.80 to −0.07) | 0.088 | 0.90 |
| Taiwan, future years | 0.0312 | 0.0316 | −0.95 (−1.71 to −0.18) | −0.41 (−0.97 to 0.16) | 0.042 | 0.78 |
| Taiwan, held-out country | 0.0202 | 0.0245 | −21.60 (−31.82 to −11.39) | −3.36 (−6.52 to −0.20) | 0.0039 | 1.0 |

*Note:* Same rows, tests and selected controls as Table 2. The reference WIS is week-weighted. The embedding gain compares L3 with L4, and the control gain compares L3 with L3 + C*. DM p is the unadjusted p value of the excess gain. The Holm TOST p tests equivalence within ±1% of the reference WIS, with Holm adjustment over the 14 comparisons. As in Table 2, estimates average seeds 0 to 2 where three seeds exist. With Peru or Taiwan held out, the embedding and control gains use seed 0. Test years and rows are in Table 1.

**Table S22. Selection of the location control at the 2019 origin: reference model, average past cases, coordinates and shuffled embedding (row mean).**

| Comparison | Reference (L3 or N3) | + C1 | + C2 | + C3 | Selected C* |
|---|---|---|---|---|---|
| Brazil, future years | 0.17525 | 0.17446 | 0.17715 | 0.17681 | C1 |
| Brazil, held-out states | 0.17542 | 0.17597 | 0.17733 | 0.17757 | C1 |
| Brazil, held-out regions | 0.17715 | 0.17749 | 0.17989 | 0.17916 | C1 |
| Brazil, held-out districts | 0.17500 | 0.17517 | 0.17716 | 0.17684 | C1 |
| Brazil, held-out country | 0.19102 | – | 0.19168 | 0.19182 | C3 |
| Colombia, future years | 0.17019 | 0.17046 | 0.17045 | 0.17042 | C1 |
| Colombia, held-out states | 0.17169 | 0.17176 | 0.17211 | 0.17219 | C1 |
| Colombia, held-out regions | 0.17311 | 0.17315 | 0.17387 | 0.17381 | C1 |
| Colombia, held-out districts | 0.17158 | 0.17167 | 0.17201 | 0.17225 | C1 |
| Colombia, held-out country | 0.18208 | – | 0.17809 | 0.18352 | C2 |
| Peru, future years | 0.16194 | 0.16258 | 0.16431 | 0.16537 | C1 |
| Peru, held-out country | 0.13892 | – | 0.14227 | 0.13997 | C3 |
| Taiwan, future years | 0.00411 | 0.00413 | 0.00439 | 0.00402 | C1 |
| Taiwan, held-out country | 0.00656 | – | 0.00912 | 0.00784 | C3 |
| Brazil, future years, without case history | 0.35428 | – | 0.34633 | 0.35963 | C2 |
| Brazil, held-out states, without case history | 0.36193 | – | 0.35921 | 0.36860 | C2 |

*Note:* Mean WIS over rows (not week-weighted) at the 2019 validation origin (trained on 2018 targets and scored on 2019 targets; horizons 1 and 4) of the reference model and of the reference model plus each candidate control. C3 is a mean over three seeds; the other columns use one seed. The selection rule (D19) is given in full in Supplementary Methods, section B. In short, candidates whose forecasts equal the reference model's are excluded. Each remaining candidate has a standard error $\mathrm{SE}_k = \sigma_{\mathrm{seed}}/\sqrt{n_k}$, where $\sigma_{\mathrm{seed}}$ is the seed SD pooled over the multi-seed candidates (C3, C4, C4s) and $n_k$ is the number of seeds; single-seed candidates therefore get $\mathrm{SE}_k = \sigma_{\mathrm{seed}}$. The tie set is every candidate with $\mathrm{Score}_k - \mathrm{Score}_{\mathrm{best}} \le 2\sqrt{\mathrm{SE}_{\mathrm{best}}^2 + \mathrm{SE}_k^2}$, and its first member in the order C1, C3, C2, C4s, C4, C5 is selected. Without case history the order is C3, C2, C6, C4s, C4. The plain reference model was not a candidate, so a control was selected even where every candidate scored worse than the reference model. A rule that allows "no control" is a sensitivity analysis (Table S45). A dash means not a candidate. C1 is not defined across countries. C4s is not a candidate in Peru and Taiwan, where its range equals that of C4. C5 is not a candidate without case history, where C1 forms its own step. C6 is used only in the future-years split without case history. A control scored below L3 only in Brazil (future years), Colombia (held-out country) and Taiwan (future years). In Taiwan the lower-scoring C3 was not selected because of the order of preference. Taiwan's 2019 validation WIS (about 0.004) rests on 56 non-zero validation targets, so its choice of control is effectively arbitrary. The test-year results of the two comparisons without case history are in Tables S41 and S42.

**Table S23. Selection of the location control at the 2019 origin: random smooth fields, snapshot and district label (row mean).**

| Comparison | + C4 | + C4s | + C5 | + C6 |
|---|---:|---:|---:|---:|
| Brazil, future years | 0.17782 | 0.17840 | 0.17732 | – |
| Brazil, held-out states | 0.17773 | 0.17900 | 0.17766 | – |
| Brazil, held-out regions | 0.18093 | 0.18162 | 0.18033 | – |
| Brazil, held-out districts | 0.17799 | 0.17855 | 0.17731 | – |
| Brazil, held-out country | 0.19235 | 0.19247 | 0.19252 | – |
| Colombia, future years | 0.17096 | 0.17088 | 0.17093 | – |
| Colombia, held-out states | 0.17288 | 0.17279 | 0.17249 | – |
| Colombia, held-out regions | 0.17490 | 0.17453 | 0.17455 | – |
| Colombia, held-out districts | 0.17269 | 0.17269 | 0.17279 | – |
| Colombia, held-out country | 0.18349 | 0.18367 | 0.18290 | – |
| Peru, future years | 0.16684 | 0.16664 | 0.16372 | – |
| Peru, held-out country | 0.14111 | – | 0.14149 | – |
| Taiwan, future years | 0.00439 | – | 0.00446 | – |
| Taiwan, held-out country | 0.00782 | – | 0.01108 | – |
| Brazil, future years, without case history | 0.34895 | 0.35076 | – | 0.34722 |
| Brazil, held-out states, without case history | 0.35977 | 0.36747 | – | – |

*Note:* As Table S22. C4 and C4s are means over three seeds.

**Table S24. Equivalence of the excess gain at margins of ±0.5%, ±1% and ±2% of the reference WIS.**

| Comparison | Excess gain, % (90% CI) | ±0.5%: unadjusted / Holm | ±1%: unadjusted / Holm | ±2%: unadjusted / Holm |
|---|---:|---|---|---|
| Brazil, future years | +0.23 (−0.23 to 0.69) | no / no | yes / yes | yes / yes |
| Brazil, held-out states | 0.00 (−0.28 to 0.28) | yes / yes | yes / yes | yes / yes |
| Brazil, held-out regions | −0.16 (−0.31 to −0.01) | yes / yes | yes / yes | yes / yes |
| Brazil, held-out districts | +0.20 (−0.23 to 0.62) | no / no | yes / yes | yes / yes |
| Brazil, held-out country | −0.68 (−0.89 to −0.47) | no / no | yes / no | yes / yes |
| Colombia, future years | −0.42 (−0.60 to −0.25) | no / no | yes / yes | yes / yes |
| Colombia, held-out states | −0.52 (−0.70 to −0.34) | no / no | yes / yes | yes / yes |
| Colombia, held-out regions | −0.68 (−1.01 to −0.36) | no / no | no / no (near margin) | yes / yes |
| Colombia, held-out districts | −0.45 (−0.64 to −0.25) | no / no | yes / yes | yes / yes |
| Colombia, held-out country | +0.22 (−0.12 to 0.55) | no / no | yes / yes | yes / yes |

*Table S24 (continued)*

| Comparison | Excess gain, % (90% CI) | ±0.5%: unadjusted / Holm | ±1%: unadjusted / Holm | ±2%: unadjusted / Holm |
|---|---|---|---|---|
| Peru, future years | −0.82 (−1.37 to −0.26) | no / no | no / no | yes / yes |
| Peru, held-out country | +0.93 (0.04 to 1.83) | no / no | no / no | yes / no |
| Taiwan, future years | −0.71 (−1.28 to −0.15) | no / no | no / no | yes / yes |
| Taiwan, held-out country | −18.29 (−27.77 to −8.81) | no / no | no / no | no / no |
| Comparisons within the margin (of 14) | | 2 / 2 | 9 / 8 | 13 / 12 |

*Note:* The excess gain is shown with its 90% CI. A comparison is equivalent within the margin when both one-sided tests (TOST) reject at 5%, either unadjusted or with the TOST p values Holm-adjusted over the 14 comparisons. "Near margin" means that a CI bound lies within 0.05 percentage points of the margin. The ±1% margin was chosen after the Brazil and Peru future-years results had been seen. Equivalence for Brazil (future years, held-out states, regions and districts) and Colombia (held-out country) assumes that the weekly loss difference does not drift across seasons. In these five comparisons the surrogate-series check shows that the main test finds effects too often (Table S47).

#### C. Robustness of the primary family

##### Table S25. Excess gain at horizons 1 and 4 separately.

| Comparison | Horizon 1: % (95% CI); p | Horizon 4: % (95% CI); p | Horizons 1 to 4, future years only: % (95% CI); p; years |
|---|---|---|---|
| Brazil, future years | +0.11 (−0.14 to 0.36), 0.36 | +0.32 (−0.48 to 1.13), 0.40 | +0.25 (−0.33 to 0.82), 0.38; 2020–24 |
| Brazil, held-out states | −0.01 (−0.19 to 0.18), 0.95 | 0.00 (−0.47 to 0.47), 0.99 | – |
| Brazil, held-out regions | −0.13 (−0.24 to −0.02), 0.022 | −0.18 (−0.43 to 0.07), 0.14 | – |
| Brazil, held-out districts | +0.11 (−0.14 to 0.35), 0.37 | +0.26 (−0.46 to 0.98), 0.46 | – |
| Brazil, held-out country | −0.59 (−0.80 to −0.38), <0.001 | −0.77 (−1.17 to −0.36), 0.0010 | – |
| Colombia, future years | −0.29 (−0.43 to −0.15), <0.001 | −0.56 (−0.87 to −0.25), 0.0021 | −0.41 (−0.64 to −0.18), 0.0024; 2020–22 |
| Colombia, held-out states | −0.42 (−0.56 to −0.27), <0.001 | −0.64 (−0.96 to −0.32), 0.0010 | – |
| Colombia, held-out regions | −0.67 (−0.90 to −0.45), <0.001 | −0.71 (−1.31 to −0.11), 0.025 | – |
| Colombia, held-out districts | −0.35 (−0.53 to −0.17), 0.0015 | −0.55 (−0.88 to −0.23), 0.0032 | – |
| Colombia, held-out country | −0.07 (−0.29 to 0.15), 0.49 | +0.51 (−0.13 to 1.15), 0.11 | – |
| Peru, future years | −0.22 (−0.51 to 0.07), 0.13 | −1.25 (−2.24 to −0.26), 0.017 | −0.69 (−1.38 to −0.01), 0.047; 2020–23 |
| Peru, held-out country | +0.34 (−0.09 to 0.78), 0.11 | +1.34 (−0.19 to 2.87), 0.081 | – |
| Taiwan, future years | −0.38 (−0.76 to 0.01), 0.054 | −1.01 (−2.07 to 0.04), 0.059 | −0.46 (−1.09 to 0.17), 0.14; 2020–24 (no 2021) |
| Taiwan, held-out country | −14.63 (−22.48 to −6.78), 0.0011 | −20.65 (−35.08 to −6.22), 0.0081 | – |

*Note:* The future-years comparisons also show a secondary estimate pooled over horizons 1 to 4. It is not part of the Holm family and uses only test years scored at all four horizons. For Taiwan it therefore leaves out 2021, whose runs cover horizons 1 and 4 only.

##### Table S26. Excess gain under the two stricter checks.

| Comparison | Excess gain, % (95% CI) | p / Holm p | B = 4: 95% CI | B = 4: p / Holm p | Surrogate-calibrated p / Holm p | 95% CI, reference WIS estimated |
|---|---|---|---|---|---|---|
| Brazil, future years | +0.23 (−0.33 to 0.79) | 0.39 / 1.0 | −0.53 to 1.00 | 0.45 / 1.0 | 0.51 / 1.0 | −0.28 to 0.75 |
| Brazil, held-out states | 0.00 (−0.34 to 0.34) | 1.0 / 1.0 | −0.57 to 0.56 | 1.0 / 1.0 | 1.0 / 1.0 | −0.34 to 0.34 |
| Brazil, held-out regions | −0.16 (−0.34 to 0.03) | 0.090 / 0.53 | −0.49 to 0.18 | 0.26 / 1.0 | 0.21 / 1.0 | −0.36 to 0.05 |
| Brazil, held-out districts | +0.20 (−0.32 to 0.71) | 0.43 / 1.0 | −0.49 to 0.89 | 0.47 / 1.0 | 0.52 / 1.0 | −0.28 to 0.68 |
| Brazil, held-out country | −0.68 (−0.94 to −0.42) | <0.001 / <0.001 | −1.06 to −0.31 | 0.0073 / 0.095 | <0.001 / 0.0070 | −1.01 to −0.35 |

*Table S26 (continued)*

| Comparison | Excess gain, % (95% CI) | p / Holm p | B = 4: 95% CI | B = 4: p / Holm p | Surrogate-calibrated p / Holm p | 95% CI, reference WIS estimated |
|---|---|---|---|---|---|---|
| Colombia, future years | −0.42 (−0.64 to −0.21) | 0.0012 / 0.014 | −0.70 to −0.14 | 0.014 / 0.15 | 0.0035 / 0.038 | −0.64 to −0.21 |
| Colombia, held-out states | −0.52 (−0.74 to −0.30) | <0.001 / 0.0034 | −0.71 to −0.33 | 0.0016 / 0.022 | <0.001 / 0.0070 | −0.76 to −0.29 |
| Colombia, held-out regions | −0.68 (−1.08 to −0.29) | 0.0030 / 0.030 | −1.09 to −0.28 | 0.0093 / 0.11 | <0.001 / 0.0070 | −1.13 to −0.24 |
| Colombia, held-out districts | −0.45 (−0.68 to −0.21) | 0.0017 / 0.019 | −0.75 to −0.14 | 0.015 / 0.15 | 0.0060 / 0.054 | −0.68 to −0.21 |
| Colombia, held-out country | +0.22 (−0.19 to 0.63) | 0.27 / 1.0 | −0.58 to 1.01 | 0.49 / 1.0 | 0.46 / 1.0 | −0.17 to 0.60 |
| Peru, future years | −0.82 (−1.49 to −0.14) | 0.022 / 0.18 | −1.94 to 0.31 | 0.11 / 0.68 | 0.040 / 0.28 | −1.55 to −0.08 |
| Peru, held-out country | +0.93 (−0.16 to 2.02) | 0.088 / 0.53 | 0.25 to 1.62 | 0.020 / 0.18 | 0.0050 / 0.050 | −0.20 to 2.06 |
| Taiwan, future years | −0.71 (−1.40 to −0.03) | 0.042 / 0.29 | −1.36 to −0.07 | 0.037 / 0.27 | 0.0070 / 0.056 | −1.65 to 0.22 |
| Taiwan, held-out country | −18.29 (−29.80 to −6.78) | 0.0039 / 0.035 | −34.29 to −2.29 | 0.034 / 0.27 | 0.048 / 0.29 | −43.79 to 7.21 |

*Note:* Main test as in Table 2, with Holm adjustment over the 14 comparisons. B = 4: the strict-variance check, the same test with 4 cosine terms in the variance, which is less affected by slow drift across seasons. Surrogate-calibrated p: the surrogate-series check, which compares the observed statistic with 2,000 iterative amplitude-adjusted Fourier transform (IAAFT) surrogates of the comparison's own weekly loss-difference series. Last column: delta-method 95% CI of the percentage, which allows for uncertainty in the reference WIS. In Brazil, future years (three seeds), the seed standard deviation of the excess gain is 0.00002, against a standard error of 0.00044. The standard errors of the three-seed comparisons average over seeds. In each of them the seed standard deviation of the excess gain is at most about the size of its standard error (Brazil held out 0.00024 against 0.00023). With Peru held out, the excess gain was positive before adjustment under both stricter checks (B = 4 p = 0.020; surrogate-calibrated p = 0.0050). After Holm correction the B = 4 p was 0.18, and the surrogate-calibrated p sat exactly at the threshold (0.04998). This is the only gain in the family that reaches the threshold under any of the tests.

**Table S27. Excess gain on the case scale, on all rows and without the Espírito Santo exclusion.**

| Comparison | Case scale, % | Case scale, p | All rows, % | All rows, p | Earlier exclusions, % | Earlier exclusions, p |
|---|---|---|---|---|---|---|
| Brazil, future years | −0.03 | 0.92 | +0.26 | 0.39 | +0.23 | 0.40 |
| Brazil, held-out states | −0.73 | 0.025 | +0.03 | 0.89 | −0.01 | 0.94 |
| Brazil, held-out regions | −1.30 | 0.065 | −0.10 | 0.40 | −0.19 | 0.045 |
| Brazil, held-out districts | −0.37 | 0.13 | +0.23 | 0.42 | +0.19 | 0.44 |
| Brazil, held-out country | −0.89 | 0.18 | −0.69 | <0.001 | −0.70 | <0.001 |
| Colombia, future years | −0.84 | 0.052 | −0.42 | <0.001 | – | – |
| Colombia, held-out states | −0.75 | 0.15 | −0.53 | <0.001 | – | – |
| Colombia, held-out regions | −0.78 | 0.20 | −0.71 | 0.0025 | – | – |
| Colombia, held-out districts | −1.05 | 0.074 | −0.45 | 0.0011 | – | – |
| Colombia, held-out country | +0.51 | 0.068 | +0.18 | 0.33 | – | – |
| Peru, future years | −1.35 | 0.077 | −0.73 | 0.046 | – | – |
| Peru, held-out country | −0.61 | 0.58 | +0.99 | 0.051 | – | – |
| Taiwan, future years | −0.13 | 0.12 | −0.66 | 0.062 | – | – |
| Taiwan, held-out country | −3.66 | 0.0033 | −18.46 | 0.0029 | – | – |

*Note:* Case scale: excess gain in the WIS of raw case counts, as a percentage of the case-scale reference WIS (secondary scale). All rows: without the origin-year filter. Earlier exclusions: Brazil scored without the Espírito Santo exclusion. A dash means not applicable.

#### D. Where the difference arises

**Table S28. Additive contributions of three types of target week to the excess gain.**

| Comparison | Total, % | Weeks with cases, % (95% CI) | Zeros listed by the source, % (95% CI) | Zero-filled weeks, % (95% CI) |
|---|---|---|---|---|
| Brazil, future years | +0.23 | +0.46 (−0.16 to 1.07) | −0.02 (−0.16 to 0.12) | −0.20 (−0.33 to −0.08) |
| Brazil, held-out states | 0.00 | +0.14 (−0.27 to 0.54) | +0.05 (−0.07 to 0.16) | −0.18 (−0.30 to −0.06) |
| Brazil, held-out regions | −0.16 | +0.22 (−0.02 to 0.47) | −0.03 (−0.17 to 0.11) | −0.35 (−0.52 to −0.18) |
| Brazil, held-out districts | +0.20 | +0.40 (−0.16 to 0.96) | 0.00 (−0.12 to 0.12) | −0.20 (−0.34 to −0.07) |
| Brazil, held-out country | −0.68 | +0.16 (−0.08 to 0.40) | −0.16 (−0.44 to 0.11) | −0.68 (−0.88 to −0.48) |
| Colombia, future years | −0.42 | +0.18 (−0.19 to 0.56) | – | −0.60 (−0.99 to −0.21) |
| Colombia, held-out states | −0.52 | +0.16 (−0.23 to 0.55) | – | −0.68 (−1.05 to −0.32) |
| Colombia, held-out regions | −0.68 | +0.07 (−0.20 to 0.35) | – | −0.76 (−1.21 to −0.30) |
| Colombia, held-out districts | −0.45 | +0.22 (−0.20 to 0.64) | – | −0.66 (−1.10 to −0.23) |
| Colombia, held-out country | +0.22 | +0.27 (0.06 to 0.49) | – | −0.05 (−0.51 to 0.41) |
| Peru, future years | −0.82 | −0.14 (−0.89 to 0.61) | – | −0.67 (−0.92 to −0.43) |
| Peru, held-out country | +0.93 | +1.20 (0.11 to 2.29) | – | −0.27 (−0.42 to −0.13) |
| Taiwan, future years (with 2021) | −0.71 | −0.24 (−0.73 to 0.25) | – | −0.47 (−1.12 to 0.18) |
| Taiwan, held-out country (with 2021) | −18.29 | +1.33 (−1.01 to 3.67) | – | −19.62 (−30.92 to −8.32) |
| Brazil, future years, per-level tuning, 31 leaves (sensitivity) | +0.25 | +0.27 (−0.04 to 0.59) | +0.01 (−0.08 to 0.11) | −0.03 (−0.09 to 0.02) |
| Taiwan, future years, without 2021 | −0.44 | −0.07 (−0.35 to 0.20) | – | −0.36 (−1.04 to 0.31) |
| Taiwan, held-out country, without 2021 | −11.16 | +1.33 (−1.12 to 3.77) | – | −12.48 (−16.99 to −7.97) |
| Brazil, future years, without case history (N3 + C2 vs N4 + C2) | −0.43 | +0.77 (−1.27 to 2.80) | −0.29 (−0.78 to 0.20) | −0.91 (−1.46 to −0.35) |
| Brazil, held-out states, without case history (N3 + C2 vs N4 + C2) | −0.01 | +0.99 (−0.54 to 2.51) | −0.27 (−0.72 to 0.18) | −0.73 (−1.17 to −0.29) |

*Note:* Each contribution is a percentage of the comparison's reference WIS, and the three contributions add up exactly to the total. Zeros listed by the source are zero weeks that the source itself reports (Brazil 2020 to 2021 test years only); zero-filled weeks are weeks missing from the source and set to zero. A dash means that the comparison has no rows of this type. The extra rows are Brazil, future years, with each model tuned separately (31 leaves); Taiwan without 2021; and Brazil, future years and held-out states, without case history. The rows without case history compare N4 + C2 with N3 + C2, as percentages of the WIS of N3. Estimates average seeds 0 to 2 where three seeds exist, as in Table 2.

**Table S29. Excess gain within each type of target week.**

| Comparison | Cases > 0: share | Cases > 0: % (95% CI) | Zeros listed by the source: share | Zeros listed by the source: % (95% CI) | Zero-filled: share | Zero-filled: % (95% CI) |
|---|---|---|---|---|---|---|
| Brazil, future years | 27% | +0.30 (−0.16 to 0.76) | 21% | −0.27 (−2.74 to 2.20) | 52% | −1.67 (−2.65 to −0.69) |
| Brazil, held-out states | 27% | 0.00 (−0.42 to 0.42) | 21% | +0.72 (−1.33 to 2.78) | 52% | −1.48 (−2.58 to −0.38) |
| Brazil, held-out regions | 27% | +0.28 (−0.01 to 0.58) | 21% | −0.34 (−2.71 to 2.03) | 52% | −2.75 (−4.39 to −1.11) |
| Brazil, held-out districts | 27% | +0.27 (−0.18 to 0.72) | 21% | +0.07 (−2.14 to 2.29) | 52% | −1.59 (−2.65 to −0.54) |
| Brazil, held-out country | 27% | +0.44 (0.03 to 0.85) | 21% | −1.89 (−5.43 to 1.64) | 52% | −5.18 (−6.39 to −3.97) |
| Colombia, future years | 21% | +0.24 (−0.31 to 0.79) | – | – | 79% | −2.17 (−3.52 to −0.81) |
| Colombia, held-out states | 21% | +0.19 (−0.40 to 0.77) | – | – | 79% | −2.45 (−3.73 to −1.16) |
| Colombia, held-out regions | 21% | +0.09 (−0.33 to 0.52) | – | – | 79% | −2.55 (−4.06 to −1.03) |
| Colombia, held-out districts | 21% | +0.26 (−0.37 to 0.89) | – | – | 79% | −2.40 (−3.92 to −0.89) |
| Colombia, held-out country | 21% | +0.37 (0.07 to 0.66) | – | – | 79% | −0.13 (−2.25 to 1.98) |

*Table S29 (continued)*

| Comparison | Cases > 0: share | Cases > 0: % (95% CI) | Zeros listed by the source: share | Zeros listed by the source: % (95% CI) | Zero-filled: share | Zero-filled: % (95% CI) |
|---|---|---|---|---|---|---|
| Peru, future years | 44% | −0.27 (−1.11 to 0.57) | – | – | 56% | −5.64 (−8.10 to −3.19) |
| Peru, held-out country | 44% | +1.52 (0.14 to 2.90) | – | – | 56% | −2.69 (−4.31 to −1.08) |
| Taiwan, future years (with 2021) | 2% | −0.10 (−1.15 to 0.95) | – | – | 98% | −8.59 (−20.34 to 3.16) |
| Taiwan, held-out country (with 2021) | 2% | +2.41 (0.56 to 4.26) | – | – | 98% | −51.00 (−79.68 to −22.33) |
| Brazil, future years, per-level tuning, 31 leaves (sensitivity) | 27% | +0.22 (−0.03 to 0.46) | 21% | +0.30 (−1.54 to 2.14) | 52% | −0.06 (−0.65 to 0.54) |
| Taiwan, future years, without 2021 | 3% | −0.15 (−1.21 to 0.91) | – | – | 97% | −6.74 (−19.50 to 6.02) |
| Taiwan, held-out country, without 2021 | 3% | +2.16 (0.27 to 4.04) | – | – | 97% | −35.22 (−48.16 to −22.27) |
| Brazil, future years, without case history (N3 + C2 vs N4 + C2) | 27% | +0.58 (−1.16 to 2.31) | 21% | −6.09 (−17.44 to 5.25) | 52% | −15.77 (−24.77 to −6.76) |
| Brazil, held-out states, without case history (N3 + C2 vs N4 + C2) | 27% | +1.04 (−0.24 to 2.32) | 21% | −5.38 (−14.67 to 3.91) | 52% | −11.40 (−17.98 to −4.81) |

*Note:* Share is the share of rows. Each estimate is a percentage of that stratum's own reference WIS, with weeks weighted equally; these estimates are not shares of the total (Table S28). Rows as in Table S28. All strata are defined by the outcome and are descriptive. With Brazil held out, the estimate in the stratum of zeros listed by the source was −1.89% (95% CI −5.43 to 1.64). The additive contribution of the same rows to the total was −0.16 (Table S28; −0.44 to 0.11). Both 95% CIs include zero, and all zeros listed by the source fall in 2020 to 2021.

#### Table S30. Excess gain by test year.

| Comparison | 2020 | 2021 | 2022 | 2023 | 2024 |
|---|---|---|---|---|---|
| Brazil, future years | −0.22 | −0.67* | +0.12 | +0.42 | +0.88 |
| Brazil, held-out states | −0.38 | −0.77* | −0.07* | +0.32 | +0.41 |
| Brazil, held-out regions | −0.19* | −0.97* | −0.05 | +0.05 | +0.02 |
| Brazil, held-out districts | −0.15 | −0.66* | +0.07 | +0.37 | +0.78 |
| Brazil, held-out country | −0.17 | −1.94* | −0.68* | −0.67* | −0.46* |
| Colombia, future years | −0.65* | −0.36 | −0.21 | – | – |
| Colombia, held-out states | −0.62* | −0.64 | −0.31* | – | – |
| Colombia, held-out regions | −0.64 | −0.87* | −0.59* | – | – |
| Colombia, held-out districts | −0.69* | −0.42 | −0.18 | – | – |
| Colombia, held-out country | +0.80* | −0.31 | −0.02 | – | – |
| Peru, future years | −1.50 | −1.35 | −0.58 | −0.09 | – |
| Peru, held-out country | +1.31 | +1.40 | +1.11 | +0.16 | – |
| Taiwan, future years | −13.76 (−3.4) | −65.39 (−2.0) | +2.76 (+0.2) | −0.28 (−4.0) | −2.57 (−1.9) |
| Taiwan, held-out country | −38.01* (−31.8) | −209.94 (−80.4) | −88.10* (−29.5) | −1.50 (−11.0) | −26.83* (−31.8) |

*Note:* Percentage of that year's reference WIS. An asterisk marks unadjusted $p < 0.05$; CIs are in the source file. For Taiwan the absolute excess gain ($\times 10^{-4}$ log-WIS) is given in parentheses, because the reference WIS is close to zero outside the 2023 outbreak and the percentages are unstable (2021 had no local cases). Context: 2020 to 2021, COVID-19 pandemic; Peru 2023, record season; Taiwan 2023, outbreak; Brazil 2024, record epidemic.

In all five Brazilian comparisons the excess gain was lowest in 2021, a year of the COVID-19 pandemic (−0.66% to −1.94%; each unadjusted $p < 0.05$) [33,34]. In the four within-Brazil comparisons it was highest in 2023 or 2024. The largest yearly estimate in Brazil was in the future-years comparison in 2024, the year of Brazil's largest recorded epidemic (+0.88%, 95% CI −0.93 to 2.69) [35]. Colombia's four within-country comparisons were negative in every year. Peru (future years) was negative in every year and closest to zero in its 2023 record season (−0.09%) [36]. Taiwan's yearly percentages are unstable because its reference WIS is near zero outside the 2023 outbreak. In 2023 the excess gain was −0.28% for future years and −1.50% with Taiwan held out.

**Table S31. Upper quantiles of the forecasts on rows with zero cases.**

| Comparison | Zero-case share of rows | Δq0.9, log(1 + cases) (95% CI); p | Δq0.9, cases (95% CI) | Zero-row contribution, log-WIS × $10^{-4}$ (95% CI) | Share from quantiles above the median | Share from q0.9 alone |
|---|---|---|---|---|---|---|
| Brazil, future years | 73% | +0.006 (0.001 to 0.011); 0.029 | +0.003 (−0.008 to 0.014) | −3.8 (−7.3 to −0.2) | 0.77 | 0.24 |
| Brazil, held-out states | 73% | +0.009 (0.004 to 0.015); 0.0013 | +0.001 (−0.007 to 0.009) | −2.3 (−5.5 to 0.9) | 1.27 | 0.63 |
| Brazil, held-out regions | 73% | +0.027 (0.019 to 0.035); <0.001 | +0.031 (0.020 to 0.042) | −6.5 (−10.6 to −2.3) | 1.19 | 0.65 |
| Brazil, held-out districts | 73% | +0.009 (0.004 to 0.015); 0.0011 | +0.009 (−0.0004 to 0.019) | −3.4 (−6.8 to −0.02) | 1.00 | 0.43 |
| Brazil, held-out country | 73% | +0.042 (0.027 to 0.056); <0.001 | +0.045 (0.011 to 0.078) | −16.1 (−22.8 to −9.4) | 0.99 | 0.45 |
| Colombia, future years | 79% | +0.017 (0.010 to 0.025); <0.001 | +0.020 (0.004 to 0.036) | −7.4 (−12.1 to −2.6) | 0.97 | 0.40 |
| Colombia, held-out states | 79% | +0.019 (0.013 to 0.026); <0.001 | +0.018 (0.004 to 0.032) | −8.4 (−12.9 to −3.9) | 1.04 | 0.40 |
| Colombia, held-out regions | 79% | +0.025 (0.018 to 0.032); <0.001 | +0.017 (−0.002 to 0.036) | −9.4 (−15.1 to −3.8) | 1.11 | 0.47 |
| Colombia, held-out districts | 79% | +0.017 (0.010 to 0.025); <0.001 | +0.018 (0.003 to 0.033) | −8.1 (−13.4 to −2.8) | 1.06 | 0.36 |
| Colombia, held-out country | 79% | +0.007 (0.0004 to 0.014); 0.041 | +0.007 (−0.009 to 0.022) | −0.7 (−6.4 to 5.0) | 2.94 | 1.98 |
| Peru, future years | 56% | +0.036 (0.019 to 0.053); <0.001 | +0.054 (0.026 to 0.081) | −16.3 (−22.2 to −10.4) | 0.78 | 0.25 |
| Peru, held-out country | 56% | +0.017 (0.011 to 0.023); <0.001 | +0.035 (0.018 to 0.052) | −6.4 (−9.8 to −3.0) | 0.63 | 0.34 |
| Taiwan, future years | 98% | +0.002 (−0.002 to 0.005); 0.33 | +0.001 (−0.004 to 0.006) | −1.5 (−3.5 to 0.6) | 1.14 | 0.26 |
| Taiwan, held-out country | 98% | +0.093 (0.061 to 0.125); <0.001 | +0.132 (0.082 to 0.181) | −39.6 (−62.3 to −16.8) | 0.95 | 0.51 |

*Note:* Δq0.9 is the mean 0.9 quantile of L4 + C* minus that of L3 + C* on rows with zero cases, on the log(1 + cases) scale and on the case scale, with a 95% CI from the weekly series. The zero-row contribution is the additive contribution of all zero-case rows (recorded and filled) to the absolute excess gain. The last two columns give the share of that contribution from the quantile levels above the median (0.6 to 0.9) and from the 0.9 quantile alone; values above 1 mean that the lower quantiles offset part of the loss. Colombia's held-out-country shares are unstable because its contribution from zero-case rows is close to zero.

#### Supporting analyses

The analyses in Tables S32 to S35 are exploratory and not adjusted for multiplicity. Outbreak weeks made up 0.7% to 22.9% of rows. On outbreak weeks, the excess gain had a CI above zero with Peru and Taiwan held out (+3.62% and +1.32%). It had a CI below zero in Brazil's held-out states and held-out regions and in Colombia's held-out regions (Table S32). When districts were weighted by population or by past cases, only Colombia held out had a CI above zero (+0.49% with case weights). Colombian losses grew under population weighting, to −1.34% with held-out regions. With Peru held out the gain turned negative under both weightings (Table S33). Adjusting the forecast quantiles with earlier test years only (causal recalibration) changed the excess gain by 0.08 percentage points or less in most comparisons. The exceptions were Brazil held out (−0.68% to −0.47% with Mondrian shifts), Peru, future years (−0.82% to −0.61%), Peru held out (+0.93% to 0.00%) and Taiwan held out (−18.29% to −16.71%). Recalibration raised coverage on weeks with cases mainly in Peru (Table S34). Some subgroups were computed before the origin-year filter and the Espírito Santo exclusion. In them, no subgroup defined by population, surveillance history or urban growth showed a significant gain, except with Peru held out. Where losses occurred, they were largest in districts with no reported case in 2014 to 2017 (Table S35).

**Table S32. Excess gain on outbreak and non-outbreak weeks.**

| Comparison | Outbreak share of rows | Outbreak weeks: % (95% CI); p | Other weeks: % (95% CI); p | Difference, p |
|---|---|---|---|---|
| Brazil, future years | 8.2% | +0.08 (−0.77 to 0.92); 0.85 | +0.01 (−0.19 to 0.20); 0.94 | 0.85 |
| Brazil, held-out states | 8.2% | −0.98 (−1.96 to −0.01); 0.048 | +0.15 (−0.06 to 0.36); 0.16 | 0.041 |
| Brazil, held-out regions | 8.2% | −0.98 (−1.73 to −0.23); 0.013 | +0.13 (−0.19 to 0.44); 0.41 | 0.011 |
| Brazil, held-out districts | 8.2% | +0.01 (−0.83 to 0.86); 0.97 | +0.03 (−0.16 to 0.23); 0.73 | 0.99 |
| Brazil, held-out country | 8.2% | +0.50 (−0.07 to 1.08); 0.081 | −0.81 (−1.18 to −0.45); <0.001 | 0.030 |
| Colombia, future years | 2.3% | −0.70 (−2.33 to 0.94); 0.37 | −0.42 (−0.63 to −0.21); 0.0012 | 0.44 |
| Colombia, held-out states | 2.3% | −1.12 (−2.88 to 0.64); 0.19 | −0.48 (−0.68 to −0.28); <0.001 | 0.23 |
| Colombia, held-out regions | 2.3% | −1.86 (−3.31 to −0.42); 0.016 | −0.54 (−1.02 to −0.07); 0.028 | 0.023 |
| Colombia, held-out districts | 2.3% | −0.96 (−2.80 to 0.88); 0.27 | −0.42 (−0.65 to −0.19); 0.0018 | 0.32 |
| Colombia, held-out country | 2.3% | −0.34 (−1.47 to 0.79); 0.52 | +0.30 (−0.17 to 0.77); 0.19 | 0.46 |
| Peru, future years | 22.9% | −1.64 (−3.30 to 0.03); 0.053 | −0.69 (−1.23 to −0.14); 0.017 | 0.089 |
| Peru, held-out country | 22.9% | +3.62 (1.36 to 5.89); 0.0043 | −0.96 (−1.57 to −0.36); 0.0045 | 0.0017 |
| Taiwan, future years | 0.7% | −0.46 (−1.16 to 0.24); 0.14 | −1.05 (−2.25 to 0.16); 0.084 | 0.15 |
| Taiwan, held-out country | 0.7% | +1.32 (0.21 to 2.44); 0.030 | −22.45 (−36.24 to −8.67); 0.0033 | 0.024 |

*Note:* An outbreak week has cases at least max(5, the district's 90th percentile of weekly cases over 2013 to 2019); this window includes the training years 2018 to 2019 but no test year. Each estimate is a percentage of that stratum's own reference WIS. The last column tests the weekly series of the difference between outbreak and non-outbreak weeks.

**Table S33. Excess gain weighted by population or by past cases.**

| Comparison | Unweighted: % (95% CI); p | Population-weighted: % (95% CI); p | Weighted by 2013 to 2017 mean weekly cases + 1: % (95% CI); p |
|---|---|---|---|
| Brazil, future years | +0.23 (−0.33 to 0.79); 0.39 | +0.03 (−0.40 to 0.46); 0.88 | −0.13 (−0.53 to 0.27); 0.51 |
| Brazil, held-out states | 0.00 (−0.34 to 0.34); 1.0 | −0.15 (−0.53 to 0.22); 0.40 | −0.21 (−0.64 to 0.22); 0.32 |
| Brazil, held-out regions | −0.16 (−0.34 to 0.03); 0.090 | −0.42 (−0.90 to 0.07); 0.086 | −0.58 (−0.99 to −0.16); 0.010 |
| Brazil, held-out districts | +0.20 (−0.32 to 0.71); 0.43 | −0.16 (−0.58 to 0.25); 0.42 | −0.11 (−0.51 to 0.28); 0.55 |
| Brazil, held-out country | −0.68 (−0.94 to −0.42); <0.001 | −0.68 (−1.18 to −0.18); 0.011 | −0.65 (−1.15 to −0.14); 0.015 |
| Colombia, future years | −0.42 (−0.64 to −0.21); 0.0012 | −1.02 (−1.52 to −0.52); <0.001 | −0.72 (−1.14 to −0.30); 0.0032 |
| Colombia, held-out states | −0.52 (−0.74 to −0.30); <0.001 | −0.68 (−1.34 to −0.02); 0.043 | −0.52 (−1.04 to −0.001); 0.050 |
| Colombia, held-out regions | −0.68 (−1.08 to −0.29); 0.0030 | −1.34 (−2.20 to −0.48); 0.0056 | −0.48 (−0.90 to −0.06); 0.028 |
| Colombia, held-out districts | −0.45 (−0.68 to −0.21); 0.0017 | −0.86 (−1.59 to −0.13); 0.025 | −0.75 (−1.46 to −0.03); 0.042 |
| Colombia, held-out country | +0.22 (−0.19 to 0.63); 0.27 | +0.51 (−0.02 to 1.04); 0.058 | +0.49 (0.07 to 0.92); 0.026 |
| Peru, future years | −0.82 (−1.49 to −0.14); 0.022 | −1.41 (−3.50 to 0.69); 0.17 | −4.19 (−6.83 to −1.55); 0.0045 |
| Peru, held-out country | +0.93 (−0.16 to 2.02); 0.088 | −1.37 (−3.32 to 0.58); 0.15 | −0.89 (−2.36 to 0.59); 0.22 |

*Table S33 (continued)*

| Comparison | Unweighted: % (95% CI); p | Population-weighted: % (95% CI); p | Weighted by 2013 to 2017 mean weekly cases + 1: % (95% CI); p |
|---|---|---|---|
| Taiwan, future years | −0.71 (−1.40 to −0.03); 0.042 | −0.92 (−1.80 to −0.03); 0.044 | −1.07 (−1.98 to −0.16); 0.025 |
| Taiwan, held-out country | −18.29 (−29.80 to −6.78); 0.0039 | −15.19 (−25.04 to −5.34); 0.0048 | −9.43 (−15.65 to −3.21); 0.0054 |

*Note:* Within each horizon and week, rows are weighted by the district's population in the target year, or by its mean weekly cases in 2013 to 2017 plus 1. Weeks are then averaged as in the primary estimator.

**Table S34. Excess gain after causal recalibration of the quantiles.**

| Comparison | Raw: %; p | Global shift: %; p | Mondrian shift: %; p | Mondrian, from second test year: %; p | 80% coverage of L4 + C*, cases > 0: raw → Mondrian |
|---|---|---|---|---|---|
| Brazil, future years | +0.23; 0.39 | +0.23; 0.39 | +0.24; 0.36 | +0.33; 0.29 | 0.66 → 0.69 |
| Brazil, held-out states | 0.00; 1.0 | 0.00; 1.0 | +0.06; 0.76 | +0.15; 0.51 | 0.66 → 0.69 |
| Brazil, held-out regions | −0.16; 0.090 | −0.16; 0.090 | −0.08; 0.51 | −0.06; 0.69 | 0.66 → 0.69 |
| Brazil, held-out districts | +0.20; 0.43 | +0.20; 0.43 | +0.22; 0.39 | +0.29; 0.33 | 0.66 → 0.69 |
| Brazil, held-out country | −0.68; <0.001 | −0.60; <0.001 | −0.47; <0.001 | −0.53; <0.001 | 0.64 → 0.67 |
| Colombia, future years | −0.42; 0.0012 | −0.42; 0.0012 | −0.43; 0.0056 | −0.29; 0.11 | 0.71 → 0.70 |
| Colombia, held-out states | −0.52; <0.001 | −0.52; <0.001 | −0.53; 0.0018 | −0.47; 0.032 | 0.70 → 0.70 |
| Colombia, held-out regions | −0.68; 0.0030 | −0.68; 0.0031 | −0.64; 0.0037 | −0.64; 0.0026 | 0.70 → 0.70 |
| Colombia, held-out districts | −0.45; 0.0017 | −0.45; 0.0017 | −0.46; 0.0070 | −0.32; 0.12 | 0.71 → 0.70 |
| Colombia, held-out country | +0.22; 0.27 | +0.22; 0.27 | +0.25; 0.20 | −0.10; 0.26 | 0.75 → 0.73 |
| Peru, future years | −0.82; 0.022 | −0.79; 0.021 | −0.61; 0.12 | −0.33; 0.45 | 0.67 → 0.76 |
| Peru, held-out country | +0.93; 0.088 | +0.76; 0.16 | 0.00; 0.99 | −0.36; 0.35 | 0.67 → 0.76 |
| Taiwan, future years | −0.71; 0.042 | −0.71; 0.042 | −0.71; 0.042 | −0.50; 0.13 | 0.23 → 0.23 |
| Taiwan, held-out country | −18.29; 0.0039 | −16.74; 0.0052 | −16.71; 0.0052 | −14.76; 0.019 | 0.61 → 0.59 |

*Note:* Separately for L3 + C* and L4 + C*, and per horizon and seed, additive shifts of each quantile on the log(1 + cases) scale are fitted only on earlier test years. A global shift uses one shift per quantile; a Mondrian shift uses one shift per group of forecasts with a similar median. The first test year has no earlier year and is left unadjusted in the first three result columns; the fourth drops it.

**Table S35. Excess gain by surveillance history and population (exploratory; earlier scoring).**

| Comparison | No case 2014 to 2017 | Cases in 1 to 2 years | Cases in 3 to 4 years | Population, lowest third | Middle third | Highest third |
|---|---|---|---|---|---|---|
| Brazil, future years | +0.55 (n = 507) | +0.58 | +0.21 | +0.27 | +0.33 | +0.21 |
| Brazil, held-out states | +0.04 (n = 507) | +0.15 | 0.00 | +0.02 | +0.05 | −0.01 |
| Brazil, held-out regions | −1.76* (n = 507) | −0.45* | −0.03 | −0.17 | −0.08 | −0.14 |
| Brazil, held-out country | −2.47* (n = 507) | −1.29* | −0.47* | −0.77* | −0.61* | −0.58* |
| Colombia, future years | −1.14* (n = 65) | −0.66* | −0.38* | −0.82* | −0.42* | −0.28* |
| Colombia, held-out states | −19.74* (n = 65) | −2.74* | −0.39* | −1.27* | −0.52* | −0.27 |
| Colombia, held-out regions | −40.70* (n = 65) | −5.52* | −0.49* | −2.36* | −0.84* | −0.21 |
| Colombia, held-out districts | −7.14* (n = 65) | −2.31* | −0.38* | −1.19* | −0.50* | −0.24 |
| Colombia, held-out country | −11.63* (n = 65) | −0.81 | +0.22 | −0.63* | +0.15 | +0.39 |
| Peru, future years | −0.31 (n = 34) | +0.50 | −0.81* | 0.00 | −0.49 | −1.13* |
| Peru, held-out country | −0.75 (n = 34) | −0.35 | +1.35* | +0.75 | +2.18* | −0.09 |
| Taiwan, future years | −0.50 (n = 42) | −0.56 | −0.24 | +0.12 | −0.09 | −0.66 |
| Taiwan, held-out country | −28.83* (n = 42) | −13.79* | −5.88* | −16.71* | −11.99* | −9.66* |

*Note:* Excess gain as a percentage of the stratum's own reference WIS; an asterisk marks unadjusted p < 0.05; n is the number of districts with no reported case in 2014 to 2017. History is the number of years in 2014 to 2017 with at least one reported case. Population thirds are formed within each comparison and test year from the population in the year before the test year. These strata were computed before the changes made after review and were not recomputed. They use all rows without the origin-year filter, the earlier exclusions with Espírito Santo included, and Taiwan without 2021, so they differ slightly from the primary estimates. Brazil's held-out-districts comparison had not been run when they were computed and is not shown. Thirds of urban growth (change in built-up share, 2010 to 2015) are in the source file and show no consistent pattern.

#### E. Location controls, sensitivity analyses and transfer to Taiwan

**Table S36. Control and excess gains with each fixed location control in the future-years split.**

| Country | Control | Control gain with this control, % (95% CI); p | Excess gain with this control, % (95% CI); p |
|---|---|---|---|
| Brazil | C1 | +0.02 (−0.15 to 0.20); 0.77 | +0.25 (−0.32 to 0.81); 0.37 |
| Brazil | C2 | +0.14 (−0.16 to 0.43); 0.34 | +0.02 (−0.25 to 0.29); 0.88 |
| Brazil | C3 | −0.04 (−0.08 to −0.003); 0.038 | +0.28 (−0.27 to 0.82); 0.30 |
| Brazil | C6 | −2.11 (−2.68 to −1.54); <0.001 | +0.30 (−0.12 to 0.71); 0.15 |
| Colombia | C1 | −0.07 (−0.34 to 0.20); 0.60 | −0.39 (−0.58 to −0.19); 0.0011 |
| Colombia | C2 | −0.12 (−0.21 to −0.03); 0.016 | −0.34 (−0.59 to −0.10); 0.010 |
| Colombia | C3 | −0.12 (−0.20 to −0.04); 0.0053 | −0.33 (−0.49 to −0.16); 0.0011 |
| Colombia | C6 | −2.37 (−2.80 to −1.94); <0.001 | −0.13 (−0.37 to 0.12); 0.27 |
| Peru | C1 | −0.26 (−1.05 to 0.53); 0.49 | −0.70 (−1.41 to 0.02); 0.056 |
| Peru | C2 | −0.31 (−0.73 to 0.11); 0.13 | −0.86 (−1.57 to −0.14); 0.022 |
| Peru | C3 | −0.76 (−1.48 to −0.04); 0.040 | −0.37 (−0.80 to 0.05); 0.081 |
| Peru | C6 | −6.28 (−9.87 to −2.69); 0.0023 | −0.29 (−1.53 to 0.96); 0.63 |
| Taiwan | C1 | −0.63 (−1.68 to 0.43); 0.22 | −0.60 (−1.20 to −0.003); 0.049 |
| Taiwan | C2 | −0.36 (−0.76 to 0.03); 0.070 | −0.94 (−2.03 to 0.15); 0.087 |
| Taiwan | C3 | −0.58 (−1.29 to 0.12); 0.097 | −0.68 (−1.45 to 0.09); 0.079 |
| Taiwan | C6 | −2.29 (−5.59 to 1.02); 0.16 | −0.52 (−1.16 to 0.13); 0.11 |
| Brazil | none (embedding gain, L3 vs L4) | – | +0.29 (−0.27 to 0.84); 0.29 |
| Colombia | none (embedding gain, L3 vs L4) | – | −0.36 (−0.58 to −0.13); 0.0052 |
| Peru | none (embedding gain, L3 vs L4) | – | −0.94 (−1.70 to −0.18); 0.020 |
| Taiwan | none (embedding gain, L3 vs L4) | – | −1.06 (−1.96 to −0.16); 0.024 |

*Note:* Each row replaces the selected control by one fixed control. C1 is average past cases, the district's mean log(1 + cases) in the years before 2018. C2 is coordinates with spherical harmonics. C3 is the AlphaEarth series of a fixed other district. C6 is a district label, entered as a categorical input. The control gain compares L3 with L3 + Ck, and the excess gain compares L3 + Ck with L4 + Ck. The last four rows give the embedding gain without a control. Seed 0 only, so the C1 rows differ slightly from the three-seed values in Table 2. The panel is scored on the same primary rows as Table 2. Taiwan includes 2021, giving 141,645 rows; the values without 2021 are in Table S53. No adjustment for multiplicity.

None of the fixed controls turned the embeddings into a gain, and none of the 16 excess-gain CIs lies above zero. In Brazil the excess gain was positive with every control (+0.02% to +0.30%) but never significant. With coordinates (C2) almost none of it remained (+0.02%). The district label (C6) made the reference model worse in every country. Its control gain was −2.11% in Brazil, −2.37% in Colombia, −6.28% in Peru and −2.29% in Taiwan, the last not significant.

**Table S37. Sensitivity analyses of the embedding, control and excess gains.**

| Variant | Gain | Variant: % (95% CI); p | Primary comparison, % |
|---|---|---|---|
| Brazil, future years, yearly inputs keyed to the origin year (strictly causal) | embedding | +0.26 (−0.29 to 0.82); 0.33 | +0.26 (seed 0: +0.29) |
| Brazil, future years, 2-week reporting delay | embedding | +0.60 (−0.56 to 1.76); 0.29 | +0.26 (seed 0: +0.29) |
| Brazil, future years, C3 in place of C* | embedding | +0.26 (−0.31 to 0.82); 0.35 | +0.26 |
| Brazil, future years, C3 in place of C* | control | −0.04 (−0.08 to −0.003); 0.038 | +0.03 (seed 0: +0.02) |
| Brazil, future years, C3 in place of C* | excess | +0.28 (−0.27 to 0.82); 0.30 | +0.23 (seed 0: +0.25) |
| Brazil, future years, C6 in place of C* | embedding | +0.26 (−0.31 to 0.82); 0.35 | +0.26 |
| Brazil, future years, C6 in place of C* | control | −2.11 (−2.68 to −1.54); <0.001 | +0.03 (seed 0: +0.02) |
| Brazil, future years, C6 in place of C* | excess | +0.30 (−0.12 to 0.71); 0.15 | +0.23 (seed 0: +0.25) |
| Brazil, future years, per-level tuning (31 leaves; original protocol) | embedding | +0.21 (−0.05 to 0.47); 0.10 | +0.26 |
| Brazil, future years, per-level tuning (31 leaves; original protocol) | control | −0.06 (−0.30 to 0.18); 0.62 | +0.03 |
| Brazil, future years, per-level tuning (31 leaves; original protocol) | excess | +0.25 (−0.07 to 0.56); 0.11 | +0.23 |
| Taiwan, future years, Brazil's parameters and C* (no 2021) | embedding | −0.87 (−1.78 to 0.03); 0.057 | −0.95 |
| Taiwan, future years, Brazil's parameters and C* (no 2021) | control | −0.71 (−1.67 to 0.26); 0.14 | −0.41 |
| Taiwan, future years, Brazil's parameters and C* (no 2021) | excess | −0.33 (−0.89 to 0.23); 0.22 | −0.71 |
| Taiwan, future years without 2021 | embedding | −0.94 (−1.85 to −0.03); 0.043 | −0.95 |
| Taiwan, future years without 2021 | control | −0.63 (−1.69 to 0.44); 0.23 | −0.41 |
| Taiwan, future years without 2021 | excess | −0.44 (−1.01 to 0.14); 0.12 | −0.71 |
| Taiwan, held-out country without 2021 | embedding | −14.68 (−20.24 to −9.11); <0.001 | −21.60 |
| Taiwan, held-out country without 2021 | control | −2.31 (−5.52 to 0.90); 0.14 | −3.36 |
| Taiwan, held-out country without 2021 | excess | −11.16 (−15.66 to −6.65); <0.001 | −18.29 |
| Brazil, future years, without case history (reference N3; C* = C2) | embedding | +2.54 (−0.82 to 5.89); 0.13 | +0.26 |
| Brazil, future years, without case history (reference N3; C* = C2) | control | +3.30 (1.68 to 4.93); <0.001 | +0.03 |
| Brazil, future years, without case history (reference N3; C* = C2) | excess | −0.43 (−2.06 to 1.19); 0.58 | +0.23 |
| Brazil, held-out states, without case history (reference N3; C* = C2) | embedding | +1.52 (−1.27 to 4.32); 0.27 | −0.02 |
| Brazil, held-out states, without case history (reference N3; C* = C2) | control | +0.88 (−0.48 to 2.23); 0.19 | −0.05 |
| Brazil, held-out states, without case history (reference N3; C* = C2) | excess | −0.01 (−1.28 to 1.27); 0.99 | 0.00 |

*Note:* The last column repeats the matching primary comparison. Per-level tuning: L3 and L4 tuned separately on the 2019 grid, giving 31 leaves as in the original protocol, with the earlier control selection. Taiwan, future years, with Brazil's configuration and selected control excludes 2021. Taiwan without 2021 is the set of test years used before review. The runs with strictly causal yearly inputs and with a two-week reporting delay refit L3 and L4 only, so they estimate only the embedding gain. Because they use seed 0, they are compared with the seed-0 embedding gain of Brazil, future years, +0.29% (95% CI −0.27 to 0.84; Table S36), rather than with the three-seed +0.26%. Rows without case history use N3 as the reference and coordinates (C2) as the selected control. The C3 and C6 rows replace the selected control by that fixed control (seed 0). Their embedding gain equals the primary one by construction, and their control and excess gains are compared with the seed-0 values given in brackets.

**Table S38. Forecast error in Taiwan's 2021 test year.**

| Comparison | L3 | L4 | L3 + C* | L4 + C* | Excess gain (absolute) | p |
|---|---|---|---|---|---|---|
| Taiwan, future years | 0.00031 | 0.00050 | 0.00031 | 0.00058 | −0.00026 | 0.077 |
| Taiwan, held-out country | 0.00394 | 0.01171 | 0.00512 | 0.01193 | −0.00665 | 0.091 |

*Note:* Mean WIS on log(1 + cases) on the 2021 rows, where every target is zero because Taiwan had no local cases in 2021. Seed 0. The last two columns give the absolute excess gain and its unadjusted p value. Percentages are not shown, because the reference WIS is close to zero.

**Table S39. Correlation between a held-out district's gain and its embedding similarity to the training districts.**

| Comparison | Districts | Spearman ρ, excess gain vs similarity (naive p) | Spearman ρ, embedding gain vs similarity (naive p) |
|---|---|---|---|
| Brazil, held-out states | 5,439 | +0.04 (0.0018) | +0.07 (<0.001) |
| Brazil, held-out regions | 5,439 | +0.12 (<0.001) | +0.07 (<0.001) |
| Brazil, held-out districts | 5,439 | +0.09 (<0.001) | +0.10 (<0.001) |
| Brazil, held-out country | 5,439 | +0.09 (<0.001) | +0.24 (<0.001) |
| Colombia, held-out states | 1,070 | +0.01 (0.70) | −0.01 (0.86) |
| Colombia, held-out regions | 1,070 | −0.04 (0.17) | −0.02 (0.49) |
| Colombia, held-out districts | 1,070 | +0.10 (0.0010) | +0.07 (0.022) |
| Colombia, held-out country | 1,070 | +0.15 (<0.001) | +0.02 (0.50) |
| Peru, held-out country | 115 | +0.52 (<0.001) | +0.48 (<0.001) |
| Taiwan, held-out country | 285 | −0.25 (<0.001) | −0.22 (<0.001) |

*Note:* Similarity is the cosine similarity between a held-out district's rescaled AlphaEarth vector for year y − 1 and that of the most similar training district. The p values treat districts as independent, which they are not, so they are too small. Exploratory, unadjusted.

**Table S40. Excess gain by similarity quintile in the held-out-country split.**

| Comparison | Similarity quintile (maximum cosine) | Districts | Reference WIS (L3) | Excess gain, log-WIS × $10^{-4}$ (95% CI) | Districts harmed |
|---|---|---|---|---|---|
| Brazil, held-out country | Q1 (0.43–0.67) | 1088 | 0.1035 | −15.6 (−16.4 to −14.8) | 92% |
| Brazil, held-out country | Q2 (0.67–0.72) | 1088 | 0.1521 | −13.2 (−14.0 to −12.3) | 84% |
| Brazil, held-out country | Q3 (0.72–0.76) | 1087 | 0.1781 | −13.6 (−14.4 to −12.7) | 84% |
| Brazil, held-out country | Q4 (0.76–0.80) | 1088 | 0.2429 | −12.2 (−13.1 to −11.2) | 81% |
| Brazil, held-out country | Q5 (0.80–0.99) | 1088 | 0.2802 | −11.7 (−12.8 to −10.6) | 77% |
| Colombia, held-out country | Q1 (0.53–0.78) | 214 | 0.1015 | −1.6 (−3.6 to 0.5) | 73% |
| Colombia, held-out country | Q2 (0.78–0.81) | 214 | 0.1307 | +1.7 (−0.4 to 3.8) | 58% |
| Colombia, held-out country | Q3 (0.81–0.84) | 214 | 0.1305 | +4.1 (2.1 to 6.1) | 50% |
| Colombia, held-out country | Q4 (0.84–0.86) | 214 | 0.1363 | +6.6 (4.2 to 9.1) | 47% |
| Colombia, held-out country | Q5 (0.86–0.99) | 214 | 0.1201 | +3.4 (1.4 to 5.4) | 57% |
| Peru, held-out country | Q1 (0.62–0.76) | 23 | 0.2211 | −33.1 (−53.0 to −13.2) | 83% |
| Peru, held-out country | Q2 (0.76–0.84) | 23 | 0.2562 | +7.1 (−28.0 to 42.3) | 61% |
| Peru, held-out country | Q3 (0.85–0.88) | 23 | 0.2336 | +38.1 (2.4 to 73.9) | 48% |
| Peru, held-out country | Q4 (0.88–0.92) | 23 | 0.1768 | +36.6 (9.0 to 64.2) | 39% |
| Peru, held-out country | Q5 (0.93–0.99) | 23 | 0.3040 | +64.3 (33.0 to 95.6) | 17% |
| Taiwan, held-out country | Q1 (0.49–0.74) | 57 | 0.0054 | −30.8 (−34.7 to −26.9) | 100% |
| Taiwan, held-out country | Q2 (0.74–0.76) | 57 | 0.0147 | −40.8 (−46.0 to −35.6) | 100% |
| Taiwan, held-out country | Q3 (0.76–0.78) | 57 | 0.0171 | −35.5 (−39.4 to −31.6) | 100% |
| Taiwan, held-out country | Q4 (0.78–0.80) | 57 | 0.0182 | −35.3 (−38.4 to −32.1) | 100% |
| Taiwan, held-out country | Q5 (0.80–0.88) | 57 | 0.0473 | −46.2 (−50.0 to −42.3) | 100% |

*Note:* Similarity as in Table S39. The excess gain is absolute (log-WIS), and the reference WIS of each quintile is week-weighted. "Districts harmed" is the share of districts with a negative excess gain. For comparison, the quintile means of similarity are 0.95 to 0.99 in Brazil's and 0.93 to 0.98 in Colombia's held-out-states splits. Exploratory, unadjusted.

With Taiwan held out, the loss did not shrink as similarity rose (−30.8 to −46.2 × $10^{-4}$ log-WIS per quintile), and all 285 districts had a loss. With Peru held out the gain rose with similarity, from −33.1 × $10^{-4}$ log-WIS in the least similar quintile to +64.3 × $10^{-4}$ in the most similar (ρ = +0.52).

#### F. Forecasts without case history

**Table S41. Forecast error without case history, Brazil, future years and held-out states (row mean).**

| Model | Future years: horizon 1 | Future years: horizon 4 | Future years: horizons 1 and 4 | Future years: 80% coverage, all rows / cases > 0 (pooled) | Held-out states: horizons 1 and 4 | Held-out states: 80% coverage, all rows / cases > 0 |
|---|---|---|---|---|---|---|
| N3: season, climate, remote sensing and population | 0.3520 | 0.3533 | 0.3526 | 0.85 / 0.43 | 0.3613 | 0.84 / 0.42 |
| N3 + C1: static history (mean log(1 + cases), 2014 to 2017) | 0.3473 | 0.3483 | 0.3478 | 0.84 / 0.43 | 0.3622 | 0.84 / 0.42 |
| N4: N3 + AlphaEarth | 0.3437 | 0.3437 | 0.3437 | 0.83 / 0.41 | 0.3557 | 0.82 / 0.40 |
| N4 + C1 | 0.3404 | 0.3413 | 0.3409 | 0.83 / 0.41 | 0.3555 | 0.82 / 0.40 |
| N3 + C2: coordinates (C*) | 0.3404 | 0.3419 | 0.3411 | 0.84 / 0.43 | 0.3582 | 0.82 / 0.42 |
| N4 + C2 | 0.3423 | 0.3429 | 0.3426 | 0.84 / 0.42 | 0.3581 | 0.82 / 0.41 |
| With case history, L3 / L4 (same rows) | 0.1419 / 0.1418 | 0.1959 / 0.1952 | 0.1681 / 0.1677 | 0.90 / 0.67 (L4) | 0.1689 / 0.1690 | 0.90 / 0.66 (L4 + C*) |

*Note:* Mean WIS over rows (lower is better). Seed 0. All 21 case-derived inputs are removed. Static history (C1) is a separate step and not a candidate for the selected control, which is coordinates (C2) in both comparisons. The row with case history gives L3 and L4 on the same rows for reference. For Brazil, future years, it uses three seeds and the coverage of L4; for held-out states it uses seed 0 and the coverage of L4 + C*.

**Table S42. Embedding, control and excess gains without case history, Brazil, future years and held-out states.**

| Comparison and horizon | Reference WIS (N3) | Embedding gain, % (95% CI); p | Control gain (C2), % (95% CI); p | Excess gain, % (95% CI); p | Minimum detectable excess gain, % |
|---|---|---|---|---|---|
| Future years, horizon 1 | 0.3520 | +2.35 (−0.83 to 5.53); 0.14 | +3.28 (1.60 to 4.96); <0.001 | −0.53 (−1.99 to 0.94); 0.46 | 2.07 |
| Future years, horizon 4 | 0.3533 | +2.72 (−0.92 to 6.37); 0.13 | +3.22 (1.61 to 4.84); <0.001 | −0.29 (−2.13 to 1.55); 0.74 | 2.59 |
| Future years, horizons 1 and 4 | 0.3520 | +2.54 (−0.82 to 5.89); 0.13 | +3.30 (1.68 to 4.93); <0.001 | −0.43 (−2.06 to 1.19); 0.58 | 2.29 |
| Held-out states, horizon 1 | 0.3607 | +1.36 (−1.36 to 4.07); 0.31 | +0.88 (−0.62 to 2.39); 0.23 | −0.06 (−1.18 to 1.06); 0.91 | 1.58 |
| Held-out states, horizon 4 | 0.3619 | +1.73 (−1.23 to 4.69); 0.23 | +0.82 (−0.34 to 1.97); 0.15 | +0.13 (−1.40 to 1.65); 0.86 | 2.14 |
| Held-out states, horizons 1 and 4 | 0.3606 | +1.52 (−1.27 to 4.32); 0.27 | +0.88 (−0.48 to 2.23); 0.19 | −0.01 (−1.28 to 1.27); 0.99 | 1.80 |

*Note:* N3 is L3 and N4 is L4 without the 21 case-derived inputs. The selected control is coordinates (C2) in both comparisons. Gains are percentages of the WIS of N3. Horizon rows give the mean over rows; pooled rows weight target weeks equally, as in the gains. The minimum detectable effect is at 80% power. Seed 0. The identity share, the control gain as a share of the embedding gain, cannot be estimated in either comparison, because the CI of the embedding gain includes zero. Static-history step (C1, pooled horizons): for future years, the gain from N3 to N3 + C1 was +1.38% (95% CI −0.09 to 2.85; p = 0.064) and from N3 + C1 to N4 + C1 +1.95% (−1.77 to 5.66; p = 0.28). For held-out states the same gains were −0.24% (−0.82 to 0.34; p = 0.39) and +1.85% (−1.09 to 4.79; p = 0.20).

In held-out Brazilian states, no estimate without case history was significant. The embedding gain (+1.52%) was of the same order as the gain from coordinates (+0.88%). Beyond coordinates the embeddings added nothing (−0.01%, 95% CI −1.28 to 1.27). The significant gain from coordinates in the future-years comparison (+3.30%) was therefore not detected in states that were not used for fitting. We had pre-specified the pattern that would point to identity: an embedding gain above zero together with an excess gain equivalent to zero.

It was not met in either comparison, because neither embedding gain was significant and neither excess gain was equivalent within ±1%.

#### G. Post hoc analyses added after the third review

The analyses in this section were specified in response to reviewers' reports. By then 13 of the 14 comparisons of the primary family had been read, but not the result for Brazil's held-out districts. Each analysis was specified before any of it was scored (deviation rows 39 to 53, Supplementary Methods Table S14; specification commit 8001a92). Unless stated otherwise, they reuse the primary scoring: week-weighted, origin-year rows, primary exclusions, horizons 1 and 4. The final run repeated Tables S43 to S48 on all 14 comparisons. It used the primary estimates of Table 2 as reference values, which are the mean of seeds 0 to 2 where three seeds exist (row 53), with Holm adjustment over 14 comparisons. Tables S49 to S51 and S53 to S55 use seed 0. None of these analyses enters the Holm family of Table 2, and all are exploratory.

##### G1. As planned and as reported

**Table S43. Excess gain under the protocol as planned and as reported.**
Each column changes one reporting choice back to its planned or earlier form and keeps the others at their reported values. Entries are the excess gain as % of the reference WIS, followed by p.

| Comparison | C* | Reported (Table 2) | All rows (row 24) | Exclusions v1 (row 34) | B = 4 variance | Seed 0 only (before row 53) | Other planned settings |
|---|---|---|---|---|---|---|---|
| Brazil, future years (S1) | C1 | +0.23; 0.39 | +0.26; 0.39 | +0.23; 0.40 | +0.23; 0.45 | +0.25; 0.37 | Horizons 1 to 4 +0.25; 0.38. Per-level tuning +0.25; 0.11 |
| Brazil, held-out states (S2) | C1 | 0.00; 1.0 | +0.03; 0.89 | −0.01; 0.94 | 0.00; 1.0 | = reported | – |
| Brazil, held-out regions (S3) | C1 | −0.16; 0.090 | −0.10; 0.40 | **−0.19; 0.045** | −0.16; 0.26 | = reported | – |
| Brazil, held-out districts (S4) | C1 | +0.20; 0.43 | +0.23; 0.42 | +0.19; 0.44 | +0.20; 0.47 | = reported | – |
| Brazil, held-out country (S5) | C3 | −0.68; <0.001 | −0.69; <0.001 | −0.70; <0.001 | −0.68; 0.0073 | −0.60; <0.001 | rule v1 (C4): no valid run |
| Colombia, future years (S1) | C1 | −0.42; 0.0012 | −0.42; <0.001 | = reported | −0.42; 0.014 | −0.39; 0.0011 | Horizons 1 to 4 −0.41; 0.0024 |
| Colombia, held-out states (S2) | C1 | −0.52; <0.001 | −0.53; <0.001 | = reported | −0.52; 0.0016 | −0.47; <0.001 | – |
| Colombia, held-out regions (S3) | C1 | −0.68; 0.0030 | −0.71; 0.0025 | = reported | −0.68; 0.0093 | −0.65; 0.0033 | – |
| Colombia, held-out districts (S4) | C1 | −0.45; 0.0017 | −0.45; 0.0011 | = reported | −0.45; 0.015 | −0.43; 0.0028 | – |
| Colombia, held-out country (S5) | C2 | +0.22; 0.27 | +0.18; 0.33 | = reported | +0.22; 0.49 | = reported | rule v1: same control |
| Peru, future years (S1) | C1 | −0.82; 0.022 | −0.73; 0.046 | = reported | −0.82; 0.11 | −0.70; 0.056 | Horizons 1 to 4 −0.69; 0.047. **Per-level tuning +0.65; 0.24** |
| Peru, held-out country (S5) | C3 | +0.93; 0.088 | +0.99; 0.051 | = reported | **+0.93; 0.020** | +0.96; 0.079 | – |
| Taiwan, future years (S1) | C1 | −0.71; 0.042 | −0.66; 0.062 | = reported | −0.71; 0.037 | −0.60; 0.049 | Without 2021 −0.44; 0.12. Horizons 1 to 4 −0.46; 0.14. Per-level tuning −0.14; 0.23. Rule v1 (C3) −0.68; 0.079 |
| Taiwan, held-out country (S5) | C3 | −18.29; 0.0039 | −18.46; 0.0029 | = reported | −18.29; 0.034 | −17.30; 0.0017 | Without 2021 −11.16; <0.001 |

*Note:* "Reported" is the primary estimate of Table 2, the mean of seeds 0 to 2 where three seeds exist (row 53). The seed-0 column gives the estimate before seed averaging, which is the version the reviewers saw; "= reported" marks comparisons with seed 0 only.

Bold marks an entry whose conclusion differs from the reported one at the unadjusted 5% level. The unadjusted Taiwan future-years loss is discussed below. In the exclusions column, "= reported" means the estimate is identical, because the earlier exclusions differ from the primary ones only in Brazil (Espírito Santo). The control chosen by the first selection rule (rule v1) was the same as the one chosen by the final rule (rule v2) wherever it was recorded: Brazil and Peru, future years, and Colombia, held-out country. Those rule-v1 entries are therefore identical to the reported ones. It was not recorded for most held-out-place comparisons. Brazil held out chose C4 under rule v1, and those runs were discarded after the C4 fix (row 18). Taiwan's rule-v1 value with C3 is from the fixed control panel with 2021 (Table S36), seed 0.

The main conclusions hold under every planned setting. Brazil, future years, stays between +0.23% and +0.26%, and Brazil's held-out districts between +0.19% and +0.23%; neither is significant. Brazil's held-out states stay at zero. The losses with Brazil held out and in Colombia's four splits stay negative with p ≤ 0.015 under every planned setting, including the strict-variance check and seed 0 alone. The interpretation changes for a few estimates. In Peru, future years, the sign reverses under per-level tuning, from −0.82% to +0.65% (95% CI −0.49 to 1.79). This is the same effect of model size that first appeared in Brazil. With Peru held out, the gain becomes significant before adjustment under the strict-variance check (B = 4; +0.93%, 95% CI 0.25 to 1.62; p = 0.020). On all rows it is +0.99% (−0.003 to 1.98; p = 0.051). Neither is significant after Holm adjustment. Brazil's held-out regions show a loss that is significant before adjustment under the earlier exclusions, which keep Espírito Santo's false zeros (−0.19%, 95% CI −0.37 to −0.005; p = 0.045). The loss in Taiwan, future years, is significant only before adjustment (p = 0.042) and not after Holm adjustment. It loses even that significance under per-level tuning (−0.14%, p = 0.23), on all rows (p = 0.062), with horizons 1 to 4 (p = 0.14), without 2021 (p = 0.12) and with the rule-v1 control (p = 0.079). It keeps it only under the strict-variance check (B = 4; p = 0.037). Two further changes are covered elsewhere. Including Taiwan's 2021 (row 25) moved the estimate with Taiwan held out from −11.16% to −18.29%. The rule that allows "no control" removes the loss with Brazil held out (Table S45).

#### G2. Gains from known predictors along the ladder, as a scale for the margin

**Table S44. Gains from adding climate, remote sensing and population, and both, compared with the embedding gain, in the future-years split.**

| Comparison | Climate, L1 → L2 | Remote sensing and population, L2 → L3 | All non-case covariates, L1 → L3 | Embeddings, L3 → L4 | Excess gain | ±1% (±0.5%) as a multiple of the L1 → L3 gain |
|---|---|---|---|---|---|---|
| Brazil | **+0.57 (0.03 to 1.12); 0.041** | **+0.52 (0.18 to 0.86); 0.0053** | **+1.09 (0.44 to 1.74); 0.0026** | +0.26 (−0.31 to 0.82); 0.35 | +0.23 | 0.92 (0.46) |
| Colombia | +0.14 (−0.14 to 0.42); 0.29 | **+0.30 (0.03 to 0.57); 0.034** | +0.44 (−0.05 to 0.93); 0.072 | −0.38 (−0.61 to −0.16); 0.0035 | −0.42 | 2.26 (1.13) |
| Peru | +0.44 (−1.48 to 2.37); 0.63 | −0.24 (−0.99 to 0.51); 0.50 | +0.20 (−1.84 to 2.24); 0.84 | −0.91 (−1.62 to −0.20); 0.016 | −0.82 | 5.03 (2.51) |
| Taiwan (with 2021) | −1.10 (−2.60 to 0.40); 0.14 | −0.17 (−0.53 to 0.19); 0.34 | −1.27 (−2.92 to 0.38); 0.12 | −0.95 (−1.71 to −0.18); 0.018 | −0.71 | not defined (L1 → L3 gain negative) |

*Note:* Gains in % of WIS(L3) with 95% CI, and unadjusted p. All runs use one shared configuration per country. L3 and L4 average seeds 0 to 2 in every country. Bold marks a covariate step whose CI excludes zero. The excess gain is the primary estimate (Table 2). Without 2021 (seed 0), Taiwan's gains for the four steps were −0.93% (p = 0.21), +0.06% (p = 0.85), −0.87% (p = 0.28) and −0.94% (p = 0.043).

The design can detect a known set of predictors, but only in Brazil and, for one step, in Colombia. In Brazil, climate (+0.57%), remote sensing and population (+0.52%) and both together (+1.09%) each improved the forecast. In Colombia only remote sensing and population did (+0.30%, p = 0.034). In Peru and Taiwan no step was detectable. Taiwan's point estimates were negative, with −1.27% for all inputs other than case history (p = 0.12). The null embedding results in Peru and Taiwan therefore say little about whether useful information would have been detected. None of these p values is adjusted, and the two results with p between 0.03 and 0.05 would not stay significant after adjustment over the 16 tests. The ±1% margin equals 0.92 times the whole gain from Brazil's inputs other than case history, 2.26 times Colombia's and 5.03 times Peru's. A difference within ±1% therefore leaves room for an embedding effect as large as everything that climate, remote sensing and population add to case history. Brazil's excess gain (+0.23%) is about the size of its gain from climate.

#### G3. "No control" selection rule

**Table S45. Primary family when "no control" is allowed at the 2019 selection.**

| Comparison | C* under rule v2 → this rule | Estimand under this rule | % (95% CI) | p | Holm p (14 comparisons) | Primary excess gain (Holm p) | Control-gain CI below 0 |
|---|---|---|---|---|---|---|---|
| Brazil, future years (S1) | C1 → C1 | excess | +0.23 (−0.33 to 0.79) | 0.39 | 1.0 | +0.23 (1.0) | no |
| Brazil, held-out states (S2) | C1 → none | embedding | −0.02 (−0.34 to 0.30) | 0.88 | 1.0 | 0.00 (1.0) | no |
| Brazil, held-out regions (S3) | C1 → none | embedding | −0.13 (−0.35 to 0.09) | 0.21 | 1.0 | −0.16 (0.53) | no |
| Brazil, held-out districts (S4) | C1 → none | embedding | +0.22 (−0.32 to 0.76) | 0.40 | 1.0 | +0.20 (1.0) | no |
| Brazil, held-out country (S5) | C3 → none | embedding | −0.39 (−1.34 to 0.55) | 0.39 | 1.0 | **−0.68 (<0.001)** | no |
| Colombia, future years (S1) | C1 → none | embedding | **−0.38 (−0.61 to −0.16)** | 0.0035 | **0.035** | −0.42 (0.014) | no |
| Colombia, held-out states (S2) | C1 → none | embedding | **−0.60 (−0.81 to −0.39)** | <0.001 | **<0.001** | −0.52 (0.0034) | yes |
| Colombia, held-out regions (S3) | C1 → none | embedding | **−0.82 (−1.14 to −0.49)** | <0.001 | **0.0022** | −0.68 (0.030) | yes |
| Colombia, held-out districts (S4) | C1 → none | embedding | **−0.54 (−0.78 to −0.30)** | <0.001 | **0.0050** | −0.45 (0.019) | yes |
| Colombia, held-out country (S5) | C2 → C2 | excess | +0.22 (−0.19 to 0.63) | 0.27 | 1.0 | +0.22 (1.0) | yes |
| Peru, future years (S1) | C1 → none | embedding | −0.91 (−1.62 to −0.20) | 0.016 | 0.14 | −0.82 (0.18) | no |
| Peru, held-out country (S5) | C3 → none | embedding | +0.68 (−0.59 to 1.96) | 0.27 | 1.0 | +0.93 (0.53) | yes |
| Taiwan, future years (S1) | C1 → none | embedding | −0.95 (−1.71 to −0.18) | 0.018 | 0.15 | −0.71 (0.29) | no |
| Taiwan, held-out country (S5) | C3 → none | embedding | **−21.60 (−31.82 to −11.39)** | <0.001 | **0.0045** | −18.29 (0.035) | yes |

*Note:* Under this rule the selected control is "none" unless a candidate beat L3 at the 2019 origin by more than two standard errors, using the seed-noise standard error of the main selection rule. Comparisons without a control then contribute the embedding gain (L3 → L4). Only Brazil, future years (C1), and Colombia held out (C2) keep a control. Holm adjustment over the 14 comparisons. Bold marks Holm p < 0.05; in the primary column it is used only where the interpretation differs. The last column marks comparisons where the selected control on its own made L3 worse, that is, where the 95% CI of the control gain lies below zero. Estimates as in Table 2: seeds 0 to 2 where three seeds exist; the embedding gains of Peru and Taiwan held out use seed 0.

No comparison shows a gain under either rule. With Brazil held out, the loss appears only when the noise control of matching size (C3) is in the model. Without a control the embedding gain there is −0.39% (p = 0.39). The Colombian losses are of similar size or larger without a control and remain significant after Holm adjustment, so they do not depend on the choice of control. Under this rule the Holm-significant losses are

Colombia's four within-country splits and Taiwan held out. In six comparisons the selected control itself made the reference model worse: Colombia's held-out states, regions, districts and country, Peru held out and Taiwan held out.

#### G4. Trend over test years

**Table S46. Trend of the excess gain with the length of the training window, and Brazil's 2023 to 2024 test years.**

| Comparison | Slope, percentage points per extra training year | p, per-year SEs as given | p, SEs scaled for over-dispersion | Slope without 2020 and 2021 (scaled p) |
|---|---|---|---|---|
| Brazil, future years (S1) | +0.24 | 0.0034 | 0.12 | +0.34 (0.39) |
| Brazil, held-out states (S2) | +0.28 | <0.001 | 0.096 | +0.28 (0.31) |
| Brazil, held-out regions (S3) | +0.06 | 0.037 | 0.37 | +0.04 (0.69) |
| Brazil, held-out districts (S4) | +0.18 | 0.0080 | 0.20 | +0.32 (0.38) |
| Brazil, held-out country (S5) | +0.10 | 0.15 | 0.63 | +0.10 (0.48) |
| Colombia, future years (S1) | +0.22 | <0.001 | 0.18 | – |
| Colombia, held-out states (S2) | +0.16 | 0.019 | 0.26 | – |
| Colombia, held-out regions (S3) | +0.08 | 0.64 | 0.72 | – |
| Colombia, held-out districts (S4) | +0.26 | <0.001 | 0.16 | – |
| Colombia, held-out country (S5) | −0.12 | 0.26 | 0.79 | – |
| Peru, future years (S1) | +0.53 | 0.091 | 0.23 | – |
| Peru, held-out country (S5) | −0.39 | 0.38 | 0.47 | – |
| Taiwan, future years (S1) | +3.21 | 0.078 | 0.32 | −2.55 (0.62) |
| Taiwan, held-out country (S5) | +3.48 | 0.37 | 0.79 | −20.19 (0.54) |

| Brazil comparison, test years 2023 and 2024 only | Excess gain, % (95% CI); p (B = 4 p) | Embedding gain, % (95% CI); p |
|---|---|---|
| Future years (S1) | +0.69 (−0.40 to 1.78); 0.18 (0.23) | +0.68 (−0.45 to 1.81); 0.20 |
| Held-out states (S2) | +0.37 (−0.20 to 0.94); 0.17 (0.23) | +0.31 (−0.28 to 0.90); 0.26 |
| Held-out regions (S3) | +0.03 (−0.18 to 0.24); 0.74 (0.74) | +0.11 (−0.23 to 0.44); 0.48 |
| Held-out districts (S4) | +0.61 (−0.39 to 1.61); 0.20 (0.25) | +0.63 (−0.44 to 1.70); 0.21 |
| Held-out country (S5) | −0.54 (−0.81 to −0.28); 0.0015 (0.0089) | +0.12 (−1.63 to 1.87); 0.87 |

*Note:* Weighted least-squares regression of each comparison's yearly excess gain (Table S30) on the number of training years, with weights equal to the inverse squared standard error and 3 to 5 points per comparison. "Scaled" enlarges the standard errors to allow for year-to-year noise larger than the formula assumes (over-dispersion). The length of the training window cannot be separated from the COVID-19 years 2020 and 2021. Taiwan's yearly percentages are unstable, because the reference WIS is tiny in years with few cases. Post hoc. The yearly embedding gains behave in the same way.

With the yearly standard errors taken at face value, 7 of 14 comparisons had a positive slope with $p < 0.05$. Once the standard errors were scaled for over-dispersion, none did (smallest $p = 0.096$), and none did when 2020 and 2021 were dropped. The positive point estimates come mainly from losses in 2021, the COVID-affected year with the second-shortest training window. In Brazil's two most recent test years no comparison showed a significant gain (largest +0.69%, $p = 0.18$), and the loss with Brazil held out persisted (−0.54%, $p = 0.0015$). The training windows are short and COVID-affected, so the data cannot show whether the embeddings would help with longer training histories.

#### G5. Equivalence under other margins and stricter checks

**Table S47. Equivalence of the excess gain at ±0.5%, ±1% and ±2% under the main test, the strict-variance check (B = 4) and the surrogate-series check.**

| Comparison | 90% CI, EWC | 90% CI, B = 4 | Equivalent at ±0.5% | at ±1% | at ±2% | IAAFT size of the 5% test, EWC / B = 4 | IAAFT-calibrated 90% CI; equivalent at ±1% |
|---|---|---|---|---|---|---|---|
| Brazil, future years (S1) | −0.23 to 0.69 | −0.36 to 0.82 | – | a b c | a b c d | 18.4% / 12.5% | −0.62 to 1.08; no |
| Brazil, held-out states (S2) | −0.28 to 0.28 | −0.43 to 0.43 | a b c | a b c d | a b c d | 14.0% / 5.0% | −0.38 to 0.38; yes |
| Brazil, held-out regions (S3) | −0.31 to −0.01 | −0.41 to 0.10 | a b c | a b c d | a b c d | 15.2% / 5.0% | −0.37 to 0.05; yes |
| Brazil, held-out districts (S4) | −0.23 to 0.62 | −0.33 to 0.73 | – | a b c | a b c d | 14.9% / 9.9% | −0.48 to 0.88; yes |
| Brazil, held-out country (S5) | −0.89 to −0.47 | −0.97 to −0.39 | – | a c | a b c d | 6.5% / 3.5% | −0.91 to −0.45; yes |
| Colombia, future years (S1) | −0.60 to −0.25 | −0.64 to −0.21 | – | a b c d | a b c d | 8.7% / 4.1% | −0.63 to −0.22; yes |
| Colombia, held-out states (S2) | −0.70 to −0.34 | −0.67 to −0.38 | – | a b c d | a b c d | 1.1% / 2.0% | −0.66 to −0.39; yes |
| Colombia, held-out regions (S3) | −1.01 to −0.36 | −1.00 to −0.37 | – | c | a b c d | not run | – |
| Colombia, held-out districts (S4) | −0.64 to −0.25 | −0.68 to −0.21 | – | a b c d | a b c d | 7.4% / 3.6% | −0.66 to −0.23; yes |
| Colombia, held-out country (S5) | −0.12 to 0.55 | −0.39 to 0.83 | – | a b c | a b c d | 15.8% / 6.8% | −0.27 to 0.70; yes |
| Peru, future years (S1) | −1.37 to −0.26 | −1.68 to 0.05 | – | – | a b c d | not run | – |
| Peru, held-out country (S5) | 0.04 to 1.83 | 0.40 to 1.46 | – | – | a c d | not run | – |
| Taiwan, future years (S1) | −1.28 to −0.15 | −1.21 to −0.22 | – | – | a b c d | not run | – |
| Taiwan, held-out country (S5) | −27.77 to −8.81 | −30.57 to −6.01 | – | – | – | not run | – |
| Comparisons equivalent (a / b / c / d) | | | 2 / 2 / 2 / 0 | 9 / 8 / 10 / 5 | 13 / 12 / 13 / 13 | | 8 of 9 at ±1% |

*Note:* TOST on 90% CIs, % of reference WIS. Letters list the versions under which a comparison is equivalent: a, main test (EWC), unadjusted; b, EWC with Holm adjustment over 14 comparisons; c, strict-variance check (B = 4), unadjusted; d, B = 4 with Holm adjustment. The IAAFT columns come from the surrogate-series check, with 2,000 surrogates of each comparison's weekly loss-difference series under no effect. They were run only for the nine comparisons equivalent at ±1% under a, without Holm adjustment. Estimates as in Table 2 (seeds 0 to 2 where three seeds exist).

Equivalence depends on both the margin and the test. At ±1%, nine comparisons are equivalent under the main test without adjustment, and eight after Holm adjustment. Five are equivalent under the strict-variance check with Holm adjustment: Brazil's held-out states and regions, and Colombia's future years, held-out states and held-out districts. Three of these five are significantly negative, so "equivalent" there means a loss smaller than 1% rather than no effect. At ±0.5%, two comparisons are equivalent under the main test, with or without Holm adjustment, and none under the strict-variance check with Holm adjustment.

The surrogate-series check shows that the main test finds effects too often in five comparisons: Brazil's future years, held-out states, held-out regions and held-out districts, and Colombia held out. There, a nominal 5% test rejected 14% to 18% of the time. After this calibration, Brazil, future years, the comparison with the largest positive point estimate, is no longer equivalent at ±1% (calibrated 90% CI −0.62 to 1.08), so neither a gain nor equivalence is established there. In the comparisons with clear losses the rate is close to nominal or below it (1.1% to 8.7%).

#### G6. District-years with cases, and forecast width and coverage

**Table S48. District-years with cases, and forecast width and coverage on weeks with and without cases.**

**Panel A.** Excess and embedding gains on district-years with at least one reported case in that test year. The condition applies to the whole district-year, so weeks without cases in those years are kept.

| Comparison | Rows kept | Excess gain, % (95% CI); p | Primary excess gain | Embedding gain, % (95% CI); p |
|---|---|---|---|---|
| Colombia, future years (S1) | 71% | **−0.33 (−0.53 to −0.13); 0.0038** | −0.42 | **−0.28 (−0.49 to −0.07); 0.014** |
| Colombia, held-out states (S2) | 71% | **−0.33 (−0.52 to −0.13); 0.0040** | −0.52 | **−0.40 (−0.58 to −0.22); <0.001** |
| Colombia, held-out regions (S3) | 71% | −0.33 (−0.71 to 0.05); 0.083 | −0.68 | **−0.45 (−0.75 to −0.15); 0.0071** |
| Colombia, held-out districts (S4) | 71% | **−0.29 (−0.49 to −0.09); 0.0084** | −0.45 | **−0.40 (−0.61 to −0.19); 0.0015** |
| Colombia, held-out country (S5) | 71% | +0.28 (−0.08 to 0.65); 0.12 | +0.22 | 0.00 (−0.31 to 0.31); 0.99 |
| Brazil, held-out districts (S4) | 81% | +0.20 (−0.26 to 0.66); 0.36 | +0.20 | +0.22 (−0.26 to 0.70); 0.34 |
| Brazil, held-out country (S5) | 81% | **−0.46 (−0.71 to −0.21); 0.0013** | −0.68 | −0.08 (−0.93 to 0.78); 0.85 |
| Peru, held-out country (S5) | 76% | +1.01 (−0.12 to 2.14); 0.075 | +0.93 | +0.78 (−0.51 to 2.08); 0.21 |
| Taiwan, held-out country (S5) | 19% | **−3.28 (−4.77 to −1.79); <0.001** | −18.29 | **−5.11 (−7.17 to −3.05); <0.001** |

*Note:* % of the reference WIS on the kept rows. Brazil's future years, held-out states and held-out regions stay within ±0.25% and are not significant. Peru, future years, is −0.77% (95% CI −1.45 to −0.09; p = 0.029), and Taiwan, future years, −0.51% (p = 0.32). Rows kept are shares of each country's primary rows. Bold: CI excludes zero.

**Panel B.** Change from L3 + C* to L4 + C* in 80% interval coverage on weeks with cases, and in mean 80% interval width (log(1 + cases) scale) on weeks with and without cases.

| Comparison | Coverage on case weeks, L3 + C* | Change in coverage on case weeks, pp (p) | Change in width, zero weeks | Change in width, case weeks |
|---|---|---|---|---|
| Brazil, future years (S1) | 0.66 | −0.2 (0.31) | **+0.006 (+1.6%)** | −0.004 |
| Brazil, held-out states (S2) | 0.66 | **−0.5 (0.011)** | **+0.009 (+2.6%)** | **−0.008** |
| Brazil, held-out regions (S3) | 0.66 | +0.2 (0.088) | **+0.027 (+7.5%)** | +0.002 |
| Brazil, held-out districts (S4) | 0.66 | −0.2 (0.35) | **+0.009 (+2.7%)** | −0.003 |
| Brazil, held-out country (S5) | 0.64 | +0.4 (0.27) | **+0.042 (+9.8%)** | **+0.017** |
| Colombia, future years (S1) | 0.71 | 0.0 (0.94) | **+0.017 (+5.3%)** | +0.001 |
| Colombia, held-out states (S2) | 0.71 | −0.8 (0.15) | **+0.019 (+5.8%)** | −0.010 |
| Colombia, held-out regions (S3) | 0.71 | **−1.0 (0.0028)** | **+0.025 (+7.1%)** | **−0.014** |
| Colombia, held-out districts (S4) | 0.71 | −0.4 (0.41) | **+0.017 (+5.2%)** | −0.004 |
| Colombia, held-out country (S5) | 0.75 | +0.2 (0.29) | **+0.007 (+2.1%)** | −0.007 |
| Peru, future years (S1) | 0.67 | +0.5 (0.076) | **+0.036 (+12.3%)** | **+0.021** |
| Peru, held-out country (S5) | 0.65 | **+2.1 (<0.001)** | **+0.016 (+6.9%)** | **+0.041** |
| Taiwan, future years (S1) | 0.24 | −0.9 (0.25) | +0.002 (+6.4%) | −0.021 |
| Taiwan, held-out country (S5) | 0.57 | +3.7 (0.070) | **+0.093 (+55%)** | **+0.071** |

*Note:* Absolute changes are week-weighted differences, L4 + C* minus L3 + C*, with p values from the main test. Percentages in brackets are the relative change in the mean interval width on zero weeks, as a row mean (L4 + C* against L3 + C*). Bold: 95% CI excludes zero. Intervals on zero weeks already contain the observation more often than intended (about 96% to 100%). Estimates as in Table 2 (seeds 0 to 2 where three seeds exist).

The Colombian losses do not come only from zero weeks. On district-years with at least one case, the excess gain is still about −0.3% and significant in the future-years, held-out-states and held-out-districts splits. The embedding gain is significantly negative in all four within-country splits. The loss shrinks by a fifth to a half on these rows, so part of it lies in district-years without cases. With Taiwan held out, the loss falls from −18.29% to −3.28% on district-years with cases, so most of it lies in district-years with no local case. Brazil's held-out districts stay at +0.20% ($p = 0.36$). We also examined the forecasts themselves, without scoring them, separately for weeks with and without cases. On zero weeks, the embeddings widened the 80% intervals in all 14 comparisons. The increase was 1.6% to 12.3%, and 55% with Taiwan held out. The CI was above zero in 13 of them, although those intervals already contained the observation more often than intended. The embeddings moved coverage on weeks with cases significantly towards the nominal 80% in only one comparison (Peru held out, +2.1 percentage points). They moved it significantly away in two (Brazil's held-out states and Colombia's held-out regions). The embeddings therefore raised the upper forecast quantiles in weeks without cases and did not correct the under-coverage in epidemic weeks.

#### G7. Stability of the embeddings

**Table S49. Year-to-year similarity of a district's embedding compared with similarity between districts.**

| Country | Same district, consecutive years | Same district, first vs last year | Five nearest neighbouring districts, same year | Random district pairs | Next-year vector is the district's own nearest match | Next-year similarity above all five neighbours |
|---|---|---|---|---|---|---|
| Brazil | 0.982 | 0.958 | 0.974 | 0.70 | 57% | 87% |
| Colombia | 0.976 | 0.940 | 0.931 | 0.64 | 84% | 95% |
| Peru | 0.985 | 0.973 | 0.912 | 0.49 | 94% | 98% |
| Taiwan | 0.978 | 0.972 | 0.971 | 0.88 | 69% | 89% |

*Note:* Median cosine similarity of the rescaled, population-weighted district means of the AlphaEarth embeddings, from 2017 to the last extracted year. The last two columns are shares of district-year pairs.

In Brazil and Taiwan a district's embedding changes from year to year by about as much as it differs from its neighbours. The median similarities were 0.982 vs 0.974 in Brazil and 0.978 vs 0.971 in Taiwan. The next year's vector is the district's own nearest match only 57% and 69% of the time. In Colombia and Peru the embedding picks out each district well (84% and 94%). Year-to-year drift was therefore a possible explanation for small losses in Brazil and Taiwan. It does not explain Colombia, where the embedding picks out each district well and the losses are clearest. The fixed embedding built only from early years (V3, Table S51) did not remove the losses either.

#### G8. Information probe

**Table S50. Cross-sectional prediction of district dengue burden from embeddings, coordinates and static covariates.**

| Country (districts) | G | S | E | G + S | G + S + E | Added by E over G + S, T1 (95% CI) | Added by E over G + S, T2 (95% CI) | Ridge: added by E over G + S, T1 (95% CI) |
|---|---|---|---|---|---|---|---|---|
| Brazil (5,517) | 0.06 | 0.65 | 0.49 | 0.71 | 0.72 | +0.010 (−0.002 to 0.025) | +0.009 (−0.010 to 0.046) | +0.169 (−0.050 to 0.724) |
| Colombia (1,070) | −0.17 | 0.66 | 0.52 | 0.71 | 0.72 | +0.010 (−0.018 to 0.042) | −0.006 (−0.024 to 0.019) | +0.423 (0.015 to 1.444) |
| Peru (115) | −0.16 | 0.43 | 0.24 | 0.45 | 0.38 | −0.071 (−0.267 to 0.070) | −0.015 (−0.140 to 0.097) | +0.299 (−0.123 to 0.917) |
| Taiwan (285) | −0.38 | 0.55 | 0.45 | 0.33 | 0.40 | +0.070 (−8.20 to 0.399) | −0.104 (−1.02 to 0.047) | −0.205 (−20.1 to 3.10) |

*Note:* Out-of-fold $R^2$ of a small LightGBM (31 leaves, 200 rounds) predicting T1, the mean log(1 + weekly cases) of 2014 to 2019. Cross-validation holds out whole states, with five folds built as in the held-out-states split. G, coordinates with spherical harmonics (the C2 inputs); S, 2017 to 2018 means of the L3 static covariates and log population in 2018; E, the 2018 AlphaEarth embedding (64 dimensions, rescaled). T2 is the mean log(1 + weekly cases) over the test years. The 95% CIs come from 1,000 bootstrap resamples of states, applied to the fixed out-of-fold predictions, so they ignore variation from refitting. Taiwan's CIs are unstable because resamples without Kaohsiung and Tainan have almost no variance. The ridge column is shown for completeness only. Ridge regression on the 82 coordinate columns extrapolates badly to held-out states ($R^2$ of G alone −0.20 to −0.44 for T1), so its added $R^2$ mostly reflects ridge failing on G + S rather than information in E. In Taiwan, adding coordinates lowered $R^2$ below that of S alone (0.33 against 0.55), and in Peru it raised it only slightly (0.45 against 0.43), so there the increment over G + S is measured against a weak baseline. Measured over S alone, E also added nothing in either country (Peru −0.11, 95% CI −0.32 to 0.03; Taiwan −0.01, −9.92 to 0.18). These are cross-sectional associations. S and E overlap the T1 period; T2 does not.

The embeddings carry information about district dengue burden. On their own they explained 0.49, 0.52, 0.24 and 0.45 of the between-district variance of T1 in held-out states of Brazil, Colombia, Peru and Taiwan. This is less than the static covariates explained (0.43 to 0.66) and far above coordinates alone (−0.38 to 0.06), which in a tree model cannot extrapolate to states not used for fitting. Added to coordinates and static covariates, however, the embeddings raised $R^2$ by at most about 0.01 in Brazil and Colombia, with CIs that include zero. The reduction to 8 principal components gave the same result. In Peru and Taiwan the added $R^2$ was negative or near zero, with wide CIs. When a whole country was left out, the embeddings predicted its burden weakly or not at all. The T1 $R^2$ was −0.34, 0.28 and 0.27 with Brazil, Colombia and Peru held out, and below zero with Taiwan held out.

##### G9. Representation variants

We tried four other ways of using the embeddings in place of the 64 columns. They were added after review and are exploratory.

- V1, similarity risk, a single column: the average past dengue level of the k = 5 training districts with the most similar embeddings. Similarity is cosine similarity, and closer districts get more weight (inverse-dissimilarity weights). Past dengue level is the mean log(1 + cases) over the years before training. The district itself is excluded, and in held-out states and regions the same matched leave-out as for average past cases (C1) is used. V1 is the embedding counterpart of C1.
- V2, the first 8 principal components of the embeddings, fitted on training districts for each origin and fold.
- V3, a fixed embedding built only from early years: the rescaled mean of the 2017 and 2018 vectors, the same in every year.
- V4, substitution: the embeddings in place of the remote-sensing and population inputs of L3, that is, L2 plus embeddings.

All variants use average past cases (C1) as the selected control, the shared configurations, seed 0 and horizons 1 and 4.

**Table S51. Embedding and excess gains with alternative representations of the embeddings.**

| **Comparison** | **Original 64 columns** | **V1, risk encoding** | **V2, PCA-8** | **V3, causal static** |
|---|---|---|---|---|
| *Embedding gain, % (95% CI); p (Holm p)* | | | | |
| Colombia, future years (S1) | −0.36; 0.0052 | −0.07 (−0.13 to −0.01); 0.023 (1.0) | −0.07 (−0.17 to 0.02); 0.12 (1.0) | −0.14 (−0.25 to −0.03); 0.017 (1.0) |
| Colombia, held-out states (S2) | −0.59; <0.001 | −0.04 (−0.11 to 0.03); 0.19 (1.0) | **−0.14 (−0.22 to −0.07); <0.001 (0.027)** | **−0.30 (−0.40 to −0.20); <0.001 (0.001)** |
| Peru, future years (S1) | −0.94; 0.020 | −0.01 (−0.13 to 0.10); 0.81 (1.0) | −0.15 (−0.64 to 0.35); 0.53 (1.0) | −0.66 (−1.19 to −0.13); 0.018 (1.0) |
| Taiwan, future years (S1, with 2021) | −1.06; 0.024 | −0.66 (−1.76 to 0.45); 0.23 (1.0) | −0.46 (−1.71 to 0.78); 0.44 (1.0) | −0.60 (−1.58 to 0.37); 0.21 (1.0) |
| Brazil, held-out states (S2) | −0.02; 0.88 | −0.05 (−0.11 to 0.01); 0.10 (1.0) | −0.02 (−0.17 to 0.12); 0.75 (1.0) | not run |
| *Excess gain beyond C1, % (95% CI); p (Holm p)* | | | | |
| Colombia, future years (S1) | −0.39; 0.0011 | −0.05 (−0.09 to −0.01); 0.014 (1.0) | −0.13 (−0.24 to −0.02); 0.025 (1.0) | −0.08 (−0.19 to 0.03); 0.12 (1.0) |
| Colombia, held-out states (S2) | −0.47; <0.001 | −0.01 (−0.05 to 0.03); 0.68 (1.0) | −0.11 (−0.18 to −0.05); 0.0033 (0.085) | **−0.21 (−0.31 to −0.12); <0.001 (0.015)** |
| Peru, future years (S1) | −0.70; 0.056 | +0.04 (−0.10 to 0.19); 0.53 (1.0) | +0.06 (−0.41 to 0.54); 0.77 (1.0) | **−0.87 (−1.32 to −0.42); 0.0012 (0.032)** |
| Taiwan, future years (S1, with 2021) | −0.60; 0.049 | −0.11 (−0.29 to 0.07); 0.20 (1.0) | +0.15 (−0.53 to 0.83); 0.65 (1.0) | −0.12 (−0.32 to 0.08); 0.22 (1.0) |
| Brazil, held-out states (S2) | 0.00; 1.0 | −0.02 (−0.06 to 0.02); 0.31 (1.0) | +0.05 (−0.12 to 0.21); 0.56 (1.0) | not run |

| **Comparison** | **V1 vs C1: WIS(L3 + C1) − WIS(L3 + V1), % (95% CI); p** | **V4 substitution: WIS(L3) − WIS(L2 + embeddings), % (95% CI); p (Holm p)** | **WIS(L2) − WIS(L2 + embeddings), % (95% CI); p** |
|---|---|---|---|
| Colombia, future years (S1) | −0.01 (−0.23 to 0.21); 0.94 | **−0.79 (−1.15 to −0.42); <0.001 (0.018)** | −0.48 (−0.80 to −0.17); 0.0062 |
| Colombia, held-out states (S2) | +0.04 (−0.004 to 0.08); 0.064 | **−0.93 (−1.27 to −0.59); <0.001 (0.003)** | no L2 run |
| Peru, future years (S1) | +0.24 (−0.48 to 0.97); 0.48 | −0.84 (−1.66 to −0.01); 0.047 (0.93) | −1.10 (−1.90 to −0.31); 0.010 |
| Taiwan, future years (S1, with 2021) | −0.03 (−0.20 to 0.15); 0.74 | −0.79 (−1.77 to 0.20); 0.11 (1.0) | −0.74 (−1.37 to −0.11); 0.024 |
| Brazil, held-out states (S2) | 0.00 (−0.07 to 0.07); 0.91 | not run | not run |

*Note:* % of WIS(L3); a positive value means that the variant reduced error. Holm adjustment is within the family of 32 variant tests named in the specification: the embedding and excess gains of V1 to V3 and the substitution contrast. The contrasts of V1 with C1 and with L2 are outside that family. Bold: Holm $p < 0.05$. The first column repeats the primary seed-0 values.

No variant turned the embeddings into a significant gain. None of the 28 tests of embedding and excess gains had a CI above zero, and the largest point estimate was +0.15%. The compact forms V1 (one column) and V2 (eight columns) shrank the loss towards zero. In Colombia, future years, the embedding gain was −0.07% for both, against −0.36% with the full embedding. Part of the loss with 64 columns is therefore a cost of having many inputs, but the compact forms did not reverse it. V1 could not be told apart from average past cases in any comparison (largest difference +0.04%, $p = 0.064$). Similarity in embedding space thus carried risk from neighbours about as well as geographic closeness did, and no better. The fixed early-years embedding V3 did not remove the loss. Its excess gain was −0.21% in Colombia's held-out states and −0.87%

in Peru's future years (Holm p = 0.032), so year-to-year changes in the vector do not explain the whole loss. Used in place of the remote-sensing and population inputs, the embeddings were worse than L3 in all four comparisons (Colombia −0.79% and −0.93%, both Holm-significant). In Colombia, Peru and Taiwan they were also worse than L2, which has no place inputs at all, so in this model they did not replace the hand-built covariates. Every Holm-significant variant result is a loss, and all lie within or near ±1%.

#### G10. Seeds

**Table S52. Excess, embedding and control gains averaged over seeds 0 to 2, and seed 0 alone.**

| Comparison | C* | Excess gain, seeds 0 / 1 / 2 | Excess gain, mean of seeds 0–2 (95% CI); p | Seed SD | Holm p, seed 0 (13 comparisons) → seeds 0–2 (14 comparisons) | Embedding gain, seeds 0–2 (95% CI); p | Control gain, seeds 0–2 (95% CI); p |
|---|---|---|---|---|---|---|---|
| Brazil, held-out country (S5) | C3 | −0.60 / −0.62 / −0.82 | −0.68 (−0.94 to −0.42); <0.001 | 0.12 | <0.001 → <0.001 | −0.39 (−1.34 to 0.55); 0.39 | +0.15 (−0.89 to 1.18); 0.77 |
| Colombia, future years (S1) | C1 | −0.39 / −0.48 / −0.40 | −0.42 (−0.64 to −0.21); 0.0012 | 0.05 | 0.012 → 0.014 | −0.38 (−0.61 to −0.16); 0.0035 | −0.06 (−0.32 to 0.19); 0.60 |
| Colombia, held-out states (S2) | C1 | −0.47 / −0.58 / −0.52 | −0.52 (−0.74 to −0.30); <0.001 | 0.05 | 0.0068 → 0.0034 | −0.60 (−0.81 to −0.39); <0.001 | −0.06 (−0.09 to −0.03); 0.0019 |
| Colombia, held-out regions (S3) | C1 | −0.65 / −0.70 / −0.70 | −0.68 (−1.08 to −0.29); 0.0030 | 0.03 | 0.026 → 0.030 | −0.82 (−1.14 to −0.49); <0.001 | −0.08 (−0.14 to −0.02); 0.011 |
| Colombia, held-out districts (S4) | C1 | −0.43 / −0.48 / −0.42 | −0.45 (−0.68 to −0.21); 0.0017 | 0.03 | 0.025 → 0.019 | −0.54 (−0.78 to −0.30); <0.001 | −0.10 (−0.12 to −0.08); <0.001 |
| Peru, future years (S1) | C1 | −0.70 / −0.88 / −0.87 | −0.82 (−1.49 to −0.14); 0.022 | 0.10 | 0.34 → 0.18 | −0.91 (−1.62 to −0.20); 0.016 | −0.20 (−0.92 to 0.52); 0.56 |
| Peru, held-out country (S5) | C3 | +0.96 / +0.96 / +0.88 | +0.93 (−0.16 to 2.02); 0.088 | 0.04 | 0.39 → 0.53 | +0.68 (−0.59 to 1.96); 0.27 (seed 0) | −0.44 (−0.80 to −0.07); 0.022 (seed 0) |
| Taiwan, future years (S1, with 2021) | C1 | −0.60 / −0.87 / −0.67 | −0.71 (−1.40 to −0.03); 0.042 | 0.14 | 0.34 → 0.29 | −0.95 (−1.71 to −0.18); 0.018 | −0.41 (−0.97 to 0.16); 0.15 |
| Taiwan, held-out country (S5, with 2021) | C3 | −17.30 / −18.06 / −19.51 | −18.29 (−29.80 to −6.78); 0.0039 | 1.12 | 0.017 → 0.035 | −21.60 (−31.82 to −11.39); <0.001 (seed 0) | −3.36 (−6.52 to −0.20); 0.039 (seed 0) |

*Note:* % of WIS(L3), horizons 1 and 4. Where three seeds exist, we averaged the per-row loss over the seeds that both models of a gain share and then applied the main test. Seed SD is the standard deviation of the three per-seed estimates, % of WIS. With Peru or Taiwan held out, only L3 + C* and L4 + C* have seeds 1 and 2, because of a limit on computing time set in advance, so their embedding and control gains stay at seed 0. Brazil's future-years comparison already used three seeds (Table 2). Brazil's held-out states, regions and districts and Colombia held out have seed 0 only. Holm adjustment for seed 0 is over the 13 comparisons available before the result for Brazil's held-out districts had been seen; for seeds 0 to 2 it is over all 14 comparisons, as in Table 2. The three-seed values are the primary estimates of Table 2 (row 53).

Averaging over three seeds changed no conclusion after Holm adjustment. The same six comparisons remain significant losses: Brazil held out, Colombia's four within-country splits and Taiwan held out. Every loss grew slightly, by 0.01 to 0.12 percentage points and by 1.0 with Taiwan held out, and no seed reversed a sign. Under the strict-variance check (B = 4) with Holm adjustment, however, the loss with Brazil held out is no longer significant (p = 0.095). Only Colombia, held-out states, is therefore significant under the main test and both stricter checks (Table S26). The seed SD of the excess gain (0.03% to 0.14% of WIS; 1.1% with Taiwan held out) is small next to these effects. In Colombia every seed was negative and significant on its own, so the Colombian losses are not seed noise. Before adjustment, Peru, future years, moved from p = 0.056 to p = 0.022, and its CI now excludes zero; it is not significant after Holm adjustment (p = 0.18). With Peru held out the gain stays positive but not significant (+0.93%, p = 0.088), with every seed positive. The control gain is the quantity most sensitive to the seed. Seed 1 brought it to about zero in Colombia's held-out regions and districts and in Taiwan, future years.

#### G11. Taiwan future-years ladder including 2021

**Table S53. Baselines, ladder and fixed control panel for Taiwan, future years, with test year 2021.**

**Panel A.** Ladder and baselines (row mean; 141,645 rows).

| Model | WIS | Relative WIS vs L0 | 80% coverage, all rows / cases > 0 | Relative WIS vs L0 without 2021 |
|---|---|---|---|---|
| L0, seasonal naive | 0.0743 | 1.00 | 0.95 / 0.00 | 1.00 |
| Negative binomial regression (L1 inputs) | 0.0475 | 0.64 | 0.99 / 0.42 | 0.64 |
| LightGBM L1, case history | 0.0316 | 0.43 | 0.98 / 0.24 | 0.43 |
| LightGBM L2, + climate (shared configuration) | 0.0319 | 0.43 | 0.98 / 0.23 | not run (the earlier L2 was tuned separately, 0.52) |
| LightGBM L3, + remote sensing and population | 0.0319 | 0.43 | 0.98 / 0.25 | 0.43 |
| LightGBM L4, + embeddings | 0.0323 | 0.43 | 0.98 / 0.24 | 0.44 |
| LightGBM L3 + C* (C1) | 0.0321 | 0.43 | 0.98 / 0.24 | 0.43 |
| LightGBM L4 + C* (C1) | 0.0323 | 0.43 | 0.98 / 0.24 | 0.44 |

**Panel B.** Fixed control panel (seed 0; no multiplicity adjustment).

| Control | Control gain, % (95% CI); p | Excess gain, % (95% CI); p | Excess gain without 2021; p |
|---|---|---|---|
| C1, average past cases | −0.63 (−1.68 to 0.43); 0.22 | −0.60 (−1.20 to −0.003); 0.049 | −0.44; 0.12 |
| C2, coordinates | −0.36 (−0.76 to 0.03); 0.070 | −0.94 (−2.03 to 0.15); 0.087 | −0.77; 0.16 |
| C3, shuffled embedding | −0.58 (−1.29 to 0.12); 0.097 | −0.68 (−1.45 to 0.09); 0.079 | −0.54; 0.15 |
| C6, district label | −2.29 (−5.59 to 1.02); 0.16 | −0.52 (−1.16 to 0.13); 0.11 | −0.42; 0.18 |
| none (embedding gain) | – | −1.06 (−1.96 to −0.16); 0.024 | −0.94; 0.043 |

*Note:* Every step of the ladder, control in the panel and gain from added predictors is scored on the same 141,645 rows as Table 2. TimesFM was not run for Taiwan. The 2021 targets are all zero, so adding 2021 lowers every absolute WIS but leaves the ordering and the relative WIS almost unchanged. Seed 0; Table S17 averages seeds 0 to 2 for L3, L4, L3 + C* and L4 + C*, so its values differ by up to 0.0001.

With 2021 included, all Taiwan tables use the same rows. Every LightGBM model forecast Taiwan about 57% better than the seasonal naive baseline, and nothing added to case history helped. L1 had the lowest WIS of the LightGBM ladder, and the steps adding inputs other than case history had no detectable effect (Table S44). All four fixed controls gave a negative excess gain, significant before adjustment only with C1 (p = 0.049), and none of them improved L3. Tables S17 to S20 and S36 use these values with 2021 included.

#### G12. Colombia held-out states without case history

**Table S54. Embedding, control and excess gains without case history, Colombia, held-out states.**

| Estimand | Colombia, held-out states: % (95% CI); p | Brazil, future years (Table S42) |
|---|---|---|
| Embedding gain, N3 → N4 | **−5.30 (−8.19 to −2.41); 0.0020** | +2.54; 0.13 |
| Control gain, N3 → N3 + C2 (coordinates, C*) | **−2.08 (−3.47 to −0.69); 0.0072** | +3.30; <0.001 |
| Excess gain, N3 + C2 → N4 + C2 | **−4.17 (−6.45 to −1.90); 0.0019** | −0.43; 0.58 |
| Static history, N3 → N3 + C1 | **−1.46 (−2.90 to −0.03); 0.047** | – |
| Excess gain beyond C1, N3 + C1 → N4 + C1 | **−3.46 (−6.13 to −0.79); 0.016** | – |

| Model (row mean, 319,930 rows) | N3 | N3 + C1 | N3 + C2 | N4 | N4 + C1 | N4 + C2 |
|---|---|---|---|---|---|---|
| WIS | 0.1989 | 0.2019 | 0.2030 | 0.2096 | 0.2089 | 0.2115 |
| 80% coverage, all rows / cases > 0 | 0.91 / 0.64 | 0.90 / 0.64 | 0.89 / 0.62 | 0.88 / 0.63 | 0.87 / 0.62 | 0.88 / 0.63 |

*Note:* % of the WIS of N3 (week-weighted 0.1997), horizons 1 and 4, test years 2020 to 2022, seed 0, Colombia's shared configuration. The control was selected at 2019 by the main selection rule among C2, C3, C4 and C4s. Coordinates (C2) were the only member of the tie set (0.33733 vs 0.34366 for C4 and 0.34467 for C3), although they did not beat N3 (0.33728). The minimum detectable excess gain is 3.18%. The control share cannot be estimated, because the embedding gain is negative. Exploratory, outside the primary family. Bold: CI excludes zero.

The Brazilian pattern without case history did not appear in Colombia. In Colombia's held-out states without case history, the embeddings made forecasts clearly worse (about −4% to −5%), with or without a location control, and neither coordinates nor static history helped N3. In both countries the embeddings added nothing beyond a location control. There are only three test years and five folds, and place inputs generally reduced accuracy in held-out Colombian states. The results without case history therefore differ between countries, are exploratory and say little about mechanism.

#### G13. Equal-budget tuning

**Table S55. Excess gain in Colombia, future years, with each model tuned separately on an equal grid.**

| Model | Configuration chosen (leaves / minimum rows per leaf / feature fraction) | Validation pinball loss, 2019 | Excess gain, % (95% CI); p |
|---|---|---|---|
| L3 + C1, tuned separately | 31 / 100 / 0.9 | 0.07114 | – |
| L4 + C1, tuned separately | 31 / 100 / 0.9 | 0.07174 | – |
| Best for each | – | – | −0.39 (−0.58 to −0.19); 0.0011 |
| Shared configuration (primary, seed 0) | 31 / 100 / 0.9 | – | −0.39 (−0.58 to −0.19); 0.0011 |

*Note:* Grid of 18 configurations (leaves 15, 31, 127; minimum rows per leaf 100, 1,000, 5,000; feature fraction 0.3, 0.9; 400 rounds), trained on 2018 and validated on 2019. The second-best setting was 15 / 100 / 0.9 for both models (+0.6% and +0.4% validation loss). Seed 0.

Tuned separately on the same grid, each model chose the shared configuration. The test-year forecasts were therefore exactly the same as the shared ones, and the excess gain was unchanged (−0.39%). In Colombia the loss does not come from using one shared configuration.

##### G14. Hidden-case check (added during the final author review)

Unlike the other analyses in this section, this one was added after all 14 comparisons had been read, at a co-author's request (row 54 of Table S14 in the Supplementary Methods). The hidden-case check assumes that some zero-filled weeks hid unreported cases and asks how many such cases it would take to remove each loss. Each draw is one random choice of which filled weeks hide cases. Nothing was refitted: the stored forecasts of both models were rescored against changed targets.

**Table S56. Hidden-case check: excess gain when a share of zero-filled test weeks is changed to 1 or 3 cases.**

| Comparison | Excess gain as reported, % (Holm p) | Median excess gain (%), 2.1% of filled weeks changed, 1 case | Median excess gain (%), 10% changed, 1 case | Draws with a Holm-significant loss at 2.1% (of 20), 1 / 3 cases | Tipping share, %, 1 / 3 cases |
|---|---|---|---|---|---|
| Brazil, held-out country | −0.68 (<0.001) | −0.60 | −0.27 | 20 / 20 | 10 / 10 |
| Colombia, future years | −0.42 (0.014) | −0.34 | −0.01 | 16 / 10 | 5 / 2.1 |
| Colombia, held-out states | −0.52 (0.003) | −0.42 | −0.02 | 20 / 20 | 5 / 5 |
| Colombia, held-out regions | −0.68 (0.030) | −0.53 | +0.01 | 0 / 0 | 2 / 2 |
| Colombia, held-out districts | −0.45 (0.019) | −0.35 | +0.01 | 6 / 0 | 2 / 2 |
| Taiwan, held-out country | −18.29 (0.035) | −15.54 | −5.19 | 0 / 0 | 2 / 2 |

*Note:* The six comparisons with a loss in Table 2. Rows, exclusions, seeds and tests as in Table 2 (horizons 1 and 4 pooled; forecasts whose origin lies in the target's epidemiological year; Holm adjustment over the 14 comparisons). In each draw, a share of the zero-filled district-weeks in the test years is changed from 0 to 1 or 3 cases; weeks the source listed as 0 are never changed. A changed week changes at both horizons, for all four models and every seed, and the stored forecasts are rescored; no model is refitted. The shares were 0.5%, 1%, 2%, 2.1%, 5% and 10%, with 20 draws for each share and case count. Weeks were drawn uniformly at random within each comparison. Within a draw, the weeks changed at a smaller share are also changed at every larger share, and the same weeks are used for 1 and 3 cases. Percentages use the unperturbed reference WIS as the denominator, and gains are medians over draws. Each draw is tested with the main test, with Holm adjustment over the 14 comparisons within that draw. The tipping share is the smallest share tested at which the loss is no longer Holm-significant in at least half of the draws; because Holm is applied within each draw, a loss can also tip when the other losses weaken at the same time. The benchmark is Brazil's check against InfoDengue, in which at most 2.1% of filled weeks in a sampled year held a probable case and none held more than 8 (Table S4); 8 cases per week was not tested. The test changes only the scored targets. Hidden cases would also have entered the case-history inputs and the training targets of both models, which are not changed, and district-level rates of hidden cases in Colombia and Peru remain unknown.

At Brazil's highest observed rate of 2.1%, all six losses shrank by 13% to 22%, and each stayed significant before adjustment in every draw. After Holm adjustment, only the losses with Brazil held out and in Colombia's held-out states stayed significant in every draw; at 10%, no loss did, and the four Colombian losses were close to zero while the losses with Brazil and Taiwan held out stayed negative. Brazil's benchmark applies most directly to the comparison with Brazil held out, Taiwan's filled weeks held no local case in the surveillance data (Supplementary Methods, section A4), and Colombia's rate is unknown.

The other eight comparisons, the other shares, every result for 3 cases and the embedding gain are in the results file docs/results/review/R5_zero_fill_stress.csv. No comparison became a Holm-significant gain at any share, for the excess gain or the embedding gain.

### H. Supplementary figures

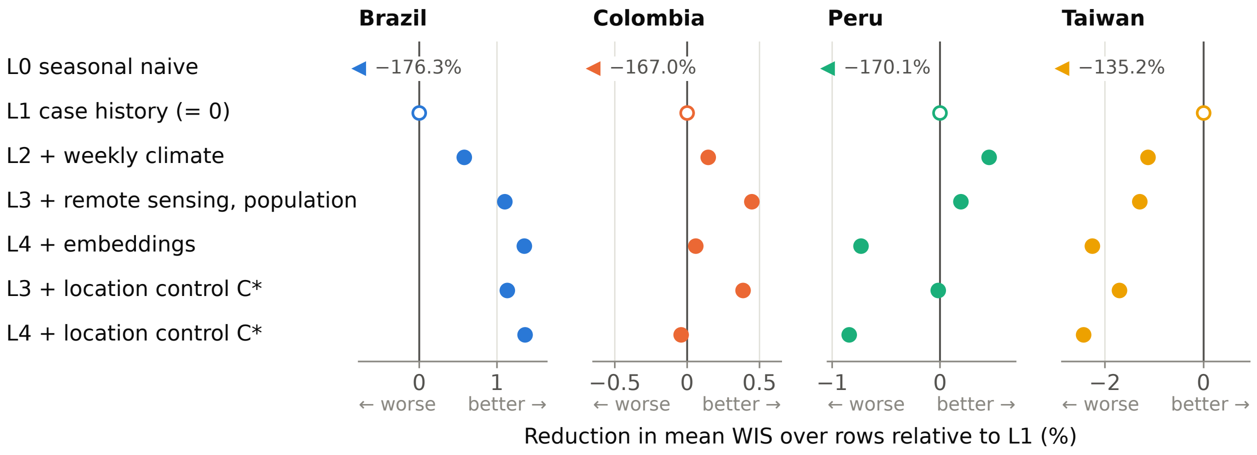

**Figure S1. Forecast error along the model ladder in the future-years split (S1), by country.** The figure shows how much each model reduces mean WIS over rows compared with L1, the model with case history only (open marker at 0), with horizons 1 and 4 pooled. Models are L0 to L4 and L3 and L4 with the selected location control, which is average past cases (C1) in every country. Values beyond a panel are drawn as an arrow with their value; this applies to the seasonal naive model L0 everywhere. Panels have their own scales.

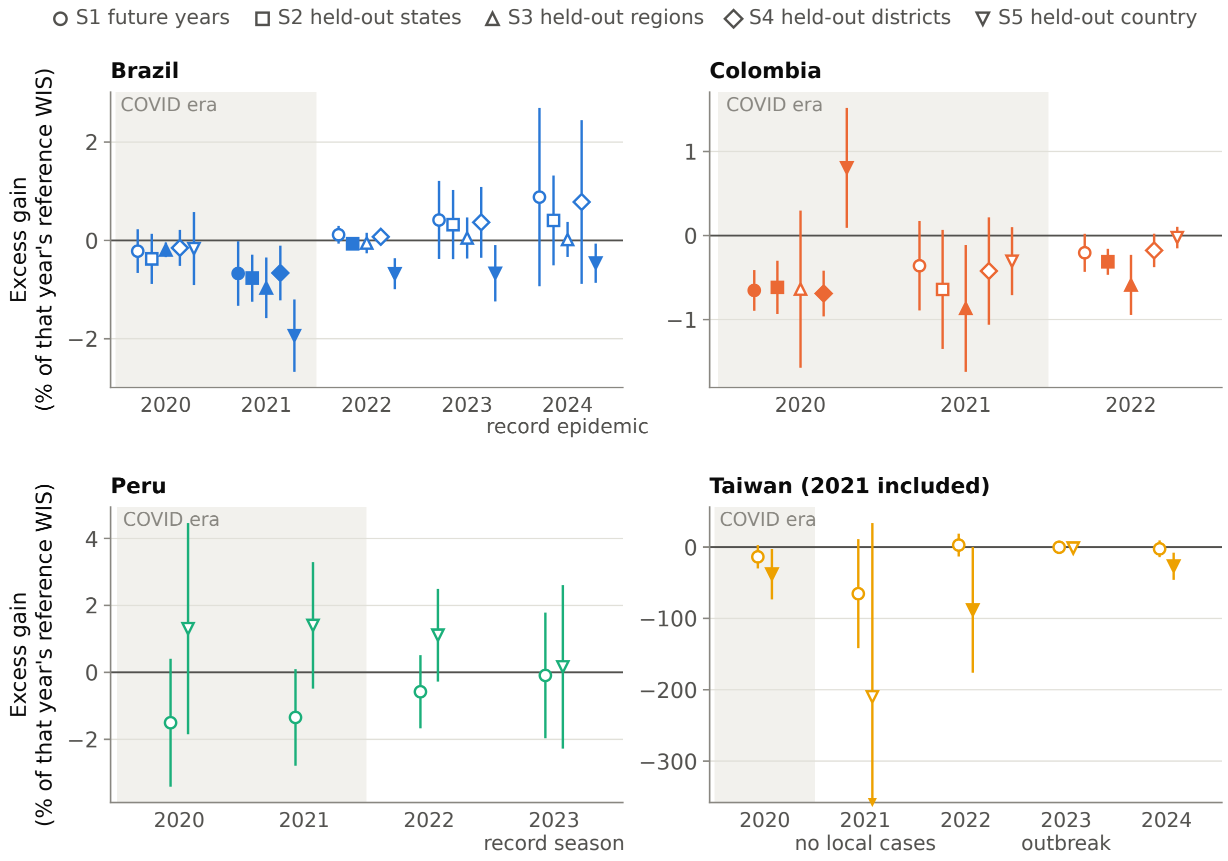

**Figure S2. Excess gain by test year.** Each point is one comparison in one test year, shown as a percentage of that year's reference WIS (weeks weighted equally) with horizons 1 and 4 pooled. Bars are unadjusted 95% confidence intervals; a filled marker means the interval excludes 0, and the marker shape gives the split. Shading marks the COVID-19 pandemic years 2020–2021. In the Taiwan panel only 2020 is shaded, and 2021 is labelled "no local cases". Intervals running past a panel are cut with an arrowhead. Taiwan includes 2021, when there were no local cases, so its reference WIS is tiny and its percentages are very large (absolute values in Table S30). Panels have their own scales.

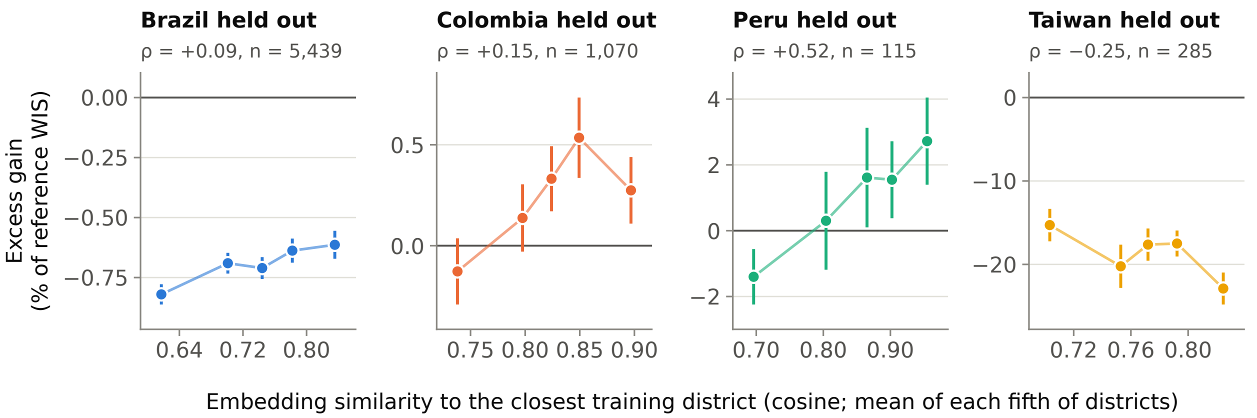

**Figure S3. Excess gain by embedding similarity when a whole country is held out (S5).** The districts of the held-out country are split into fifths by how similar their rescaled AlphaEarth embedding for year y − 1 is to that of the most similar training district (cosine similarity). Points show each fifth's mean similarity and its excess gain as a percentage of the comparison's overall reference WIS, with 95% confidence intervals. ρ is the Spearman correlation between similarity and excess gain across districts; its p values, which treat districts as independent, are in Table S39. Panels have their own vertical scales. Exploratory.

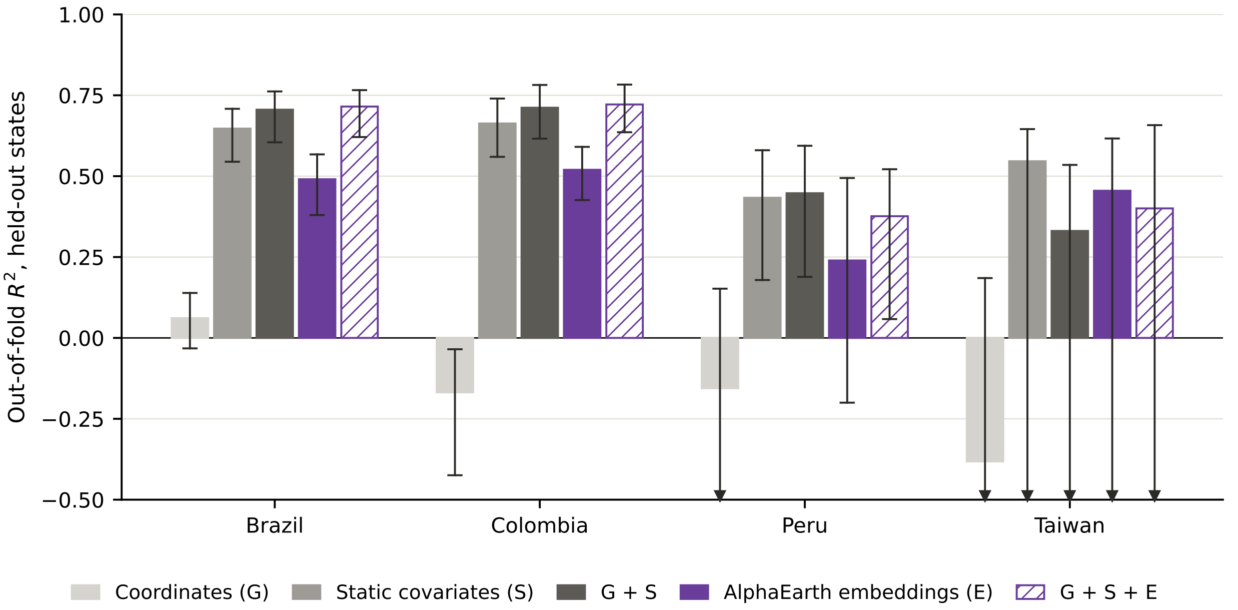

**Figure S4. How well district dengue burden can be predicted from embeddings, coordinates and static covariates in states not used for fitting.** Bars show the out-of-fold $R^2$ of a small LightGBM predicting each district's mean log(1 + weekly cases) over 2014 to 2019, under five-fold cross-validation that holds out whole states. Whiskers are 95% CIs from 1,000 bootstrap resamples of states. Grey bars use no embeddings: coordinates with spherical harmonics (G, light grey), the static covariates of L3 (S, mid grey) and both (G + S, dark grey). Purple bars use the 2018 AlphaEarth embeddings, alone (E, solid) or added to coordinates and static covariates (G + S + E, hatched). The axis is cut at −0.5, and whiskers that run below it end in an arrowhead. This affects Peru G and all of Taiwan, whose bootstrap is unstable because resamples without Kaohsiung and Tainan have almost no variance. Exploratory and cross-sectional; ridge regression results and the added $R^2$ with CIs are in Table S50.
